# CRISPR RNA-independent activation of Cas12a

**DOI:** 10.64898/2026.07.14.26358058

**Authors:** Idorenyin A. Iwe, Serena Singh, Kaoling Guan, Rodrigo Fregoso Ocampo, Severino Jefferson Ribeiro da Silva, Dagwin Wachholz Junior, Nadia Emami, Ariel Corsano, Ilan Zeisler, Kristof Bozovicar, Liu Wang, Dalton Ham, Rita Cai, Paul Kelly, Riham Zayani, Jessica Nguyen, Pouriya Bayat, Moiz Charania, Sean Palter, Frank X. Liu, Suman Shrestha, Ashayad Rayhan, Gregory A. Wasney, Tony Mazzulli, Alexander A. Green, Zhigang Li, Shuhuai Yao, Basil P. Hubbard, David W. Taylor, Keith Pardee

## Abstract

CRISPR-Cas12a nucleases are classically activated through CRISPR RNA (crRNA)-guided and PAM-dependent target recognition, which together establish a canonical heteroduplex associated with nuclease activation. Here we identify a crRNA– and PAM-independent activation pathway for Cas12a that reveals previously unrecognized conformational plasticity within its nucleic acid–recognition interface. We show that short RNAs can directly occupy the canonical crRNA–binding channel and trigger a catalytically competent *trans* cleavage state in the absence of PAM recognition or canonical R-loop formation. Biochemical assays indicate that short RNAs bind the crRNA–binding channel and are competitively displaced by cognate crRNA, consistent with binding at a conserved nucleic acid–binding interface. Cryo-electron microscopy (cryo-EM) further reveals that Cas12a maintains its global catalytic architecture while exhibiting loss of canonical PAM-dependent stabilization and increased flexibility of the RuvC lid, alongside accommodation of a noncanonical RNA–DNA hybrid with inverted polarity relative to the crRNA–target duplex. This crRNA-independent activation pathway enables programmable and amplification-free detection of DNA and RNA targets independent of canonical guide-mediated recognition. Together, these findings define an alternative activation geometry for Cas12a and expand models of Class 2 CRISPR-Cas effector activation beyond crRNA– and PAM-directed recognition.

## Introduction

CRISPR-Cas12a nucleases are activated through a CRISPR-RNA (crRNA)-directed mechanism in which target recognition and PAM engagement induce conformational rearrangements that enable nuclease activity.^1–4^ Unlike the dual RNA-guided architecture used by some Class 2 CRISPR-Cas effectors, including Cas9, Cas12a operates through a comparatively simple crRNA-only recognition system that provides a streamlined framework for coupling target recognition to nuclease activation during bacterial antiviral defense.^5–7^ In this model, target engagement is coupled to allosteric rearrangements within the nuclease lobe that enable single-turnover cleavage of dsDNA (*cis* cleavage), followed by robust multi-turnover degradation of non-specific ssDNA sequences (*trans* cleavage).^3, 8, 9^ This collateral activity arises from a tightly regulated activation pathway in which crRNA-dependent base pairing and PAM recognition jointly stabilize the catalytically competent conformation, including structural rearrangements that regulate access to the RuvC catalytic site.^10^

Structural and biochemical studies have defined this activation process as highly dependent on the integrity of the crRNA scaffold and its occupation of the canonical crRNA–binding channel within Cas12a.^1–4^ The scaffold–spacer architecture of the crRNA is therefore considered central to both target specificity and allosteric activation, with current models assuming that productive nuclease engagement requires formation of a fully guided R-loop intermediate. Consistent with this view, most engineered Cas12a systems preserve guide-dependent architecture, even when modifying target duplex structure or employing surrogate nucleic acid guides.^11–17^ Our recent development of engineered duplex activators further demonstrated that alternative nucleic acid architectures can support Cas12a activation while retaining crRNA-mediated recognition.^18^

However, emerging evidence suggests that Cas12a activation may not be strictly constrained to canonical crRNA-directed recognition and instead reflects a broader conformational landscape responsive to crRNA-independent nucleic acid inputs.^17, 19, 20^ The structural basis for activation by such inputs, specifically how they engage the crRNA–binding channel to induce catalytically competent Cas12a states independent of canonical crRNA architecture and PAM-defined target geometry, remains unknown.

Here we identify a crRNA-independent activation pathway for Cas12a that reveals previously unrecognized conformational plasticity within its nucleic acid–recognition interface, in which short RNAs and DNA– RNA hybrids directly engage the crRNA–binding channel to induce a *trans* cleavage competent state in the absence of canonical R-loop formation (**Fig. 1a**). We define the minimal RNA species capable of supporting this activation as minimal Cas-activating RNAs (miniCaRs), representing a previously unrecognized mode of nucleic acid–driven Cas12a activation that decouples nuclease activation from canonical crRNA and PAM architecture. This novel activation pathway expands the molecular sensing capabilities of Cas12a by enabling direct detection of DNA and RNA targets without canonical guide-mediated recognition.

**Fig. 1.**
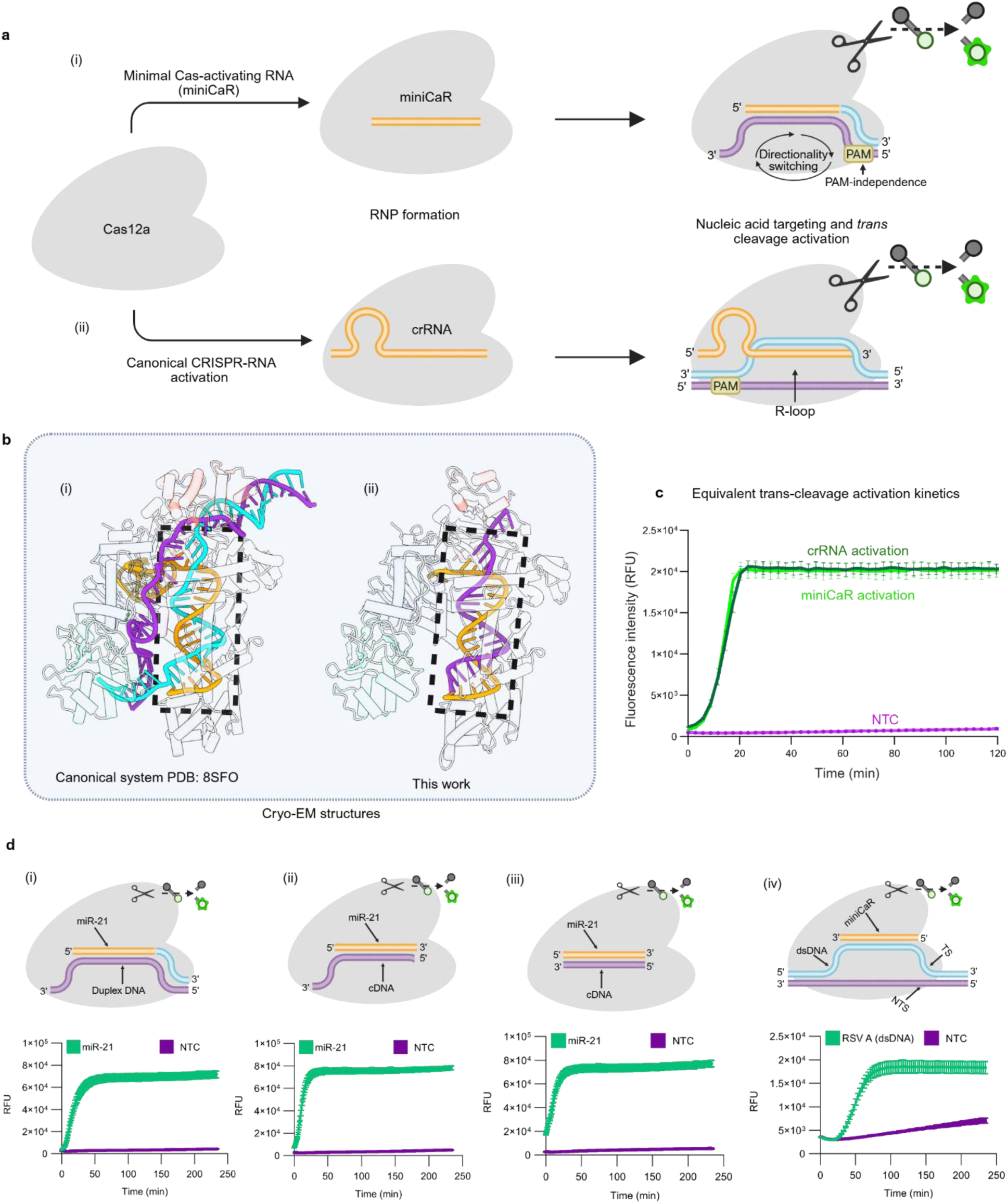
crRNA-independent Cas12a activation by minimal RNA and modes of activation. **(a)** (i) Cas12a forms a crRNA-independent ribonucleoprotein (RNP) with a minimal Cas-activating RNA (miniCaR) and is activated by a partial duplex probe. (ii) In the canonical pathway, Cas12a forms a crRNA-guided RNP, which is activated by a PAM-containing dsDNA target. The duplex probe contains a full NTS (purple) and a short TS segment (cyan), whereas the canonical substrate contains full-length TS and NTS. Both assemblies trigger robust *trans* cleavage of ssDNA reporters. **(b)** Cryo-EM structures of the crRNA-independent complex determined here and a canonical Cas12a–crRNA–DNA complex (PDB: 8SFO^21^). **(c)** Representative fluorogenic reporter kinetics showing strong and comparable *trans* cleavage activity in both systems. **(d)** Four modes of crRNA-free Cas12a configurations: (i) miniCaR activates *trans* cleavage via DNA duplex probe, providing baseline activity. (ii) Longer complementary DNA probe (cDNA > 22 nt) with a 3′ overhang, hybridizes with miniCaR (22 nt) to activate Cas12a *trans* cleavage. (iii) A 22-nt cDNA probe fully complementary to miniCaR (22 nt) activates *trans* cleavage, and (iv) miniCaR hybridizes with PAM-free dsDNA to activate strong *trans* cleavage. NTC is non-target control. Orange is RNA or miniCaR; purple, ssDNA; and cyan, target strand.

## Results

### crRNA– and PAM-independent activation of Cas12a by minimal Cas-activating RNAs

We began by optimizing CRISPR-Cas12a reaction conditions using our previously developed RAPID (RNA/DNA Advanced chimeric, PAM-independent, Integrated Nicking, Diagnostics) platform as a starting point, which detects RNA targets through a complementary split-activator architecture.^18^ During these experiments, which involved short RNAs as targets, we evaluated the effect of crRNA concentration on Cas12a *trans* cleavage activity. Unexpectedly, we observed that varying crRNA concentration had no impact on the fluorescent readout, and reactions lacking crRNA exhibited *trans* cleavage activity comparable with those containing saturating crRNA levels (**Suppl. Fig. 1**). This enabled us to identify a previously unrecognized mode of CRISPR-Cas12a activation in which short RNA sequences can directly trigger *trans* cleavage activity in the absence of a crRNA (**Fig. 1d**). In the canonical model, Cas12a requires a crRNA to direct sequence recognition and PAM-dependent target engagement, which together induce conformational rearrangements that activate its RuvC nuclease domain.^1–4^ Here, however, we show that short RNA sequences can activate Cas12a without a crRNA, producing robust collateral ssDNA cleavage of reporter substrates (**Suppl Fig. 1b**).

We define these activating species as miniCaRs, short RNAs that directly trigger Cas12a catalytic activation without requiring scaffold–spacer crRNA architecture (**Fig. 1d**). Systematic probing revealed that miniCaRs can activate Cas12a through multiple mechanisms, including interactions with ssDNA (**Fig. 1d (ii, iii)**) or dsDNA (**Fig. 1d(iv)**), as well as duplex-assisted activation by partially complementary nucleic acid probes (**Fig. 1d(i)**), demonstrating compatibility with diverse nucleic acid architectures across optimised reaction and temperature conditions (**Suppl Fig. 2 and 3**). Additional chimeric partial duplex and dsDNA systems were also identified (**Suppl Fig. 4a-c, Extended Data Fig. 1**). Importantly, collectively, these observations establish that Cas12a activation is not strictly dependent on crRNA-mediated targeting but can instead be initiated by alternative RNA inputs.

To further examine whether crRNA-independent activation depends on protospacer adjacent motif (PAM) recognition, we tested duplex probes containing a range of PAM sequences embedded within the double-stranded region. These included the canonical 5′-TTTV^22^ motif, several previously reported suboptimal PAMs (e.g., GTTC, CTTA, TTGT, TTGG, and TCTC),^23^ as well as sequences lacking PAM-like features altogether (e.g., TTTT, AAAA, CCCC, and GGGG) (**Fig. 2a**; **Suppl Fig. 5**; **Table 5**). Across all tested conditions, AsCas12a activation remained consistently high and showed no measurable dependence on PAM identity or presence. These results indicate that crRNA-independent activation by miniCaRs is fully PAM-independent, further distinguishing this mechanism from canonical target recognition pathways that rely on PAM-driven DNA interrogation.^10, 24, 25^

**Fig. 2.**
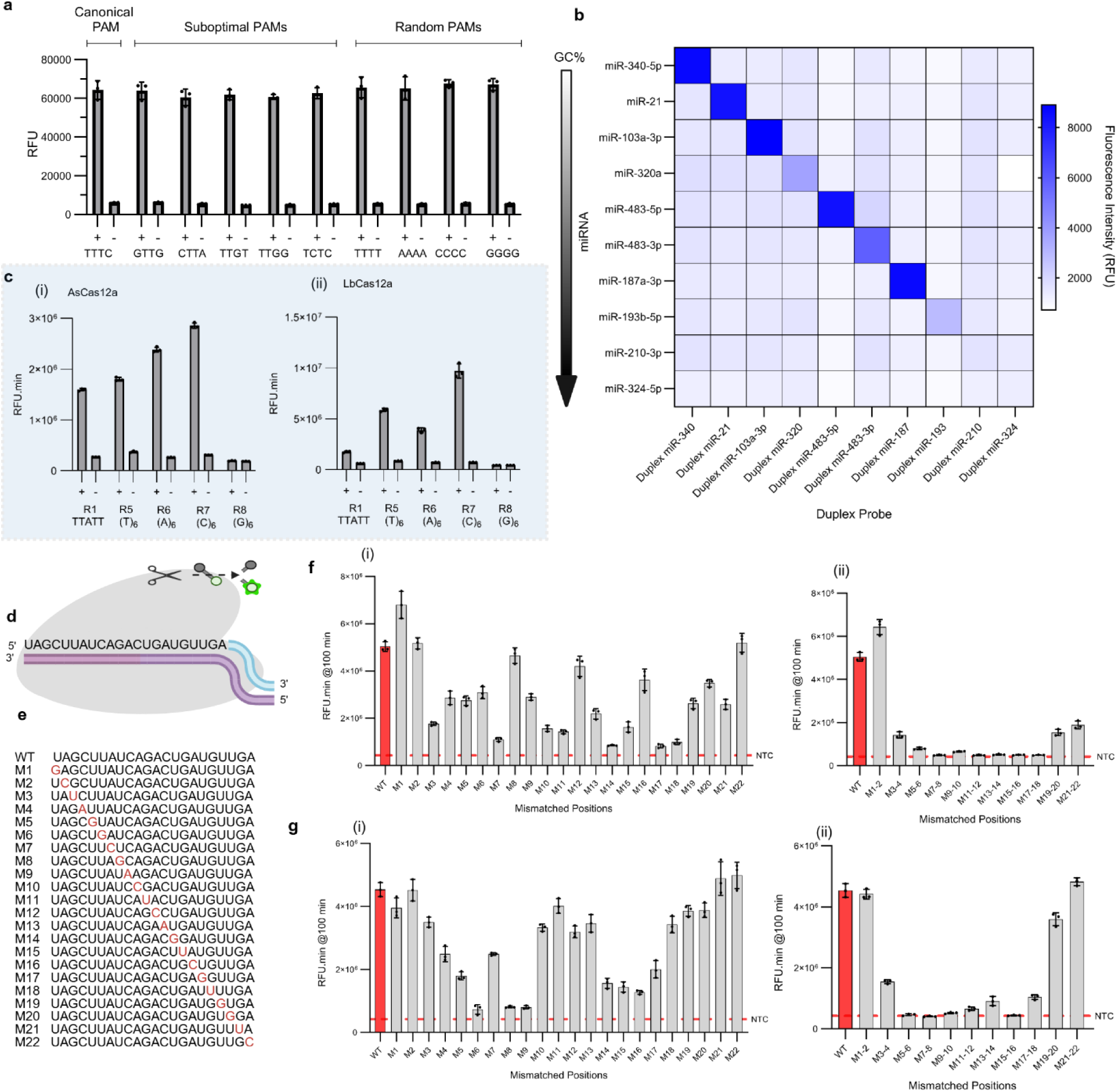
PAM, reporter, and mismatch specificity screening. (**a**) *Trans* cleavage profiles comparing canonical PAM, suboptimal PAMs, and random PAMs with mononucleotide repeats. **(b)** Orthogonality analysis of crRNA-free system across a panel of ten distinct miRNAs with varying GC content. The assay exhibits high specificity, generating fluorescence signal only for targets with ≤60% GC content. miR-210 and miR-324 (GC content >60%) fail to activate Cas12a *trans*-cleavage, consistent with GC-dependent performance constraints of the crRNA-free architecture. **(c)** Illustration of reporter *trans*-cleavage dynamics of crRNA-free Cas12a for: (i) AsCas12a, and (iv) LbCas12a. **(d)** Schematic illustration of single-nucleotide mismatch discrimination. **(e)** Wild-type miR-21 sequence aligned with single-nucleotide substitution mutants (M1–M22), where purine– pyrimidine mismatches are introduced sequentially across the sequence. **(f)** Fluorescence detection profiles of crRNA-free system against single-base (i) and paired two-base (ii) mismatches in miR-21. High discrimination is observed for several positions (e.g., M3–M7, M9–M11, M13–M15, M17–M19, and M21), while mismatches at terminal positions (e.g., M1–M2, M20–M22) show reduced selectivity. **(g)** Comparison of single-base (i) and two-base (ii) mismatches in DNA-21, the DNA analog of miR-21, tested using the same assay conditions. Mismatch tolerance patterns broadly mirror those of miR-21, though some divergence is noted at central and terminal mismatch positions. All data are presented as mean ± SD for n ≥ 3 technical replicates. All sequences can be found in **Tables 1-8**. (+) represents the present of target while (-) represents the absence of target.

### Sequence dependence, substrate preferences and specificity of crRNA-independent Cas12a activation

We next investigated the biochemical constraints governing crRNA-independent activation. Across a panel of short synthetic RNA oligonucleotides, including miRNA and arbitrary sequences (miniCaRs), each sequence activated Cas12a in sequence-specific manner with negligible cross-reactivity (**Fig. 2b**), despite the absence of conventional crRNAs. Notably, these short RNAs (∼22 nts) alone were sufficient to induce *trans* cleavage activity, indicating a crRNA-independent activation pathway that bypasses canonical R-loop formation. Activation exhibited a strong dependence on target sequence composition, with reduced activity observed for high-GC RNA sequences (≥60%), such as miR-210 and miR-324 (**Fig. 2b**). Systematic screening across an expanded panel of synthetic miRNAs revealed a consistent inverse relationship between GC content and activation efficiency, suggesting that stable secondary RNA structures or excessive hybrid stability may hinder productive engagement within the Cas12a nucleic acid-binding interface (**Suppl Fig. 6,7**). Notably, introduction of a cognate crRNA restored robust detection of high-GC targets (**Suppl Fig. 7; Extended data Fig. 2**), suggesting that reduced activity under crRNA-independent conditions can be alleviated by canonical crRNA loading.

Reporter substrate composition further modulated cleavage efficiency (**Fig. 2c, Suppl Fig. 8**). Among multiple ssDNA, RNA, and chimeric reporter designs, C-rich ssDNA sequences consistently produced the highest signal across eight Cas12a orthologs tested (**Extended Data Fig. 3**). Canonical crRNA-guided Cas12a systems, by comparison, typically use short T-rich ssDNA reporters, including sequences such as TTATT, because of their efficient cleavage kinetics.^11, 22, 26, 27^ G-rich reporters were essentially inactive, likely due to G-quadruplex formation or other inhibitory secondary structures that limit accessibility to *trans* cleavage.^28^ These results indicate that crRNA-independent activation not only alters initiation requirements but also reshapes downstream substrate preferences. Consistent with this, electrospray ionization mass spectrometry confirmed that fluorescence signals arise from bona fide reporter hydrolysis by Cas12a, yielding predominantly mononucleotide cleavage products across both canonical (TTATT) and C-rich reporters (**Suppl Fig. 9,10**). Furthermore, profiling *trans* cleavage activity across the Cas12a family, including AsCas12a, LbCas12a, FnCas12a, MbCas12a, Pb2Cas12a, ErCas12a, BoCas12a, and Lb5Cas12a (**Fig. 2c; Extended Data Fig. 3a-i**) indicates that collateral DNase activity is broadly conserved. In contrast, none of the Cas12a orthologs exhibited detectable cleavage of RNA-only reporters, reinforcing a strong intrinsic preference for ssDNA substrates (**Extended Data Fig. 3**). Together, these findings demonstrate that while crRNA-independent activation is conserved across Cas12a nucleases, its efficiency and substrate scope are modulated by ortholog-specific properties.

Despite these biochemical constraints, the system maintained high sequence specificity (**Fig. 2d-g**). To evaluate discrimination at single-nucleotide mismatch resolution for both DNA and RNA, we tested targets containing single-nucleotide polymorphisms (SNPs) across defined positions (**Fig. 2d-f(i),g(i)**). Introduction of a single mismatch markedly reduced *trans* cleavage activity in most cases, with the extent of attenuation depending on its position within the RNA–DNA heteroduplex. Mismatches located within the central region of the duplex, corresponding to the core of the crRNA-binding channel, produced the strongest loss of signal, whereas terminal mismatches were more frequently tolerated (**Fig. 2f,g**). This implies that crRNA-independent activation remains highly sensitive to local base-pairing integrity despite the absence of canonical crRNA-mediated recognition. These results underscore the ability of Cas12a to discriminate closely related sequences with single-nucleotide resolution under crRNA-independent activation.

Extending this analysis, we evaluated DNA and RNA targets containing two adjacent mismatches across all positions (**Table 12**). In all internal positions, paired mismatches reduced signal to near background levels, with only terminal mismatches (positions 1–2 and 21–22) showing partial tolerance (**Fig. 2f(ii), g(ii)**). This enhanced sensitivity to local base-pairing disruptions indicates that crRNA-independent activation supports stringent sequence discrimination at sub-seed resolution.

### miniCaRs engage the canonical crRNA–binding channel

To understand the molecular basis of crRNA-independent activation, we investigated how miniCaRs interact with Cas12a. Computational modeling using Boltz-2^29^ revealed that, in the absence of a crRNA, short RNAs can stably occupy the canonical crRNA-binding pocket of Cas12a (**Suppl Fig. 11**). In this configuration, the RNA is predicted to adopt a spatial arrangement that closely overlaps with the position of the crRNA scaffold and spacer in canonical complexes.

Fluorescence polarization and electrophoretic mobility shift assay (EMSA) analyses confirmed direct and high-affinity interactions between Cas12a, dAsCas12a, and fluorescently labeled miniCaRs, with nanomolar dissociation constants (**Fig. 3**). Importantly, miniCaR binding was competitively displaced by canonical crRNA, indicating that miniCaRs and crRNA occupy overlapping binding sites (**Fig. 3a(i)**). In addition, high-GC percentage miR-210–FAM and FAM-duplex DNA exhibited detectable, albeit weaker, binding affinities with K_D_ values of approximately 460 nM and 56 nM, respectively (**Fig. 3a(ii)**). Unexpectedly, miR-21–FAM could not be effectively displaced by excess unlabeled miR-21 (**Fig. 3a(iii)**), suggesting that oligonucleotide binding may be accompanied by a conformational change that stabilizes a tightly bound or partially caged state. Collectively, these data demonstrate that miniCaRs function as surrogate guides by engaging the native crRNA-binding channel of Cas12a. Similarly, EMSA was performed to assess binding of catalytically inactive AsCas12a (dAsCas12a) to fluorescently labeled nucleic acid substrates (**Fig. 3b**). Concentration-dependent dAsCas12a binding to miR-21 miniCaR substrate resulted in depletion of the lower labeled free-miniCaR species and accumulation of slower-migrating retarded material at higher protein concentrations (**Fig. 3b(i)**), and dAsCas12a binding to partial duplex DNA substrate resulted in progressive depletion of the unbound labeled duplex band and formation of slower-migrating protein–DNA complexes. Multiple protein-independent RNA bands were observed for miniCaR binding, consistent with distinct RNA conformations or species.^30^

**Fig. 3.**
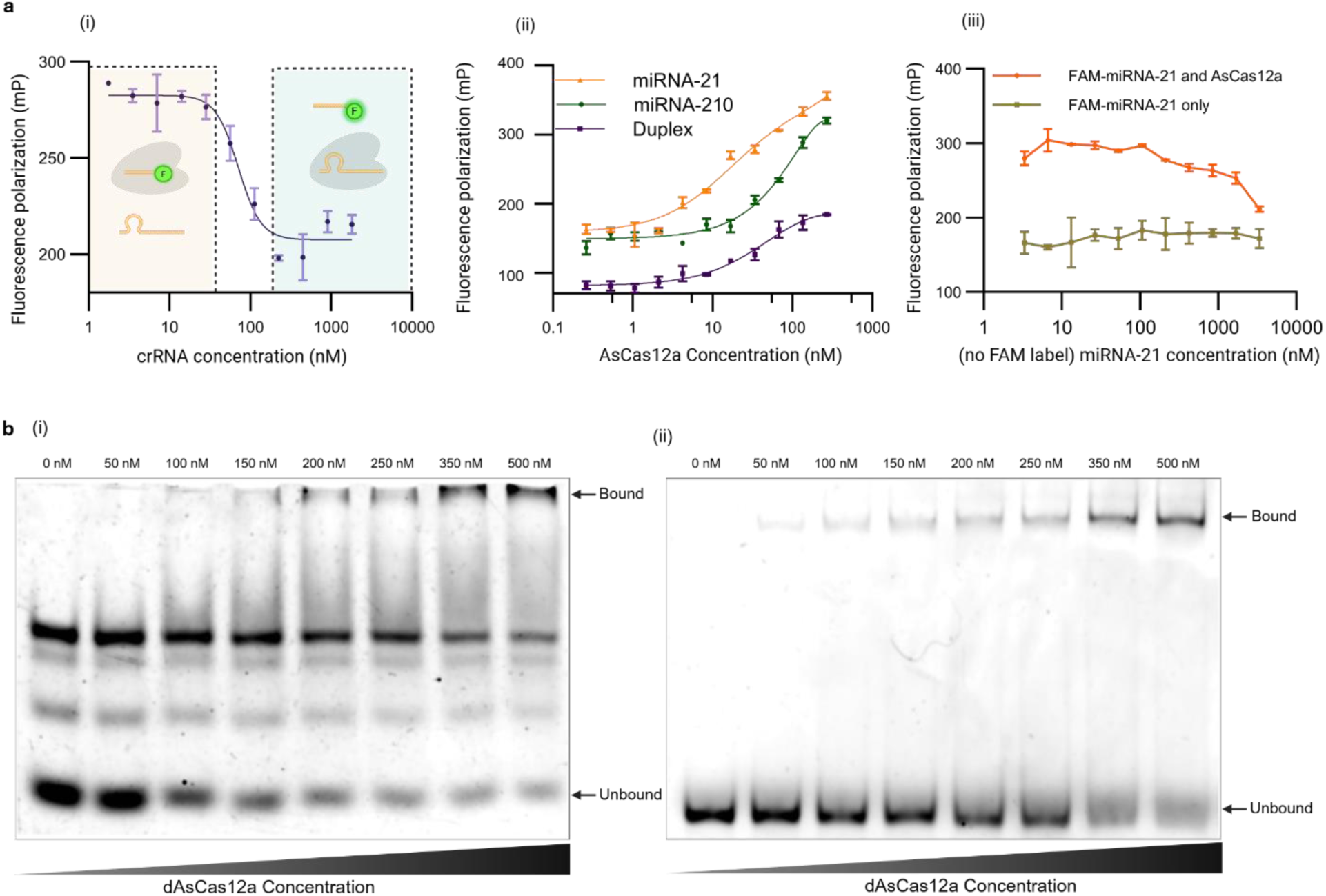
Binding properties of the crRNA-free Cas12a system. **(a)** Fluorescence-polarization binding assays with FAM-labeled miRNA. (i) Titration of the crRNA (purple) with the complex decreases polarization (EC_50_ ≈ 71 nM), suggesting displacement of FAM-miR-21 by the crRNA. (ii) The free FAM-miR-21 probe (orange trace) has low polarization, whereas addition of Cas12a produces a large increase in polarization (K_D_ ≈ 16 nM), indicating stable complex formation. By comparison, high GC miR-210 (green) and only duplex probe (plum) show weaker binding (K_D_ ≈ 460 nM and 56 nM, respectively). (iii) Titration of excess unlabeled miR-21 with the Cas12a:FAM-miR-21 complex (orange) and only FAM-miR-21 (gold) failed to reduce polarization, suggesting the bound FAM-miRNA is locked into the protein. These data confirm that Cas12a can directly engage short RNA within its native crRNA pocket. (**b**) Electrophoretic mobility shift assays (EMSA) demonstrating guide-independent binding of dAsCas12a to nucleic acid substrates. (i) Binding of dAsCas12a to a FAM-labeled 22 nt miR-21 miniCaR substrate resulted in depletion of the lower free-RNA species and accumulation of slower-migrating retarded material at higher protein concentrations. Multiple protein-independent RNA bands were observed in the absence of protein, consistent with distinct RNA conformations or species. (ii) Binding of dAsCas12a to a FAM-labeled partial DNA duplex substrate resulted in progressive depletion of the unbound duplex band and formation of slower-migrating protein– DNA complexes. Bound and unbound species are indicated by arrows. Samples were resolved on 6% native polyacrylamide gels in 0.5× TBE and imaged directly using FAM/fluorescein detection. All data are shown as mean ± SD for n ≥ 3 technical replicates.

### A crRNA-independent activated Cas12a state revealed by cryo-EM

To directly visualize the crRNA-independent activation state, we determined cryo-electron microscopy structures of AsCas12a bound to nucleic acid substrates in the absence of crRNA (**Fig. 4; Suppl Fig. 12,13**). The resulting structures reveal canonical bilobed Cas12a architecture,^21^ with REC and nuclease lobes preserved but significant rearrangements in regulatory regions (**Fig. 4b, c**).

**Fig. 4.**
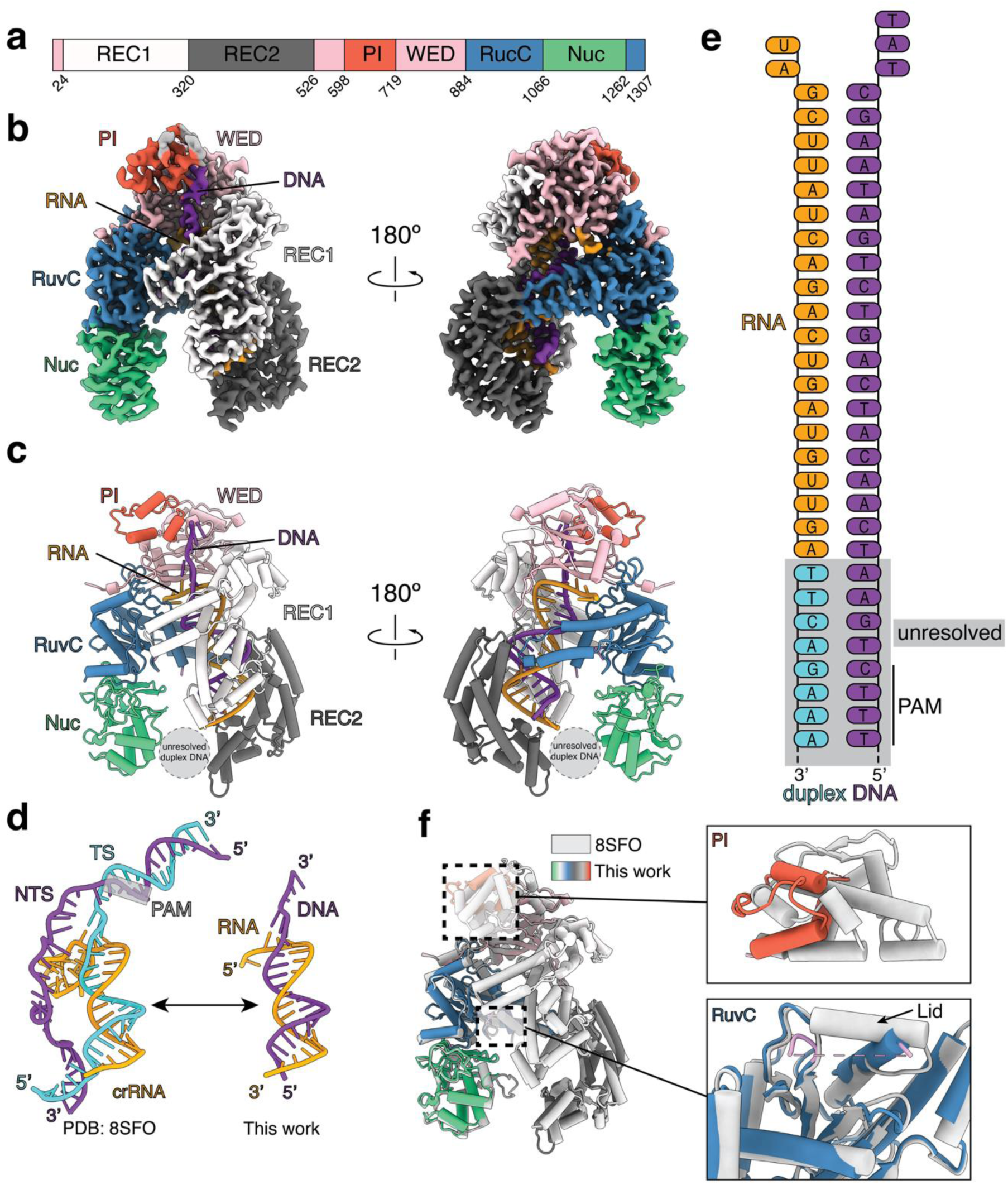
Cryo-EM structure reveals activation mechanism of crRNA-free Cas12a. (**a)** Domain organization of AsCas12a. REC1: light gray; REC2: dark gray; PI: tomato; WED: pink; RuvC: steel blue; Nuc: spring green. DNA: dark orchid; RNA: orange. (**b)** EMReady-processed cryo-EM map of Cas12a assembled with miniCaR and duplex. Same color codes were used as shown in panel **a**. (**c)** Structural model of complex shown in **b**. (**d)** Comparisons between the crRNA, dsDNA of 8SFO and the DNA-RNA heteroduplex of the crRNA-independent system. (**e)** Schematic diagram of DNA-RNA heteroduplex. The unresolved duplex DNA is under gray shadow and partially showed. (**f)** Structural superposition of 8SFO (gray) and the crRNA-independent structure. Insets show the zoom-in figures of the PI domain and RuvC lid. Dashed pink line denotes the unresolved RuvC lid in crRNA-independent system.

In the crRNA-free state, the short RNA and complementary DNA strand form a ∼20 bp heteroduplex that mimics the canonical crRNA–target DNA hybrid observed in active Cas12a complexes (**Fig. 4d, e**). Notably, the RNA–DNA hybrid is positioned within the canonical crRNA-binding channel, consistent with fluorescence polarization measurements indicating that short RNA engages the crRNA-binding interface. However, unlike canonical PAM-stabilized assemblies, the PI domain is poorly ordered, reflecting loss of stable PAM engagement (**Fig. 4f**). Consistent with this, the PAM-containing duplex DNA is not resolved in its canonical binding site and instead localizes to the base of the complex between the Nuc and REC2 domains (**Fig. 4c**), adopting an inverted orientation compared to the canonical AsCas12a ternary complex structure (8SFO^21^).

At the same time, the RuvC lid region exhibits pronounced flexibility, and is not resolved as observed by the absence of map density, in contrast to the ordered loop and helical conformations observed in canonical AsCas12a structures that correspond to inactive and active states, respectively. This suggests that the RuvC lid remains open, exposing the catalytic residues of AsCas12a. Together, these observations suggest that crRNA engagement and PAM-recognition are not strictly required for catalytic activation when a surrogate nucleic acid occupies the spacer-target DNA binding channel. Despite these differences, the global catalytic architecture remains intact, and the complex adopts a catalytically competent configuration capable of supporting *trans* cleavage activity.

We also aimed to compare our crRNA-independent system to our previously characterized RAPID platform, which incorporates conventional crRNA-mediated targeting. Two new RNA target-bound structures of RAPID reveal that addition of a crRNA^18^ reverts Cas12a to a more canonical mechanism, including completion of the bridge helix, continuous density for the ∼20-bp hybrid duplex, and clear ordering of REC2, consistent with full PAM-distal engagement (**Extended Data Fig. 4; Suppl Fig. 14**).

Together, these structural data reveal that Cas12a can access an alternative activation landscape in which crRNA-independent nucleic acids induce an active-like state through noncanonical RNA–DNA hybrid formation within the crRNA channel, coupled to inverted PAM-proximal DNA positioning and partial relaxation of regulatory gating elements.

### Amplification-free and amplification-assisted nucleic acid detection enabled by crRNA-independent Cas12a activation

The discovery of crRNA-independent Cas12a activation enables new diagnostic modalities that do not require engineered crRNAs, potentially improving deployment without a requirement for labile RNA components. Leveraging this mechanism, we developed a nucleic-acid detection platform in which minimal activating nucleic acids directly trigger Cas12a-mediated reporter cleavage, enabling sensitive readout without amplification. This system, which we call COMPANION (CRISPR-Cas12 Omnidirectional Molecular Probe Activation for Nucleic acid Identification ON demand), supports detection of both RNA and DNA targets through flexible probe architectures and operates across multiple Cas12a orthologs (**Fig. 1d**, **Fig. 5**). It enables sensitive, sequence-specific detection of miRNAs from low picomolar down to high femtomolar concentrations without pre-amplification (**Suppl Fig. 15**).

**Fig. 5.**
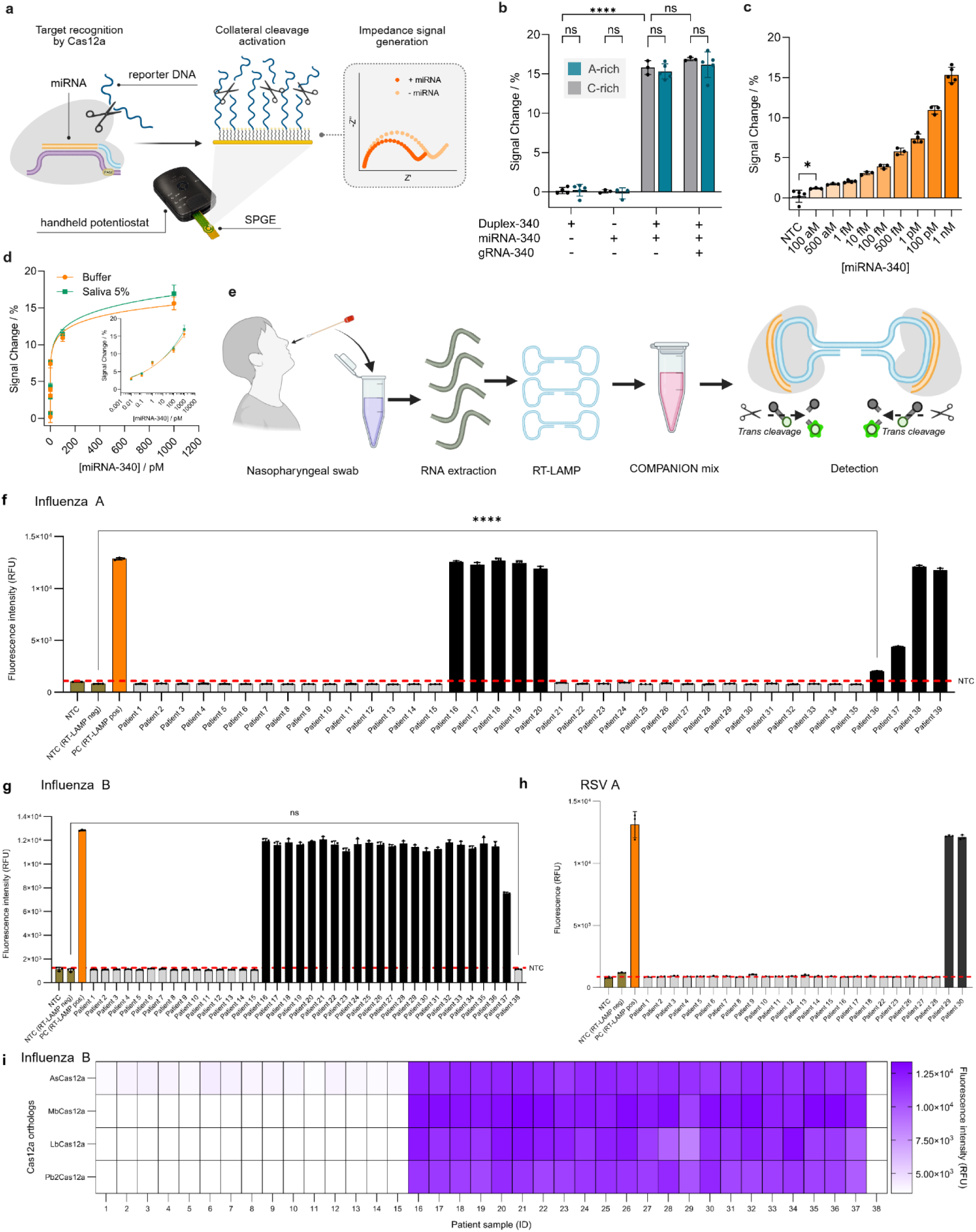
Diagnostic performance of the crRNA-free Cas12a platform COMPANION. **(a)** Electrochemistry workflow of the crRNA-free system, otherwise known as eCOMPANION. Here, Cas12a is activated by Cas12a–miniCaR-duplex complex, catalysing collateral cleavage of ssDNA reporters immobilized on gold electrode, resulting in a decrease in charge-transfer resistance (Rct), measured by electrochemical impedance spectroscopy. **(b)** Percentage reduction in Rct as a function of different nucleic acid input with A-rich and C-rich reporters. **(c)** Percentage reduction in Rct due to different miR-340 concentrations ranging from 100 aM to 1 nM. **(d)** Biosensor responses to spiked miRNA-340 (10 fM – 1 nM) measured in buffer and in 5% human saliva. **(e)** Schematics of the workflow for detecting respiratory viruses from nasopharyngeal swab samples using the COMPANION-RT-LAMP assay. After sample collection and RNA extraction, viral RNA is amplified by one-step RT-LAMP, and the amplicons are then detected by the COMPANION Cas12a-based trans-cleavage system. **(f)** Detection of Influenza A (H1N1) in 39 patient samples (9 true positive and 30 true negative cases). All 9 Influenza A-positive samples were correctly identified by COMPANION-RT-LAMP, with no false positives; appropriate positive and negative controls are included for comparison. **(g)** Detection of Influenza B in 38 patient samples (22 true positives, 15 true negatives, and 1 false negative – **Suppl** Fig. 23 for sample #38). One low-titer Influenza B sample was not detected by the assay (Sample #38), while all other positive samples were correctly identified. **(h)** Detection of RSV A in 25 patient samples (2 true positives and 23 true negatives). Both RSV-positive samples were correctly detected, and no false responses were observed. **(i)** Comparison of four Cas12a orthologs (AsCas12a, MbCas12a, LbCas12a, and Pb2Cas12a) for detecting Influenza B in patient samples. A heatmap (purple = target detected, white = no signal) summarizes results for samples 1–38. Samples 1–15 were true negatives (all remained white), and samples 16–38 were true positives (purple), except that sample 38 yielded a false-negative signal. All four Cas12a orthologs demonstrated high accuracy (≥ 97%) in identifying Influenza B positive and negative samples. All assays were performed with a 30 min COMPANION-RT-LAMP reaction at 37 °C. Data are shown as mean ± SD (n = 3 technical replicates). Black bars are positive sample. Grey bars are negative samples. NTC, no-template control; Neg, negative; Pos, positive. Statistical significance was analyzed using a two-tailed t test: where ns = not significant with ρ > 0.05, and the asterisk (*) denotes significant differences with ρ < 0.05 and (****) denotes significant differences with ρ < 0.0001.

We extended the COMPANION platform to a portable label-free electrochemical format (eCOMPANION), where cleavage of a ssDNA reporter on a gold electrode by Cas12a upon target recognition decreases charge-transfer resistance (R_ct_) proportionally to target concentration (**Fig. 5a**).^31, 32^ The assay showed attomolar sensitivity and high specificity (**Fig. 1b,c; Extended Data Fig. 5a,b; Suppl Fig. 16**). Four representative Cas12a orthologs (AsCas12a, MbCas12a, Pb2Cas12a, LbCas12a) enabled detection down to 1 fM, reaching ∼100 aM with AsCas12a and a C-rich reporter (**Extended Data Fig. 5c,d**). Across the orthologs, C-rich reporters outperformed A-rich sequences (**Extended Data Fig. 6c,d**). Interestingly, AsCas12a tolerated both C– and A–rich reporters and performed similarly with both, achieving ≥ 75-fold signal gain in comparison with NTC (**Fig. 5b; Extended Data Fig. 5b,c; Suppl Fig. 18**). Attomolar detection of multiple miRNAs was achieved without amplification, and performance was maintained in 5% saliva, demonstrating robustness for direct electronic detection in complex samples aiming towards point-of-care diagnostics (**Fig. 5d**).

Finally, integration with isothermal amplification (COMPANION-LAMP; **Suppl Fig. 18-23**; **Extended Data Fig. 6,7**) enabled direct detection of respiratory viral RNA from clinical nasopharyngeal swab samples. Across multiple patient cohorts, the system demonstrated high concordance with RT-qPCR, achieving ≥97–100% accuracy for Influenza A, Influenza B, and RSV A detection (**Fig. 5(e-i)**). These results demonstrate that crRNA-independent Cas12a activation can be harnessed for rapid, amplification-coupled diagnostics while retaining the mechanistic simplicity of direct nucleic acid targeting.

## Discussion

Our findings redefine how CRISPR–Cas12a nucleases can be engaged by nucleic acids by demonstrating that activation does not require a canonical crRNA or PAM-dependent target recognition. Instead, we show that short RNA species, which we term miniCaRs, can directly occupy the canonical crRNA–binding channel and induce a catalytically competent state with high sequence fidelity. Biochemical competition experiments indicate that miniCaRs and crRNAs bind mutually exclusively, consistent with shared occupancy of a conserved interface, while structural modeling and cryo-EM support a guide-like configuration of these RNAs within the binding cleft. Together, these observations establish that engagement of the crRNA-binding channel, rather than the full scaffold–spacer architecture, is sufficient to activate Cas12a.

This mechanism contrasts with the canonical multi-step activation pathway, in which crRNA loading, PAM recognition, and progressive R-loop formation coordinate conformational rearrangements that expose the RuvC catalytic site.^21, 22^ Our structural data indicate that miniCaR-mediated activation bypasses several of these steps while still stabilizing an active-like configuration. In this crRNA-free state, Cas12a retains its global catalytic architecture but exhibits altered organization of regulatory elements, including reduced ordering of the PI domain and increased flexibility of the RuvC lid. These features suggest that canonical PAM-dependent stabilization is not strictly required for activation, and instead that a more dynamic configuration of the nuclease can support *trans* cleavage activity. Consistent with this, the observed RNA– DNA hybrid adopts a noncanonical orientation within the complex, indicating that productive activation can be achieved through alternative nucleic acid geometries.

Functional analyses further support the existence of a distinct mode of nucleic acid engagement under crRNA-independent conditions. Activation is sensitive to target sequence composition, with high-GC RNAs failing to efficiently induce activity, likely due to stable secondary structures that limit productive binding within the crRNA channel. In parallel, cleavage assays reveal a pronounced preference for C-rich DNA reporters, suggesting that substrate positioning or the catalytic environment of the RuvC domain is altered in this state. Together, these findings indicate that miniCaR-mediated activation not only bypasses canonical recognition requirements but also reprograms aspects of substrate engagement and cleavage behavior. Importantly, this mechanism is conserved across multiple Cas12a orthologs, indicating that crRNA-independent activation reflects an intrinsic property of type V nucleases rather than a feature of a single enzyme variant. This conservation suggests that the crRNA–binding channel is inherently adaptable, capable of accommodating a broader range of nucleic acid ligands than previously appreciated. In this context, the canonical crRNA can be viewed as one optimized solution within a larger functional space of nucleic acid-mediated activation.

The ability of Cas12a to respond to noncanonical nucleic acid inputs also raises important questions regarding how nuclease activation is regulated in cellular environments. Although canonical crRNA– and PAM-dependent target recognition is thought to restrict activation to foreign nucleic acids, our findings suggest that the crRNA-binding channel may possess broader ligand tolerance than previously appreciated.

Determining whether endogenous nucleic acids can modulate Cas12a activity in native or engineered systems will therefore be an important direction for future investigation. These considerations may also have implications for therapeutic applications, where cellular RNAs could influence Cas12 activity in complex eukaryotic environments.

In summary, our results demonstrate that Cas12a can be activated through direct engagement of its crRNA-binding channel by minimal nucleic acid inputs, revealing an alternative activation geometry that bypasses canonical crRNA and PAM requirements. These findings also expand current models of CRISPR effector function by showing that nuclease activation is governed not by a single defined pathway, but by access to a broader conformational landscape. This mechanistic flexibility provides a foundation for rethinking how CRISPR nucleases can be programmed and suggests new opportunities for both fundamental studies and applied technologies.

## Acknowledgement

I.A.I. was supported by the Precision Medicine Initiative (PRiME) and Canadian Institute of Health Research (CIHR) with internal fellowship numbers PRMUHT2024-001 and 202410MFE-531769-419793, respectively, including the Provost’s Postdoctoral Fellowship Program at the University of Toronto, Canada. S.J.R.d.S received a Research Award (473621) funded by the Emerging & Pandemic Infections Consortium (EPIC), University of Toronto. D.W.J was supported by The São Paulo Research Foundation (FAPESP) with a Doctoral Fellowship (Grant No. 2021/09706-0) and the Research Abroad Scholarship (BEPE – Grant No. 2024/07237-0). This work was also supported by funds to K.P from the CIHR Foundation Grant Program (201610FDN-375469); the Canada Research Chairs Program (Files 950-231075 and 950-233107); the NSERC Discovery Grants Program (RGPIN-2016-06352). B.P.H was supported by a Project Grant from the CIHR (PJT-189974) and an NSERC Discovery Grant (RGPIN-2023-04238). R.C was supported by a CGS-M award. D.H was supported by a Postdoctoral Pathway Fellowship. Furthermore, this work was also developed with funding from the Defense Advanced Research Projects Agency (DARPA), Contract No. N66001-23-2-4042 to A.A.G and K.P. The views, opinions and/or findings expressed are those of the authors and should not be interpreted as representing the official views or policies of the Department of Defense or the U.S. Government. We thank Axel Brilot (Sauer Structural Biology Laboratory, University of Texas at Austin) for cryo-EM assistance. We also thank Robert Flick, Mass Spectrometry and Metabolomics Services Manager in the Department of Chemical Engineering and Applied Chemistry at the University of Toronto, for his assistance with the mass spectrometry experiments. I.A.I thanks Abasi-ifreke Karena Idorenyin, Zachary Idorenyin Iwe, Ariel Idorenyin Iwe, and Deborah Idorenyin Iwe for their understanding and support during this work.

## Approval for Public Release

This work was cleared by DARPA and approved for public release, distribution unlimited. Further questions should be channeled to DARPA via their Public Release Center located at 675 N. Randolph Street, Room 03-028, Arlington, VA 22203-1714.

## Author contributions

I.A.I., S.S., K.G., and R.F.O. contributed equally. I.A.I. and N.E. conceived the ideas. K.P., I.A.I., S.S., S.J.R.d.S., D.W.J., K.G., R.F.O., N.E., L.W., D.H., P.K., R.C., B.P.H., A.C., K.B., G.A.W., and D.W.T designed, performed, and analysed the experiments. I.A.I. coordinated and supervised the experimental work. I.A.I, S.S., A.C., S.J.R.d.S., D.W.J., K.G., R.F.O., K.P., I.Z., and P.K. wrote the manuscript. S.J.R.d.S. designed and handled all clinical trials. D.W.J. designed and performed all the electrochemistry experiments. I.Z., K.G., and R.F.O. performed all the computational and structural studies. K.G., D.W.T., and R.F.O. performed all cryo-EM experiments. T.M. provided all the patient samples and assisted with results interpretation. F.X.L., A.A.G., S.Y., and Z.L. provided useful insights to shaping the work. I.Z. handled Cas computational structural analysis. K.P., I.A.I., S.S., S.J.R.d.S., D.W.J., K.G., R.F.O., N.E., R.Z., I.Z., R.C., D.H., B.P.H., A.C., K.B., J.N., P.B., M.Z., F.X.L., Su.S. A.R., A.A.G., Z.L., and S.Y. discussed the results, revised or commented on the manuscript. K.P. edited the manuscript and supervised all aspects of the work.

## Data availability

Cryo-EM map of COMPANION was deposited to the Electron Microscopy Data Bank (EMDB) under accession number EMD-76860. Associated atomic coordinate was deposited to the Protein Data Bank (PDB) under accession number 12YR. Cryo-EM maps of RAPID (5bp target RNA) and RAPID (full 20bp target RNA) were deposited to the Electron Microscopy Data Bank (EMDB) under accession number EMD-76961 and EMD-76960, respectively. Associated atomic coordinates were deposited to the Protein Data Bank (PDB) under accession number 13CF and 13CE, respectively. The data that support the plots within this paper and other findings of this study are available from the corresponding authors upon reasonable request.

## Supplementary information

**Extended Data** Figs. 1-7, **Supplementary** Figs. S1-23, and **Supplementary** Tables S1-18

