## Extended Data File for "CRISPR RNA-independent activation of Cas12a"

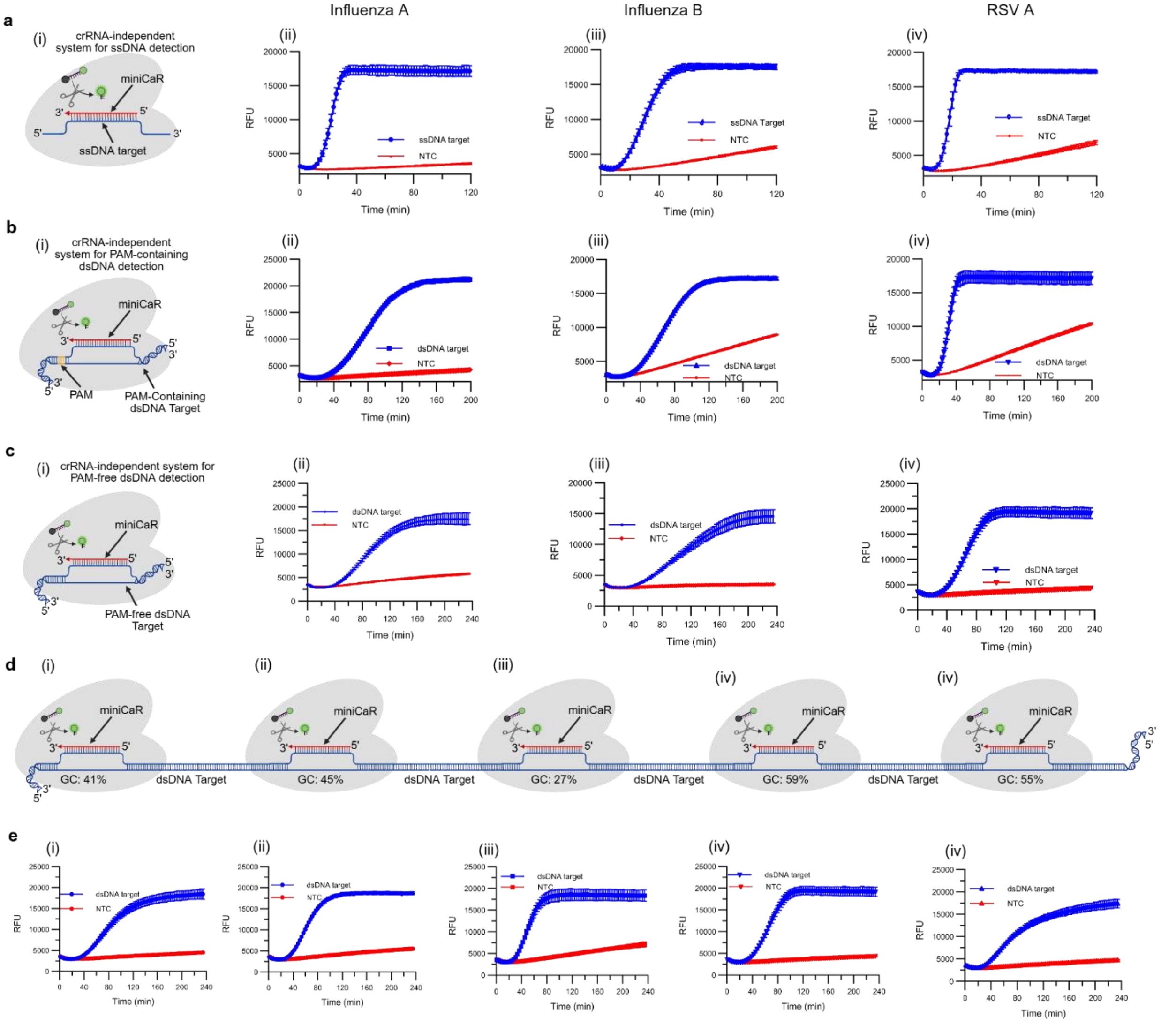

**Extended Data Fig. 1. crRNA-independent detection of ssDNA and dsDNA targets across respiratory viruses.** (**a**) PAM-independent ssDNA detection of three representative viral targets: Influenza A (i), Influenza B (ii), and RSV A (iii), using miniCaR 5, miniCaR 3, and miniCaR 4, respectively. (**b**) crRNA-independent identification of PAM-containing dsDNA targets using the same miniCaRs as in (a). (**c**) PAM-independent dsDNA detection in a crRNA-free format using miniCaR 2 (Influenza A), miniCaR 1 (Influenza B), and miniCaR 6 (RSV A). (**d**) Long RSV A **dsDNA** substrates using distinct miniCaRs binding at loci with varying GC content: miniCaR 1 (41%) (i), miniCaR 2 (45%) (ii), miniCaR 4 (27%) (iii), miniCaR 5 (59%) (iv), and miniCaR 6 (59%) (v). (**e**) Kinetic profiles corresponding to the binding regions shown in (**d**). DNA templates and all miniCaR sequences are provided in **Supplementary Table 9**. For panels (a–c), (i–iii) correspond to Influenza A, Influenza B, and RSV A, respectively.

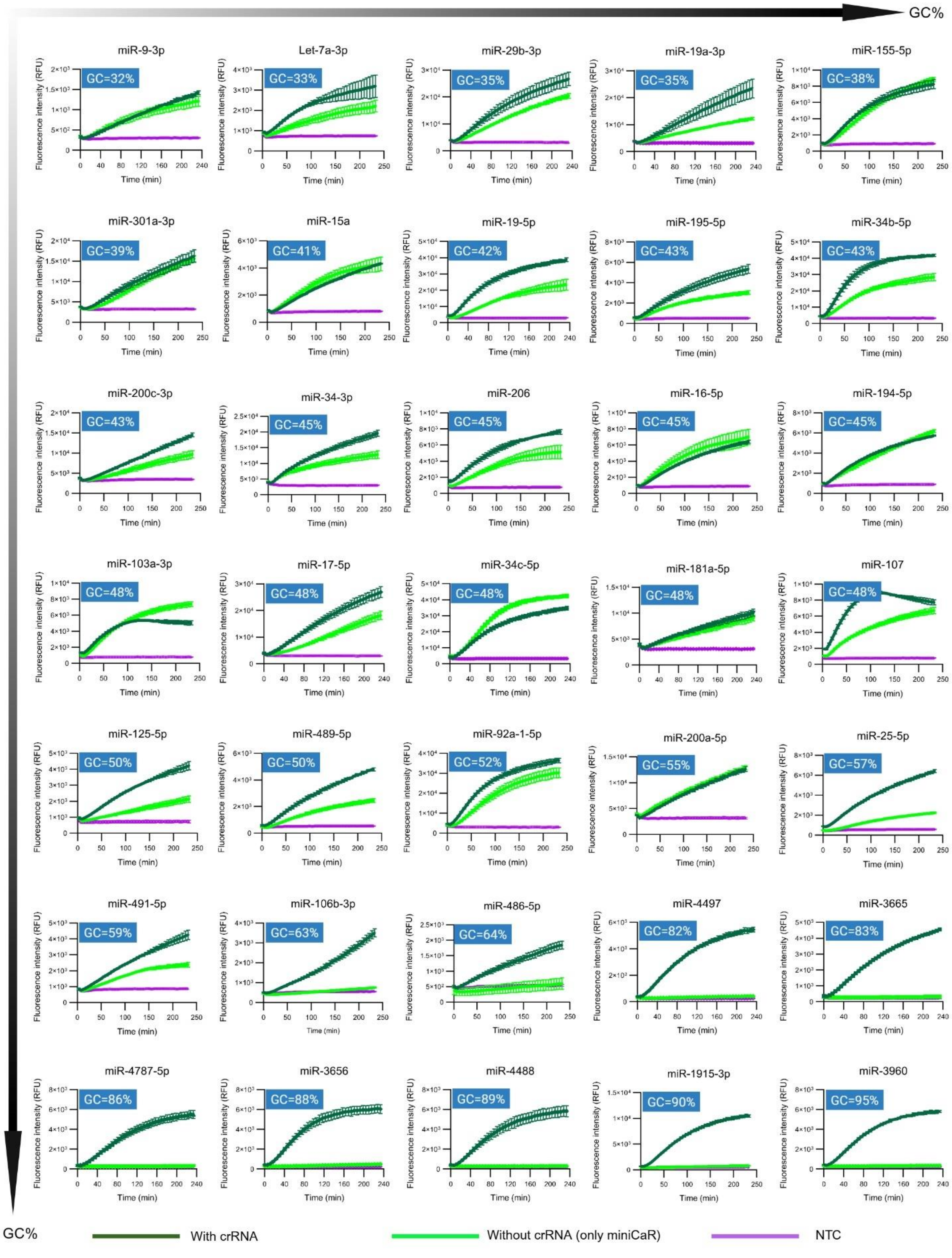
GC%
miR-9-3p
GC=32%
Let-7a-3p
GC=33%
miR-29b-3p
GC=35%
miR-19a-3p
GC=35%
miR-155-5p
GC=38%
miR-301a-3p
GC=39%
miR-15a
GC=41%
miR-19-5p
GC=42%
miR-195-5p
GC=43%
miR-34b-5p
GC=43%
miR-200c-3p
GC=43%
miR-34-3p
GC=45%
miR-206
GC=45%
miR-16-5p
GC=45%
miR-194-5p
GC=45%
miR-103a-3p
GC=48%
miR-17-5p
GC=48%
miR-34c-5p
GC=48%
miR-181a-5p
GC=48%
miR-107
GC=48%
miR-125-5p
GC=50%
miR-489-5p
GC=50%
miR-92a-1-5p
GC=52%
miR-200a-5p
GC=55%
miR-25-5p
GC=57%
miR-491-5p
GC=59%
miR-106b-3p
GC=63%
miR-486-5p
GC=64%
miR-4497
GC=82%
miR-3665
GC=83%
miR-4787-5p
GC=86%
miR-3656
GC=88%
miR-4488
GC=89%
miR-1915-3p
GC=90%
miR-3960
GC=95%
Fluorescence intensity (RFU)
Time (min)
GC%
With crRNA
Without crRNA (only miniCaR)
NTC

**Extended data Fig. 2.** *Trans* cleavage kinetics for an additional 35 miRNAs (or miniCaRs) with varying GC content (32%–95%). Dark green curve indicates the presence of crRNA, while light green shows crRNA-independent system, and purple curve depicts the blank or no-target control (NTC). Downward- and rightward-pointing arrows indicate increasing GC content across the data. miRNAs or miniCaRs with GC ≤60% produce strong signals (some comparable to the conventional crRNA-assisted system), whereas miRNAs with GC ≥60% fail to activate *trans* cleavage. This confirms that the crRNA-independent system effectively activates Cas12a for low to moderate GC targets but not for high GC-rich targets. All sequences and extended data are provided in **Suppl Tables S1,3,4**. All data are mean ± SD (n ≥ 3 technical replicates). Assays were performed with AsCas12a and reporter R7. The reduced activity on GC-rich miRNAs is consistent with reports that high-GC RNA targets form stable secondary structures, lowering hybridization efficiency and fluorescence signal.[1]

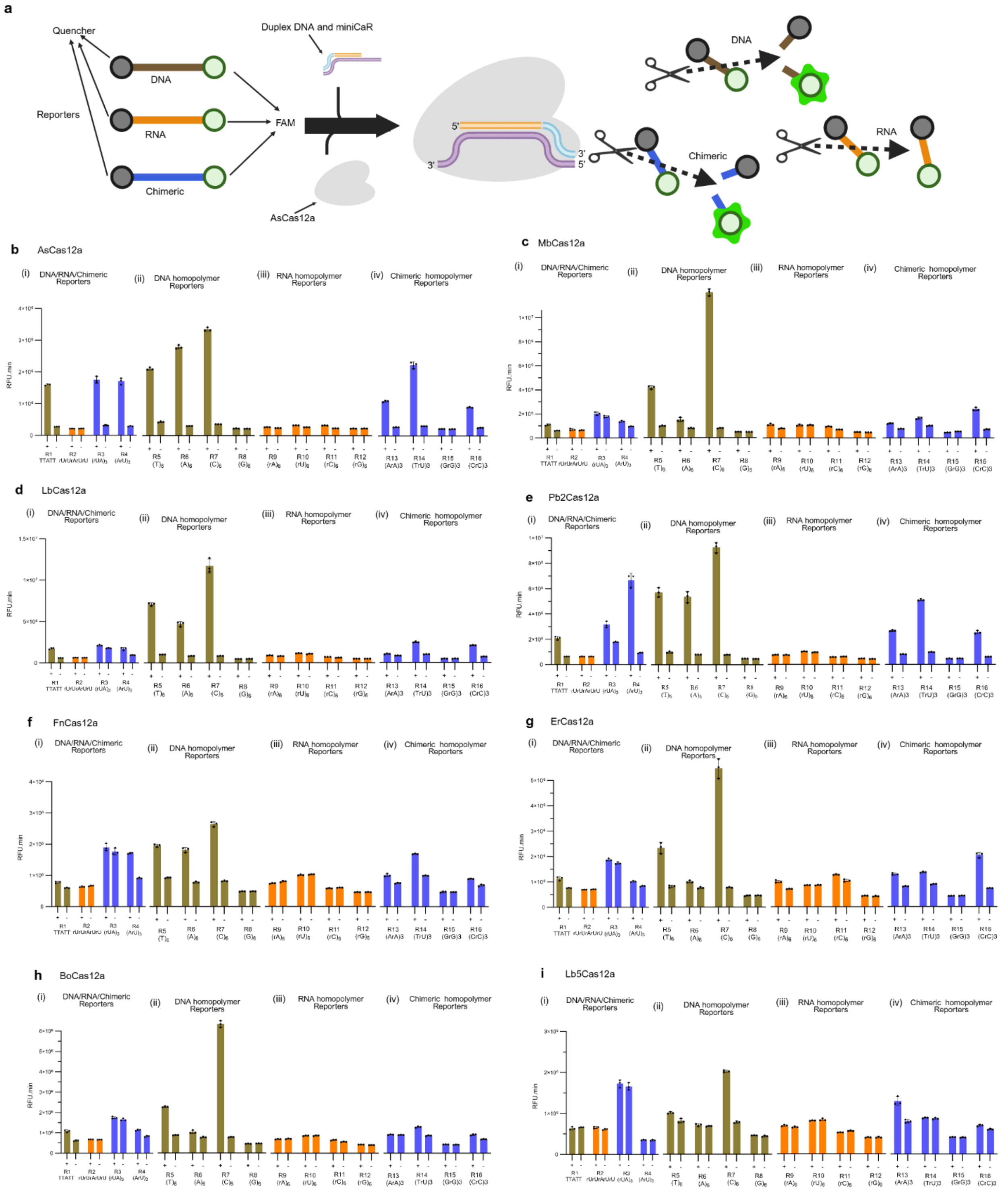

**Extended Data Fig. 3.** Reporter design and Cas12a ortholog compatibility in crRNA-independent system. (**a**) Schematic of the fluorophore–quencher reporter used to assay Cas12a *trans* cleavage on DNA, RNA, and chimeric substrates (i-iii). *Trans* cleavage performance for 16 different reporters (various DNA/RNA/chimeric sequences) tested across eight Cas12a orthologs: AsCas12a (b), MbCas12a (c), LbCas12a (d), Pb2Cas12a (e), FnCas12a (f), ErCas12a (g), BoCas12a (h), and Lb5Cas12a (i). Reporter sequence composition strongly influenced activity. All orthologs cleaved the C-rich reporter (R7: $C_6$)

efficiently, while the remaining reporters generally produced weak signals. Notably, AsCas12a robustly cleaved all DNA and chimeric reporters tested, in contrast to the other orthologs. No ortholog cleaved poly-G reporters. (Fluorescence time courses (RFU.min) over 60 min are shown to illustrate *trans* cleavage rates.) All data are mean ± SD (n ≥ 3 technical replicates). Color description: gold bars, DNA; orange bars, RNA; and blue bars, chimeric. All reporter sequences are provided in **Suppl Tables S9.** Sequences for DNA duplex, miniCaR, and crRNA for miR-21 are found in **Suppl Tables S1,3,4**.

**Extended Data Table 1**: Cryo-EM data collection, refinement, and validation statistics.

| | COMPANION | RAPID-5bp target RNA | RAPID-Full target RNA |
|---|---|---|---|
| **Data collection and processing** | | | |
| Magnification | 105,000 | 105,000 | 105,000 |
| Voltage (kV) | 300 | 300 | 300 |
| Electron exposure ($e^{-}/Å^{2}$) | 70 | 70 | 70 |
| Defocus range (μm) | –1.5 to –2.5 | –1.5 to –2.5 | –1.5 to –2.5 |
| Pixel size (Å) | 0.8332 | 0.8332 | 0.8332 |
| Initial particle (no.) | 921,805 | 3,235,710 | 3,235,710 |
| Final particle (no.) | 462,481 | 67,054 | 181,085 |
| Map resolution (Å) | 3.14 | 3.16 | 3 |
| FSC threshold | 0.143 | 0.143 | 0.143 |
| Refinement | | | |
| Initial model used | AlphaFold3 | 8SFO | 8SFO |
| Model resolution (Å) | 3.1 | 3.2 | 3 |
| FSC threshold | 0.143 | 0.143 | 0.143 |
| Map sharpening B factor ($Å^{2}$) | -168 | -99.8 | -107 |
| Model composition | | | |
| Non-hydrogen atoms | 10,241 | 9,317 | 9,636 |
| Protein residues | 1,143 | 1,058 | 1,242 |
| Nucleotides | 45 | 65 | 88 |
| **Mean B factors ($Å^{2}$)** | | | |
| Protein | 115.9 | 114.06 | 68.21 |
| Nucleotides | 83.73 | 107.22 | 51.6 |
| R.m.s deviations | | | |
| Bond lengths (Å) | 0.006 | 0.004 | 0.005 |
| Bond angles (°) | 1.173 | 0.822 | 0.817 |
| **Validation** | | | |
| MolProbity score | 1.44 | 1.36 | 1.35 |
| Clashscore | 6.91 | 4.03 | 4.76 |
| Poor rotamers (%) | 1.18 | 1.17 | 0.79 |
| Ramachandran plot | | | |
| Favored (%) | 98.05 | 97.47 | 97.42 |
| Allowed (%) | 1.95 | 2.53 | 2.58 |
| Disallowed (%) | 0 | 0 | 0 |
| Map CC (mask) | 0.78 | 0.78 | 0.82 |
| PDB | 12YR | 13CF | 13CE |
| EMDB | EMD-76860 | EMD-76961 | EMD-76960 |

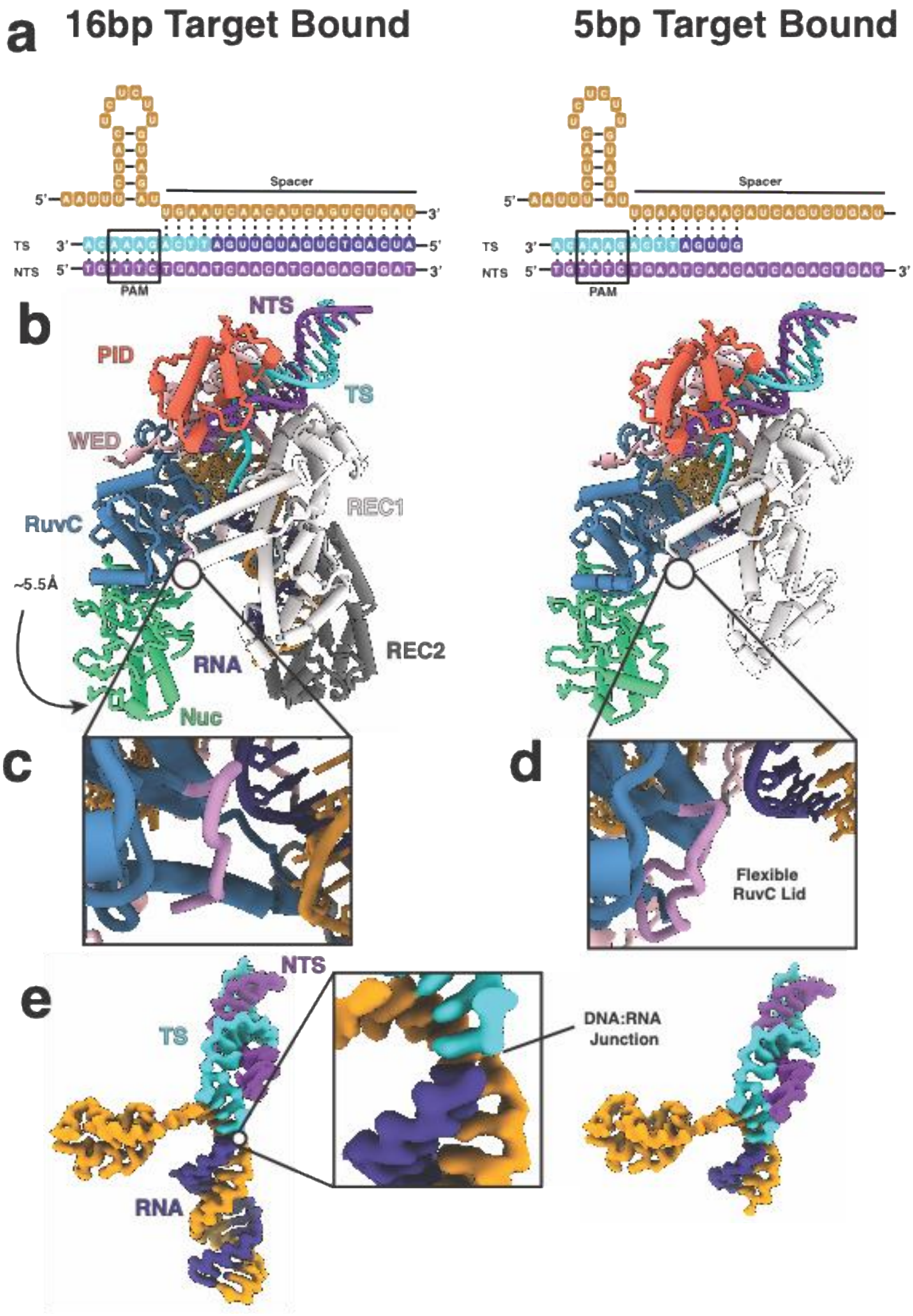
a
16bp Target Bound
5bp Target Bound
Spacer
TS
NTS
PAM
b
NTS
PID
TS
WED
RuvC
REC1
~5.5Å
RNA
REC2
Nuc
c
d
Flexible
RuvC Lid
e
NTS
TS
DNA:RNA
Junction
RNA

**Extended data Fig. 4.** Structural comparison of 5bp and 16bp target bound RAPID complex. **(a)** Nucleic acid schematic for (left) the 16 bp target bound structure and (right) 5bp target bound structure. **(b)** Models for (left) the 16 bp target bound structure and (right) 5bp target bound structure. The 16bp target bound structure closely resembles a 20bp structure of a fully matched R-loop (PDB: 8SFO) with a few exceptions including a non-resolved NTS and a flexible RuvC lid. The 5bp target bound structure closely resembles the intermediate state between an 8bp and 10bp structures previously reported (PDB: 8SFI and 8SFJ, respectively). (**c**) Direct visualization of the flexible RuvC lid in the 16bp complex and (**d**), the 5bp complex, indicating that the RuvC lid does not form in this process likely due to the increased flexibility of the NTS. (**e**) Visualization of the nucleic acid density within the cryo-EM map shows a clear break in density at the DNA:RNA junction for both the 16bp and 5bp target bound structure, which is known as RAPID.[2]

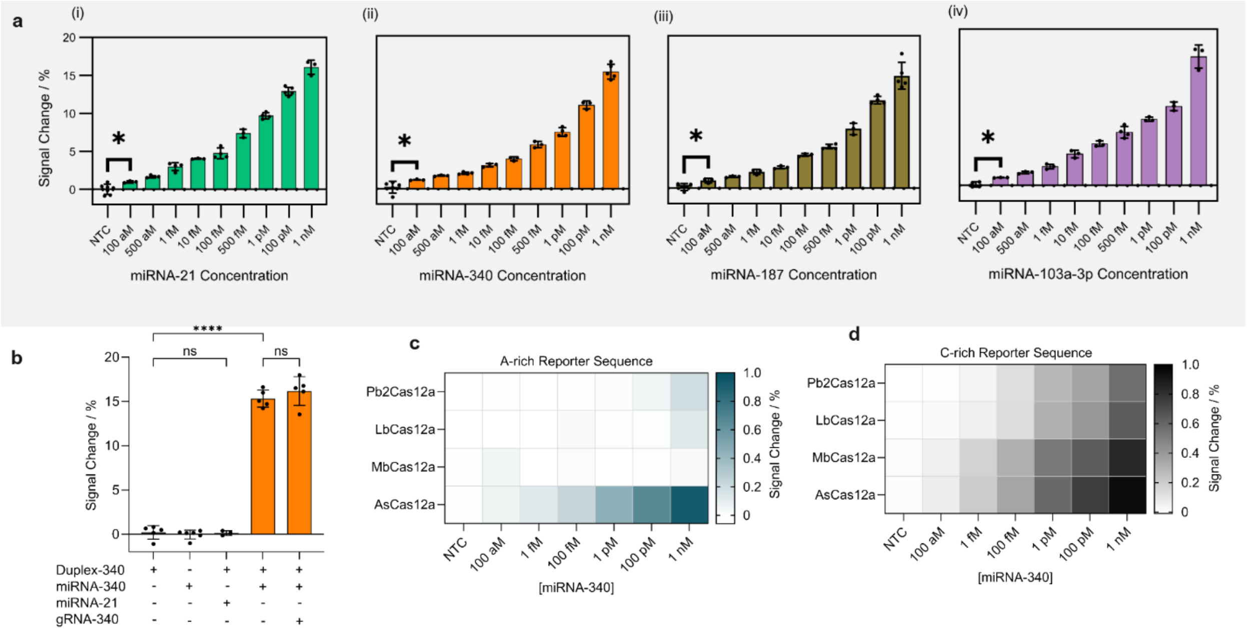

**Extended Data Fig. 5.** eCOMPANION biosensor for label-free and ultrasensitive miRNA detection. (**a**) Percentage of Rct reduction as a function of miRNA concentration (100 aM – 1 nM) for (i) miRNA-21; (ii) miRNA-340; (iii) miRNA-187, and miRNA-103a-3p. **(b)** Comparison of cleavage efficiency (signal change), calculated from the decrease in charge-transfer resistance ($\Delta Rct/Rct_0 \times 100\%$), indicating minimal background for NTC and non-complementary miRNA and similar performance for eCOMPANION and canonical-crRNA system. Effect of DNA reporter composition for A-rich reporter (**c**) and C-rich reporter (**d**) for representative Cas12a orthologs (AsCas12a, LbCas12a, MbCas12a and Pb2Cas12a) on *trans*-cleavage efficiency at different miRNA-340 concentrations, ranging from 100 aM to 1 nM. Data are shown as mean ± s.d. ($n \geq 3$ independent replicates). Statistical significance was analyzed using ANOVA test: where ns = not significant with $\rho > 0.05$, and the asterisk (*) denotes significant differences with $\rho < 0.05$ and (****) denotes significant differences with $\rho < 0.0001$. Sequences for DNA *trans*-cleavage templates are found in **Suppl Tables S15**.

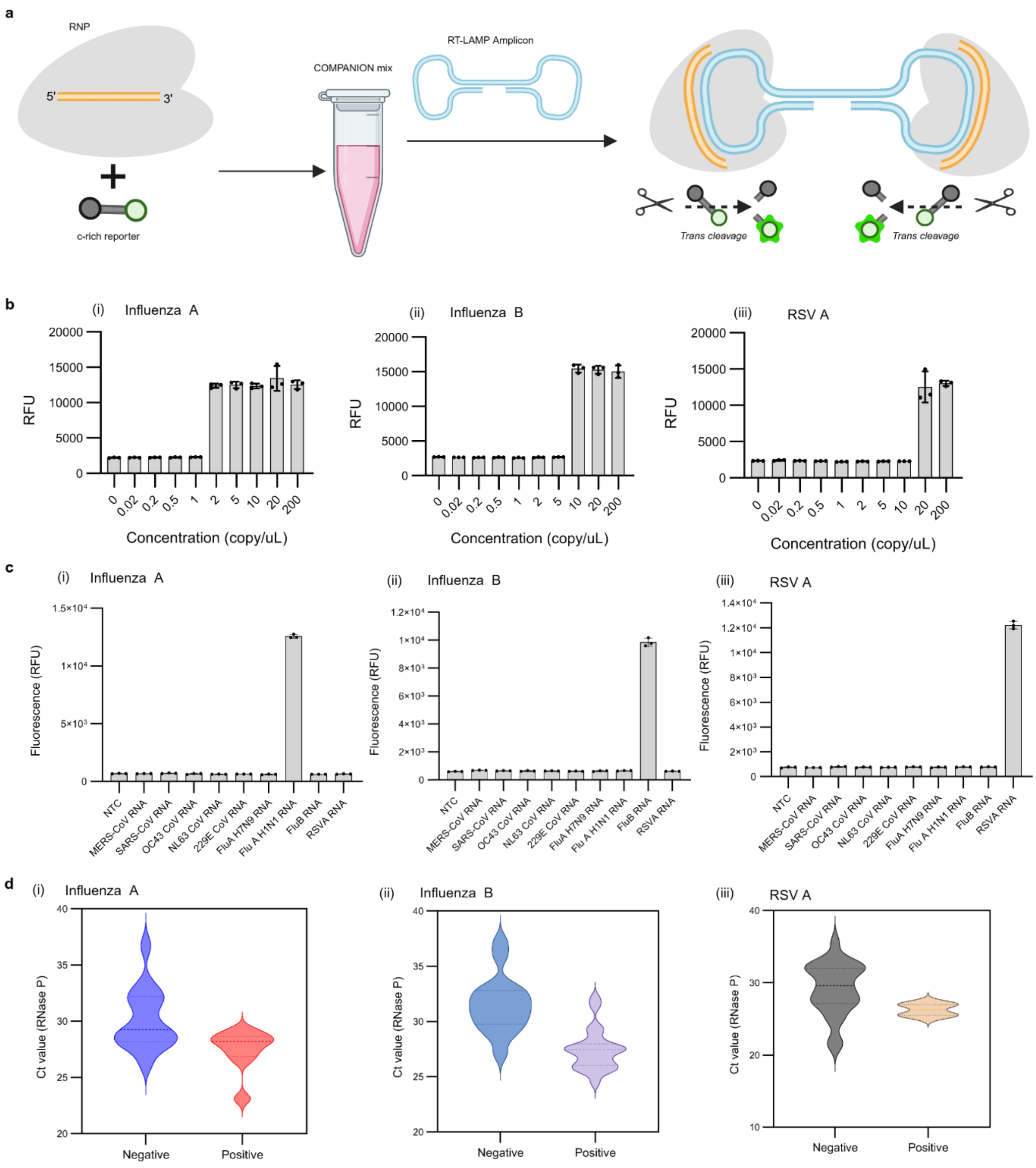

**Extended Data Fig. 6.** COMPANION-RT-LAMP detection of respiratory viruses. (**a**) Schematic illustration of the COMPANION mechanism. Here, the AsCas12a protein is combined with a short complementary RNA (miniCaR) that binds to the loop of a dumbbell-shaped DNA amplicon generated by LAMP. This binding activates Cas12a's *trans* cleavage of a FAM/quencher-labeled DNA reporter. (**b**) Limit of detection of the COMPANION-RT-LAMP assay for three target viruses: Influenza A (H1N1), Influenza B, and RSV A. The assay achieved a sensitivity of ~2 copies/µL for Influenza A (H1N1) **(i)**, ~10 copies/µL for Influenza B **(ii)**, and ~20 copies/µL for RSV A **(iii)**. (**c**) Specificity of the COMPANION-RT-LAMP platform for Influenza A, Influenza B, and RSV A. Each target was tested against six non-target respiratory viruses (MERS-CoV, SARS-CoV, HCoV-OC43, HCoV-NL63, HCoV-229E, and Influenza A (H7N1)). No detectable signal was generated for any non-target virus within 30 min, indicating high specificity. (**d**)

RNase P internal control results for patient nasopharyngeal samples tested for Influenza A (H1N1), Influenza B, or RSV A. RNase P (a human housekeeping gene) was successfully amplified in all samples, confirming adequate sample quality; the RNase P levels were similar in virus-positive and virus-negative samples. All results shown were obtained from 30 min COMPANION-RT-LAMP reactions at 37 °C. Data are presented as mean ± SD (n = 3 technical replicates). NTC, no-template control.

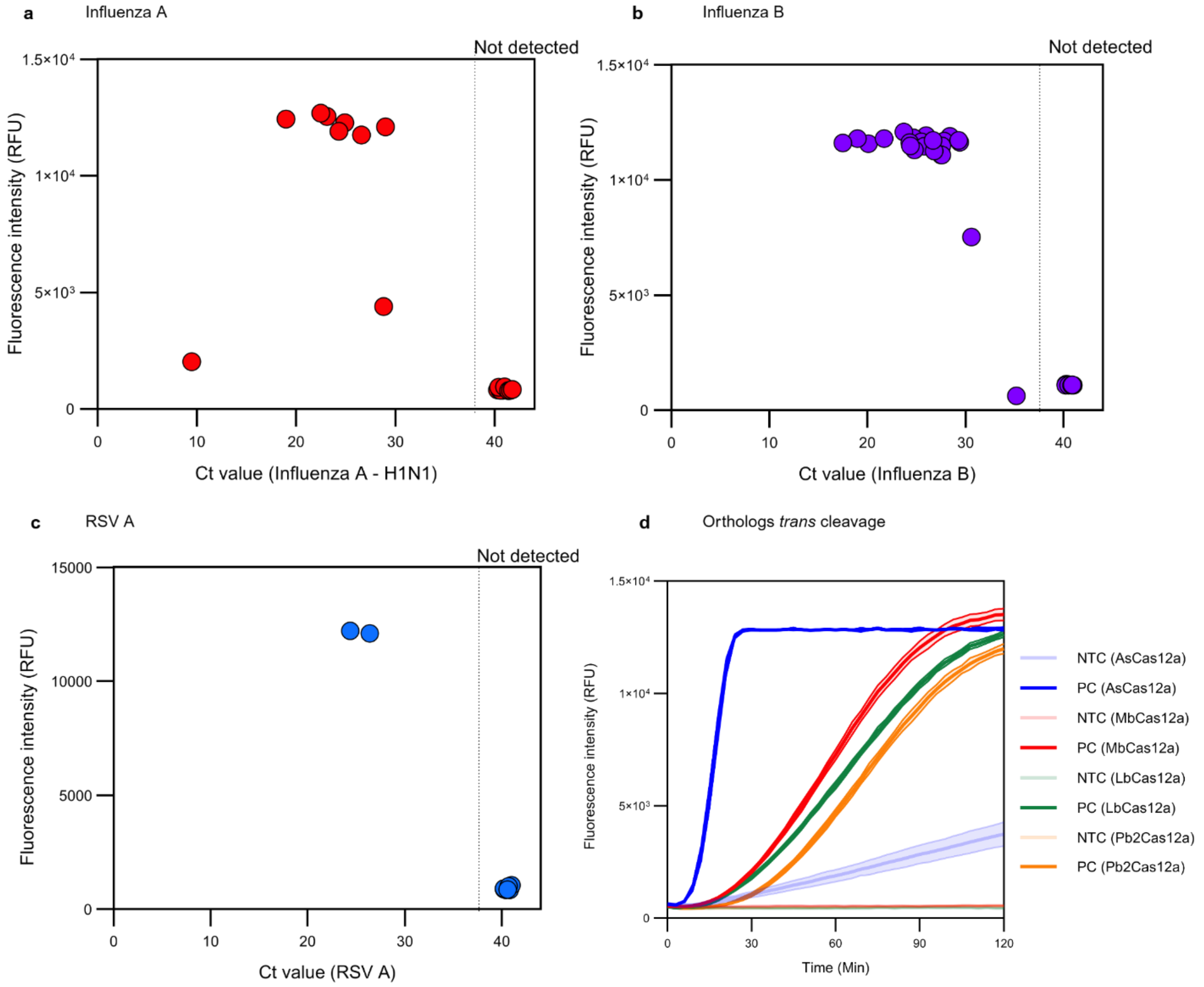

**Extended Data Fig. 7.** Correlation of COMPANION-RT-LAMP detection with qPCR Ct values, and Kinetic performance of different Cas12a orthologs. Relationship between COMPANION-RT-LAMP fluorescence signal and the RT–qPCR cycle threshold (Ct) of patient samples for Influenza A (**a**), Influenza B (**b**), and RSV A (**c**). Each point represents an individual patient sample. Samples with lower Ct values (higher viral loads) yield higher fluorescence signals with COMPANION-RT-LAMP, whereas samples that were negative by RT–qPCR (assigned Ct values ≥ 38) show no detectable fluorescence. This demonstrates that the COMPANION-RT-LAMP assay can reliably detect viral samples with Ct values up to the high 30s, and it produces no false signal for samples beyond the qPCR detection limit. (**d**) Real-time fluorescence kinetics of the COMPANION-RT-LAMP reaction for four different Cas12a orthologs (AsCas12a, MbCas12a, LbCas12a, and Pb2Cas12a) using Influenza B target. AsCas12a displays the fastest detection kinetics (fluorescence signal rises rapidly), whereas the other three orthologs produce fluorescence more slowly over the 2 h observation period. AsCas12a also exhibits a higher background signal in no-target control reactions, in contrast to MbCas12a, LbCas12a and Pb2Cas12a, which maintain a low, flat background fluorescence.

**Supplementary Information**

### CRISPR RNA-independent activation of Cas12a

Idorenyin A. Iwe[*1,2], Serena Singh[*1], Kaoling Guan[*3], Rodrigo Fregoso Ocampo[*3,4], Severino Jefferson Ribeiro da Silva[1], Dagwin Wachholz Junior[1,5,6], Nadia Emami[1], Ariel Corsano[1], Ilan Zeisler[1], Kristof Bozovicar[1], Liu Wang [1], Dalton Ham[7], Rita Cai[7], Paul Kelly[1], Riham Zayani[1], Jessica Nguyen[1], Pouriya Bayat[1], Moiz Charania[1], Sean Palter[1], Frank X. Liu[8,9], Suman Shrestha [1], Ashayad Rayhan[1], Gregory A. Wasney[10], Tony Mazzulli[11,12], Alexander A. Green[13,14,15], Zhigang Li[8], Shuhuai Yao[8], Basil P. Hubbard[1,7], David W. Taylor[**3,4,16,17], Keith Pardee[**1,2]

[1]Department of Pharmaceutical Sciences, Leslie Dan Faculty of Pharmacy, University of Toronto, Toronto, ON M5S 3M2, Canada.

[2]Department of Mechanical and Industrial Engineering, University of Toronto, Toronto, ON M5S 1A1, Canada.

[3]Department of Molecular Biosciences, University of Texas at Austin, Austin, TX, USA

[4]Interdisciplinary Life Sciences Graduate Programs, University of Texas at Austin, Austin, TX, USA.

[5]Institute of Chemistry, University of Campinas, Campinas, São Paulo, Brazil.

[6]National Institute of Science and Technology in Bioanalytic Lauro Kubota (INCTBio-LK), São Paulo, Brazil.

[7]Department of Pharmacology and Toxicology, University of Toronto, Toronto, Ontario M5G 2C8, Canada

[8]Department of Mechanical and Aerospace Engineering, The Hong Kong University of Science and Technology, Hong Kong.

[9]Orange Biotech Limited, Hong Kong.

[10]The Structural & Biophysical Core Facility, The SickKids Research Institute, Toronto, ON, Canada.

[11]Department of Laboratory Medicine and Pathobiology, University of Toronto, Toronto, M5S 1A8 ON, Canada.

[12]Department of Microbiology, Sinai Health System/University Health Network, Toronto, M5G 1X5 ON, Canada.

[13]Department of Biomedical Engineering, Boston University, Boston, MA 02215, USA

[14]Molecular Biology, Cell Biology & Biochemistry Program, Graduate School of Arts and Sciences, Boston University, Boston, MA 02215, USA

[15]Biological Design Center, Boston University, Boston, MA 02215, USA

[16]Center for Systems and Synthetic Biology, University of Texas at Austin, Austin, TX, USA.

[17]LIVESTRONG Cancer Institutes, Dell Medical School, University of Texas at Austin, Austin, TX, USA.

*Co-first Authors
**Corresponding Authors


#### Materials and Methods

##### Key supplies

All oligonucleotides were synthesized by Integrated DNA Technologies (IDT); modified oligos were HPLC-purified, while unmodified ones underwent standard desalting. Commercial AsCas12a (Acidaminococcus sp., #10001272) was obtained from IDT, while LbCas12a (#M0653T), Cas12 diluent (#B0653A), and NEBuffer™ r2.1 (#B6002S), Cas12 reaction buffer (NEBuffer™ r2.1, #B6002S), rCutSmart buffer (#B6004S), and RNase inhibitor (#M0314L) were purchased from New England Biolabs. Murine RNase inhibitor (40 U/µL) without glycerol and dithiothreitol (DTT) (#RNI04-X25A) was obtained from SignalChem Diagnostics. Magnesium chloride solution (#7786303), potassium hexacyanoferrate(III) (#244023), potassium hexacyanoferrate(II) trihydrate BioUltra (#60279), Trizma® base (#T6066), Magnesium Acetate tetrahydrate (#M0631-100G), 6-mercapto-1-hexanol (#451088) and DNase/RNase-free water (#W4502) were obtained from Sigma Aldrich. Sodium chloride (#SOD002.205), magnesium chloride hexahydrate (#MAG510.500) and potassium acetate (#POA303.250) were obtained from BioShop Canada. Polyethylene glycol/dimethyl sulfoxide solution 50% (w/v) was purchased from Sigma Aldrich (#P7306). Plasmids purchased from Addgene used in our experiments are as follows: pMBP-AsCas12a (Addgene Plasmid #113430), pMBP-FnCas12a (Addgene Plasmid #113432), pET-28b-T7-bLbCas12a (Addgene Plasmid #113431), and pKEW336-MBP-TEV-MbCas12a-33362 (Addgene Plasmid #115670) were gifts from Jennifer Doudna. 6xHis-MBP-huBoCas12a (Addgene Plasmid #174673), 6xHis-MBP-huErCas12a (Addgene Plasmid #174685), 6xHis-MBP-huLb5Cas12a (Addgene Plasmid #174684), 6xHis-MBP-huPb2Cas12a (Addgene Plasmid # 174686), and 6xHis-MBP-huTsCas12a (Addgene Plasmid # 174687) were gifts from Piyush Jain.

##### Cas12a Purification

Lab-produced Cas12a proteins were expressed and purified as previously described by the Pardee Lab.[1] Briefly, each plasmid was transformed into E. coli NiCo21 cells (DE3, NEB, C2529H). Cultures were incubated overnight at 18 °C following induction, harvested by centrifugation, and pellets were frozen at -80 °C. Pellets were resuspended in lysis buffer [0.02 M Tris-HCl, 0.5 M NaCl, 10%(v/v) glycerol, pH 7.6], sonicated, and purified with the AKTA Pure Chromatography System (GE Healthcare, FPLC). Clarified lysate was loaded on a 5 mL HiTrap IMAC HP column (charged with Ni ions), washed with Wash Buffer [0.02 M Tris-HCl, 1 M NaCl, 10%(v/v) glycerol, pH 7.6 ], and eluted with Elution Buffer [0.25 M imidazole, 0.02 M Tris-HCl, 0.5 M NaCl, 10%(v/v) glycerol, pH 7.6]. Eluates were pooled, buffer exchanged and concentrated before Super TEV protease was added in a 1:50 TEV to protein mass ratio and incubated at 4 °C overnight. Cleaved proteins were further purified by loading on a 5mL HiTrap Heparin HP column in Loading Heparin Buffer [0.02 M Tris-HCl, 0.125 M NaCl, 5%(v/v) glycerol, pH 7.5], washed with Wash Buffer, and eluted with Elution Heparin Buffer [0.02 M Tris-HCl, 1M NaCl, 10%(v/v) glycerol, pH 7.5]. The elution fractions corresponding to the chromatography peak were analyzed with SDS-PAGE. Eluates were pooled, concentrated, and buffer exchanged into Storage Buffer, pH 7.6 [0.02 M Tris-HCl, 0.5 M NaCl, 0.0001 M EDTA, 0.0001 M TCEP, 50%(v/v) glycerol]. Purified proteins were stored at −20 °C. **Table 14** has all the Cas12a extensive information.

##### COMPANION Assay Setup

Duplex probes for miR-21-5p, miR-320a-3p, miR-340-5p, miR-210-3p, miR-187-3p, miR-193b-5p, miR-324-5p, miR-483-3p, miR-483-5p, and miR-103-3p were directly purchased from IDT in duplexed form (**Table 3**). Other duplex probes were prepared by direct annealing as follows: the annealing was conducted at 95°C in nuclease-free duplex buffer (IDT: # 11-05-01-12) for 4 minutes, followed by a gradient cooling step to 4°C at a rate of 0.1°C/s. Following the annealing process, CRISPR-Cas12 assay components were prepared in final concentration of 1× NEB rCutSmart buffer to a final reaction volume of 50 µL. Final concentrations of assay components were as follows: 150 nM Cas12a, 50 U RNase inhibitor, 500 nM reporter (Reporter 7), 40 nM duplex probe, and 50 target DNA or miRNA. This mixture was aliquoted in 15 µL volumes into a 384-well plate to enable three technical replicates per condition. Reactions were monitored for 2 hours at 37 °C using the BioTek plate reader. Sequences of all nucleic acids used are listed in **Tables 1, 2, 3,** and **4**. To compare performance with the traditional gRNA-programmed system, 90 nM of gRNA was added into the reaction (See **Extended Data Fig. 1, Supplementary Fig. 6, 7**).

**In vitro transcription of RNAs**

Many RNA molecules used throughout this study were generated by *in vitro* transcription (IVT) using a non-canonical, primer-assisted T7 transcription strategy specifically optimized for short RNAs. For each RNA, a single-stranded DNA oligonucleotide ("template DNA") was synthesized consisting of the reverse complement of the RNA sequence positioned at the 5′ end, followed directly by the reverse complement of the T7 promoter sequence at the 3′ end. This design differs from conventional IVT templates, which typically encode the T7 promoter upstream of the sense strand of the RNA transcript in double-stranded DNA form. In the present approach, formation of a functional T7 promoter duplex is achieved through annealing with a complementary T7 primer during the transcription reaction. A complementary T7 promoter oligo (referred to here as the *regular T7 complementary primer*; sequence: 5′-TAGCCATTAATACGACTCACTATAGG-3′) was obtained from IDT and supplied in trans to complete the double-stranded T7 promoter region required for transcription initiation. IVT reactions were performed using the HiScribe T7 High Yield RNA Synthesis Kit (NEB #E2040L). Each 20 µL reaction contained 1× T7 Reaction Buffer, 10 mM each of ATP, GTP, UTP, and CTP, 1.5 µM template DNA, 500 nM regular T7 complementary primer, and 5 mM dithiothreitol (DTT). Reactions were incubated at 37 °C for 4–16 h, depending on the desired RNA yield. As an example, transcription of miR-29b-3p (RNA sequence: 5′-UAGCACCAUUUGAAAUCAGUGUU-3′; **Table 1**) employed a template DNA sequence of 5′-AACACTGATTTCAAATGGTGCTA CCTATAGTGAGTCGTATTAATGGCTA-3′, where the 5′ segment (red) corresponds to the reverse complement of miR-29b-3p and the 3′ segment (blue) corresponds to the reverse complement of the T7 promoter. Following transcription, reactions were treated with RNase-free DNase I (NEB #M0303L) at a final concentration of 80 U $mL^{-1}$. The reaction volume was adjusted to 50 µL using DNase- and RNase-free water (Invitrogen; Lot #2937321) and incubated for an additional 15 min at 37 °C to remove residual DNA templates. RNA products were then purified using the Monarch RNA Cleanup Kit (NEB #T2040) according to the manufacturer's instructions and quantified using a NanoDrop One spectrophotometer (Thermo Fisher Scientific). Sequences of all transcribed RNA products used in this study are provided in **Tables 1,4,11,12**.

**Reporter screening for COMAPNION**

To screen the 16 different reporters (including DNA/RNA/chimeric sequences, DNA homopolymers, RNA homopolymers, and chimeric homopolymers – **Table 9**) used in this study, a 50 µL reaction mixture was prepared as follows: 1× rCutSmart NEB buffer, 150 nM Cas12a, 50 U RNase inhibitor, 40 nM duplex probe, 100 nM miRNA21 trigger, and 500 nM of the various reporter types. This mixture was aliquoted in 15 µL volumes into a 96-well plate to enable three technical replicates per condition. Reactions were monitored for 2 hours at 37 °C using the BioTek plate reader. All the reporters were purchased from IDT.

**Mismatch detection in miRNA21 and DNA21**

All templates containing single or double mismatches in ssDNA were obtained from IDT. A 50 µL reaction mixture was prepared containing the following components: 1× rCutSmart NEB buffer, 150 nM AsCas12a, 50 U RNase inhibitor, 40 nM duplex probe, 500 nM ssDNA reporter (R7), and 100 nM ssDNA with mismatches or WT. This mixture was aliquoted in 15 µL volumes into a 96-well plate to enable three technical replicates per condition. Reactions were monitored for 2 hours at 37 °C using the BioTek plate reader. All the sequences are contained in **Tables 11 & 12**.

**COMPANION-RT-LAMP assay setup**

RT-LAMP reactions were performed using WarmStart LAMP 2× Master Mix (NEB, E1700S) in a total volume of 10 µL. Final primer concentrations were 0.2 µM for F3 and B3, 1.6 µM for FIP and BIP, and 0.4 µM for LF and LB. Each reaction contained 1.0 µL of template (nuclease-free water, *in vitro*–transcribed RNA, or extracted RNA) (Primer location, **Table 13**). Reaction mixtures were assembled on ice and incubated at 60 °C for 30 min, followed by heat inactivation at 80 °C for 5 min.

Following RT-LAMP amplification, 5 µL of the reaction product was directly transferred into a COMPANION mixture, resulting in a final reaction volume of 50 µL. The COMPANION reaction included 1× rCutSmart buffer, RNase inhibitor (50 U per reaction), 500 nM reporter 7, 150 nM AsCas12a, and 40 nM each of the F and B miniCaRs (see **Table 13** for sequences). Reactions were prepared on ice, and each 50 µL reaction was divided into three technical replicates by dispensing 15 µL per well into a 384-well clear-bottom microplate for fluorescence measurements. Fluorescence was measured at 37 °C using a BioTek plate reader.

**RT-qPCR reference assays for FluA H1N1, RSV A, and Flu B**

Patient samples were determined as positive or negative by RT-qPCR using WHO-established diagnostic assays, with Ct values ≤40 indicating positivity. RT-qPCR assays targeting Flu A (H1N1), RSV A, and Flu B were performed using the QuantiNova Probe RT-PCR Kit (Qiagen, 208354) according to the manufacturer's instructions. Briefly, each RT-qPCR reaction had a total volume of 10 µL, consisting of 6.5 µL of reaction mix and 3.5 µL of template (nuclease-free water for non-template reactions, *in vitro*–transcribed RNA, or extracted RNA), and was dispensed into a 384-well plate. Primer–probe sets specific to each viral target, and the human RNase P internal control were synthesized by IDT. Fluorescence output and data analysis were carried out on an Applied Biosystems QuantStudio 5 real-time PCR instrument.

**Patient sample collection and ethical considerations**

A total of 102 de-identified nasopharyngeal swab specimens were collected at Mount Sinai Hospital (Toronto, Canada) from patients presenting with suspected respiratory infections as part of routine clinical care. Samples were processed by the hospital's clinical diagnostics laboratory, and the extracted RNAs were subsequently provided for this study. All procedures involving patient samples were approved by the University of Toronto Research Ethics Board (protocol #: 00039531) and conducted in accordance with the Declaration of Helsinki and all applicable ethical guidelines and regulations.

**Electrochemical Experiments**

*Electrode Functionalization*: The immobilization of thiolated ssDNA reporter sequences (**Table S15**) onto the screen-printed gold electrode (SPGE) (#LX-9X85204FA-R, Lixens) was initiated by reducing disulfide bonds with 12.5 mM Tris(2-carboxyethyl)phosphine (TCEP) at 37 °C for 30 min in Tris buffer (10 mM

Tris-HCl, 100 mM NaCl, 10 mM $MgCl_2$, pH 8.0). The treated 5 µM SH-ssDNA solution was drop-cast (12 µL) onto the electrode surface and incubated overnight at room temperature in the dark. The Au/SH-ssDNA-modified electrodes were rinsed with deionized water and immersed in measurement buffer (50 mM Potassium Acetate, 20 mM Tris-acetate, 10 mM Magnesium Acetate, pH 7.9) for 1 h. Subsequently, the electrodes were functionalized with 12 µL of 1 mM 6-mercaptohexanol (MCH) aqueous solution for 15 min at room temperature to block nonspecific binding sites and for aligning the DNA monolayer, followed by immersion in measurement buffer (50 mM Potassium Acetate, 20 mM Tris-acetate, 10 mM Magnesium Acetate, pH 7.9) for 1 h. For short-term storage, the modified electrodes were kept in measurement buffer at 4 °C.

*CRISPR Reaction*: A 50 µL CRISPR/Cas12a reaction mixture was prepared in reaction buffer (50 mM Potassium Acetate, 20 mM Tris-acetate, 10 mM Magnesium Acetate, pH 7.9) containing 150 nM AsCas12a (or Cas12a ortholog), 50 U RNase inhibitor, 40 nM duplex probe and the respective concentration of miRNA target. Subsequently, 12 µL of the activated mixture of Cas12a, RNase inhibitor, duplex probe and target was drop-cast onto a screen-printed gold electrode (SPGE, Lixens) and incubated at 37 °C for 1h30min to promote trans-cleavage of the surface-bound ssDNA reporter. After incubation, electrodes were rinsed with RNase-free water and immersed in measurement buffer (50 mM Potassium Acetate, 20 mM Tris-acetate, 10 mM Magnesium Acetate, pH 7.9) for 20 min prior to electrochemical analysis.

*Electrochemical Analysis*: Electrochemical measurements were performed using a screen-printed gold electrode (SPGE, Lixens) connected to a potentiostat (Sensit BT, PalmSens, Netherlands) controlled via PSTrace 5.11 software. Measurements were carried out in measurement buffer (50 mM Potassium Acetate, 20 mM Tris-acetate, 10 mM Magnesium Acetate, pH 7.9) containing 5 mM $[Fe(CN)_6]^{3-/4-}$ redox probe. Cyclic voltammetry (CV) was conducted in the potential range from −0.45 V to 0.45 V (vs Au) at a scan rate of 0.05 V $s^{-1}$. Electrochemical impedance spectroscopy (EIS) was recorded at open circuit potential (E = 0.00 V) over a frequency range of 50 kHz to 1 Hz. Impedance data were fitted using a Randles equivalent circuit to extract the charge transfer resistance (Rct).

The performance of the CRISPR/Cas12a system was evaluated by comparing Rct values before and after *trans*-cleavage. The cleavage efficiency was calculated as the percentage reduction in Rct using the equation:

$$Signal\ Change\ (\%) = \frac{\Delta Rct}{Rct_0} = \frac{Rct_0 - Rct_{CRISPR}}{Rct_0} \times 100\%$$

where $R_{ct0}$ and $R_{ctCRISPR}$ correspond to the Rct values before and after the CRISPR/Cas12a reaction, respectively.

**Working Principle of the Electrochemical COMPANION Biosensor**

The detection principle of the label-free electrochemical biosensor relies on the collateral cleavage of non-specific DNA reporter sequences by the target-activated CRISPR/Cas12a with PAM-duplex in the presence of the miRNA target. This cleavage event induces a change in the charge-transfer resistance (Rct) at the interface between the SH-ssDNA-modified electrode and the electrolyte, which is monitored by electrochemical impedance spectroscopy (EIS) in the presence of the $[Fe(CN)_6]^{3-/4-}$ redox probe. The Nyquist plots shown in Figure 5a exhibit semicircles corresponding to the electron-transfer kinetics of the $[Fe(CN)_6]^{3-/4-}$ probe at the electrode surface, where a decrease in the semicircle diameter reflects a lower

Rct. When the electrode is incubated with the non-activated CRISPR/Cas12a complex, no change in Rct is observed, confirming the absence of ssDNA reporter cleavage. In contrast, incubation with the target-activated Cas12a/duplex complex (1 nM of miRNA-340) resulted in a clear decrease in Rct.

**Fluorescence Polarization**

Fluorescence polarization measurements were performed to assess binding of AsCas12a to fluorescently labeled probes (see **Table 7** for sequences). Recombinant AsCas12a was prepared in assay buffer (PBS, 10 mM $MgCl_2$, 0.01% Triton) and serially diluted to the indicated concentrations. Next, fluorescently labeled probes were added to each well to achieve a final concentration of 1 nM. Binding experiments were assembled in a total volume of 10 µL in black, low-binding 96-well plates, and incubated for 2 minutes at room temperature prior to measurement. Fluorescence polarization was measured using a plate reader equipped for detection (excitation 485 nm, emission 528 nm, gain = 120). Polarization values were recorded in millipolarization (mP) units. Each condition was measured in duplicate. Data were analyzed by plotting polarization as a function of AsCas12a concentration, and binding curves were generated using GraphPad Prism 10. For competition experiments, miRNA-21 or gRNA were prepared in assay buffer and serially diluted to indicated concentrations. AsCas12a and miRNA-21-FAM concentrations were held constant at 20 nM and 1 nM, respectively.

**Structural Modeling**

Protein-nucleic acid complexes were generated using Boltz2,[2] an open-source deep learning co-folding model that models multiple molecular chains simultaneously and outputs atomic-resolution structural predictions. For each complex, input configurations were prepared in YAML format with unique identifiers and sequences. Multiple sequence alignments were generated where appropriate using Colabfold[3] to inform structural context, and models in mmCIF format were generated using default parameters (three recycling steps, 200 sampling steps, one diffusion sample, and a step scale of 1.638). Post-processing and alignment were performed using PyMOL 3.1.6.1.

**miRNA-DNA Ligation**

For miRNA-DNA ligation, two DNA fragments and the miRNA target were annealed under the same conditions described above for duplex probes. The reaction components and conditions were identical to the duplex probe protocol. For both reactions, the T4 DNA ligase enzyme was excluded in conditions where ligation was not required. This method allowed for precise monitoring of *trans* cleavage activity under ligation and non-ligation conditions, enabling the evaluation of miRNA or DNA detection (see nucleic acid in **Tables 3&6**).

**Limit of Detection (fluorescence)**

COMPANION reactions were assembled by mixing AsCas12a at a final concentration of 150 nM in 1X rCutSmart buffer supplemented with 1U/µL murine RNase inhibitor (NEB). The fluorophore-quencher reporter (R7) and duplex probe were added to the Cas12 reaction at final concentrations of 500 nM and 40 nM, respectively, along with other various concentrations (at 20 µL) of the miRNA target in a 50 µL reaction volume on ice. Reactions were transferred into optical 96-well plates at 15 uL and read on the plate reader in real-time at 37 °C for 2 hours while the FAM fluorescent intensity was measured every minute. All oligos are listed in **Tables 1 to 3**. Limit of detection of the miRNAs were calculated from $3\sigma/S$, where $\sigma$ is the standard deviation of the background signal and S is the slope of the fluorescence of target concentration line.

**Cryo-EM sample preparation, data collection and processing**

Duplex DNA was prepared by mixing equimolar of two DNA strands in annealing buffer (10 mM Tris-HCl, pH 7.5, 50 mM NaCl, 1 mM DTT), followed by heating at 95 °C for 5 min, and cooling at room temperature for 15 min. gRNA and target RNA were prepared separately as described above. A flash frozen sample of purified AsCas12a was rapidly thawed and mixed with target RNA and duplex DNA at a molar ratio of 1:1.5:1.5, followed by incubation at 37 °C for 30 min to form the COMPANION complex, while the additional gRNA was included for RAPID sample.

2.5 µL of 7 µM sample was applied to Quantifoil 1.2/1.3 400-mesh copper grid that had been glow discharged at 20 mA for 30 s by a Solarus 950 plasma cleaner (Gatan). Grids were blotted using a Vitrobot Mark IV (Thermo Fisher) for 8-12 s with blot force of 1 at 4 °C and 100% humidity, and plunge-frozen in liquid ethane. Grids were stored in liquid nitrogen before screening.

Data were collected on an FEI Titan Krios 300 V Transmission Electron Microscope (Thermo Scientific) equipped with a Gatan BioContinuum Imaging Filter and a K3 direct electron detector. Data were collected on SerialEM (v4.1)[4] with a 0.8332 Å pixel size and a defocus range of –1.5 to –2.5 µm with a total exposure time of 3.6 s in a total accumulated dose of $70e^{-}/A^{2}$. Motion correction, contrast transfer function (CTF) estimation, particle picking, extraction and 2D classification were carried out in cryoSPARC live (v4.2). Subsequent processing steps were performed in cryoSPARC (v4.7.1).[5]

For COMPAINION, 2,639 micrographs were accepted. Particles were picked using blob picker with a particle diameter range of 80 - 180 Å. 1,932,845 particles were extracted at a box size of 320 pixels with a Fourier crop to 128 pixels and then classified into 50 2D classes. 637,907 particles were selected from 2D classification for further ab-initio reconstruction (3 classes). The resulting particles and volume classes were subjected to heterogeneous refinement. The best-resolved class was selected for particle re-extraction at box size of 320 pixels followed by non-uniform refinement. To mitigate the preferred orientation of particles, an additional dataset containing 3,788 micrographs was collected at –30° tilting and processed as described above. 921,805 extracted particles at box size of 320 pixels from these two datasets were combined to generate a map via non-uniform refinement. Anisotropy was improved by rebalance orientation, while the resolution was improved by reference-based motion correction and global/local CTF refinement yielding a volume at 3.14 Å (**Suppl Fig. 2**, **3**). The final map was processed using EMReady2.[6]

For RAPID, 7,814 micrographs were accepted. 3,235,710 particles were selected using blob picker. 1,759,454 particles were picked after a single round of 2D classification. A random subset of 100,000 picked particles was used for ab-initio reconstruction (3 classes). The best class was used for heterogeneous refinement and downstream non-uniform refinement, yielding a 2.88 Å reconstruction comprising 784,461 particles. The particles used for the reconstruction were then subjected to a single round of 3D classification (10 classes) to sort out heterogeneity. Classes with similar particle compositions were combined for a one final round of non-uniform refinement, yielding a target bound complex at 3.00 Å resolution comprised of 181,085 particles and a pre-target bound 3.16 Å structure comprised of 67,054 particles. The final map was processed using EMReady2.[6]

**Model building and refinement**

For COMPANION, the complex structure predicted by AlphaFold3[7] was fitted into the map as a rigid body in ChimeraX (v1.9).[8] The model was manually adjusted and refined in COOT (v0.9.8.95 EL)[13] and automatically refined by real_space_refine in PHENIX (v2.0-5936).[9] For RAPID, the initial backbone of the protein was built by rigid-body fitting a previously solved structure of AsCas12a (PDB: 8SFO) [3]. The output from this displayed a relatively well-aligned nucleic acid backbone as well as protein secondary structure. The output was used as a fiducial for de-novo model building using a combination of COOT (v1.0)[10] and ChimeraX (v1.9).[8] Once fully modeled, Isolde (v1.4)[11] was used to improve the model to map fit and for rotamer optimization. Multiple iterations of real space refinement implemented within Phenix (v1.19)[9] were carried out to improve the model geometry and map fit. All structural figures were generated using ChimeraX. (v1.9).

**Mass spectrometry analysis of Cas12a-mediated reporter cleavage**

Synthetic reporters R1 (TTATT) and R7 (CCCCCC), without any modification of labelling, were obtained from Integrated DNA Technologies (IDT) with standard desalting purification. Cleavage reactions (200 µL total volume) were assembled in 1× NEBuffer rCutSmart (New England Biolabs) and contained 150 nM AsCas12a, 90 nM miniCaR targeting influenza A ssDNA, and 50 nM influenza A ssDNA target. Reactions further included 6 µM reporter (R1, or R7), 1 U $\mu L^{-1}$ murine RNase inhibitor. Samples were incubated at 37 °C for 2 h to allow Cas12a activation and *trans cleavage* of reporter substrates.

Following incubation, reactions were subjected to molecular weight filtration using 10 kDa Amicon Ultra centrifugal filters (Millipore) at 16,000g for 2 h to remove Cas12a protein, ribonucleoprotein complexes, and other high-molecular-weight components. The filtrate, containing low-molecular-weight nucleic acid fragments, was collected and directly subjected to LC–ESI-MS analysis.

**LC–ESI-MS conditions**

Separation of cleavage products was performed using hydrophilic interaction liquid chromatography (HILIC) on a Thermo Scientific Ultimate 3000 UHPLC system equipped with a Waters Atlantis Premier BEH Z-HILIC column (1.7 µm, 2.1 × 100 mm) and guard column. The column was maintained at 40 °C, and the autosampler was held at 5 °C. A 10 µL sample volume was injected for each analysis. Mobile phase A consisted of 15 mM ammonium bicarbonate (pH 9.0) in water, and mobile phase B consisted of 15 mM ammonium bicarbonate (pH 9.0) in 90% acetonitrile/10% water. The flow rate was set to 0.5 mL $min^{-1}$. The gradient program was as follows: 0 min at 90% B; linear gradient to 65% B from 0–5 min; hold at 65% B from 5–6 min; return to 90% B from 6–7 min; and re-equilibration at 90% B from 7–13 min.

Mass spectrometric analysis was carried out on a Thermo Scientific Q Exactive Orbitrap mass spectrometer equipped with a heated electrospray ionization (HESI II) source, operated in negative ion mode. Source parameters were set as follows: spray voltage, 3.5 kV; capillary temperature, 325 °C; sheath gas flow rate, 40 arbitrary units; auxiliary gas, 20; sweep gas, 5; and S-lens RF level, 55.

**Mass spectrometry acquisition and data analysis**

Full MS scans were acquired over an m/z range of 200–1200 at a resolution of 70,000 (at m/z 200), with an automatic gain control (AGC) target of $3 \times 10^6$ and a maximum injection time of 200 ms. Data-dependent MS/MS (Top10) was performed at a resolution of 17,500, with an AGC target of $1 \times 10^5$, maximum injection time of 50 ms, isolation window of 0.4 m/z, and normalized collision energy of 35. Raw data were processed using Thermo Scientific Qual Browser (v4.5.445.18). Peak assignment was performed based on expected nucleotide fragment masses with a tolerance of ±5 ppm. Retention times (RT), mass-to-charge ratios (m/z), and normalized signal intensities were extracted and compiled for each reporter system. Fragment identities were confirmed by comparison with theoretical masses of expected mono- and oligonucleotide products.

**Electrophoretic mobility shift assay**

Electrophoretic mobility shift assays were performed to assess binding of catalytically inactive AsCas12a (D908A) to fluorescently labeled nucleic acid substrates. Binding reactions were assembled in 20 µL volumes containing 1× EMSA binding buffer (20 mM Tris pH 7.5, 100 mM NaCl, 5% glycerol, 5 mM β-mercaptoethanol, and 0.05 mg/mL BSA), 5 mM $MgCl_2$, 100 nM FAM-labeled RNA or DNA substrate, and the indicated concentrations of dAsCas12a. Reactions were incubated for 30 min at room temperature and loaded onto native polyacrylamide gels. Gels were prepared as 6% native polyacrylamide gels in 0.5× TB using 30% acrylamide/bis-acrylamide solution. Gels were pre-run in 0.5× TBE for approximately 30 min at 100 V before sample loading. Electrophoresis was performed in 0.5× TB at 150 V under cooled conditions for 30 minutes. Following electrophoresis, gels were imaged directly using Cytivia Amersham™ ImageQuant™ 800 configured for FAM/fluorescein detection. Binding was assessed by depletion of the free substrate band and appearance of slower-migrating protein–nucleic acid complexes or retarded/smeared signal. For RNA substrates that displayed multiple protein-independent bands, binding was interpreted primarily from protein-dependent depletion of the lower free-RNA species and/or appearance of retarded material relative to the no-protein control.

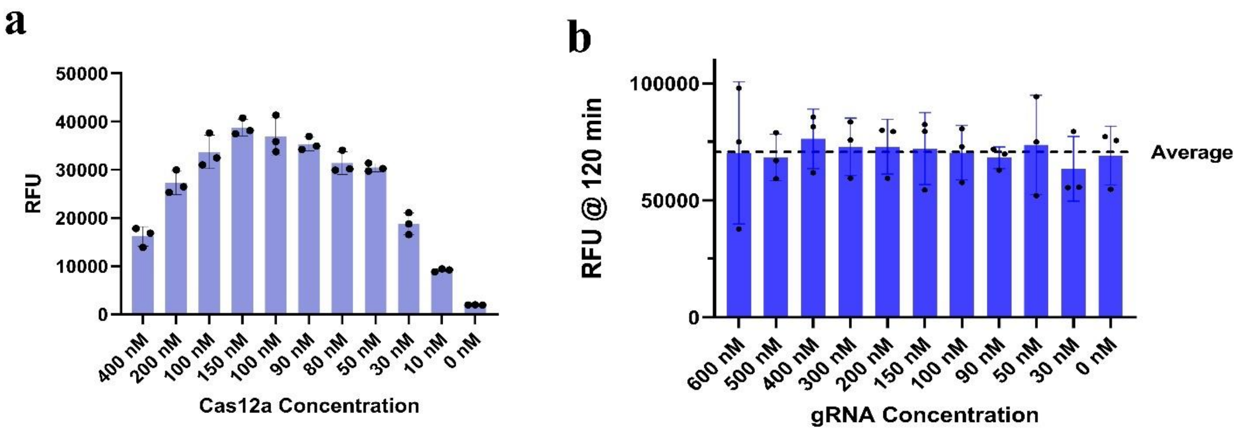

**Supplementary Fig. 1. Cas12a optimization and emergence of the crRNA-independent CRISPR/Cas12a platform.** (a) Bar graph showing AsCas12a *trans* cleavage activity optimization in the previously reported RAPID[12] assay with *in situ* ligation for miR-21 target. (b) *Trans* cleavage signal at 120 min for varying crRNA concentrations for miR-21 detection. Even in the absence of crRNA (0 nM), Cas12a was fully activated, indicating no dose-dependent effect. This finding led to the development of the crRNA-free CRISPR/Cas12a platform. All data are shown as mean ± SD ($n \geq 3$ technical replicates). Standard RAPID buffer condition for *in situ* ligation was used.

**Note**

Our crRNA-independent system began by optimizing the CRISPR/Cas12a reaction conditions using our previous RAPID platform[12] as a starting point, for miR-21 detection. Standard RAPID buffer condition for *in situ* ligation was used.[12] We titrated the AsCas12a concentration and identified an optimal enzyme level of ~150 nM for robust *trans* cleavage activity (**Suppl Fig. 1a**). Using this optimized Cas12a concentration, we next evaluated the effect of crRNA concentration on the reaction. Unexpectedly, we observed that varying the crRNA from 0 to 600 nM had no impact on the readout, even reactions completely lacking a crRNA exhibited *trans* cleavage activity comparable to those with saturating crRNA (**Suppl Fig. 1b**). This result was unexpected, as Cas12a typically requires a crRNA-bound complex to recognize targets.[13] We repeated this experiment multiple times and obtained the same outcome, confirming that under these conditions the presence of a crRNA was dispensable for Cas12a activation. Notably, this crRNA-independent activation phenomenon only occurred at high Cas12a concentrations under the prevailing standard Cas12 buffer condition and was not seen at lower enzyme levels, suggesting a threshold effect for unleashing Cas12a's activity in the absence of a guide. However, the optimised buffer condition allows the crRNA-independent CRISPR/Cas12a platform to function at lower Cas12a concentration. This surprising finding laid the foundation for our subsequent discoveries.

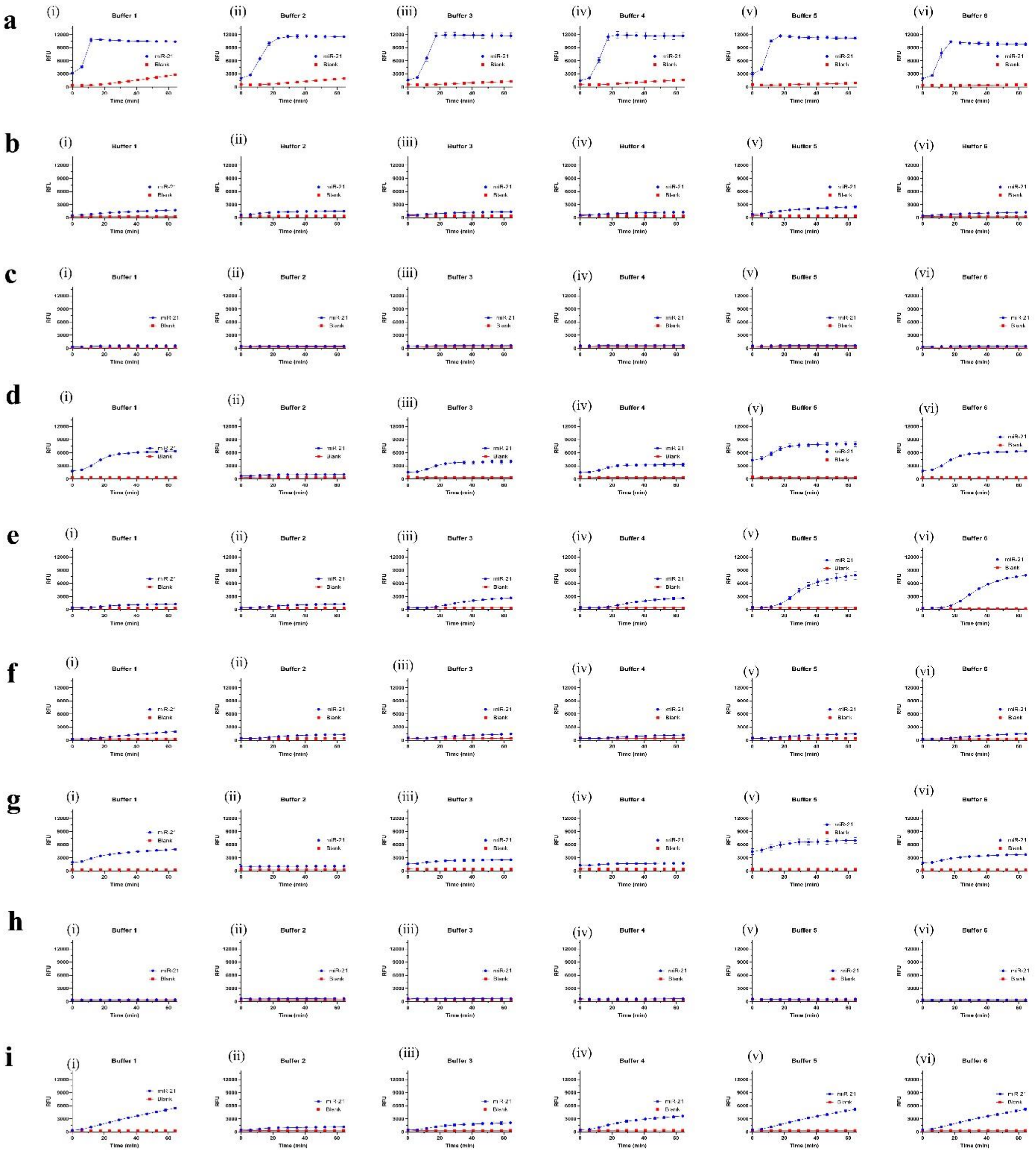

**Supplementary Fig. 2.** Buffer optimization for crRNA-free Cas12a system across 9 Cas12a orthologs. Systematic buffer screening for: (**a**) AsCas12a, (**b**) BoCas12a, (**c**) Lb5Cas12a, (**d**) MbCas12a, (**e**) ErCas12a, (**f**) FnCas12a, (**g**) LbCas12a, (**h**) TsCas12a, and (**i**) Pb2Cas12a. Six buffer formulations were tested with Reporter 1. **Buffer 1** (1x conc.): 30 mM Tris-HCl (pH 7.8, 25 °C), 10 mM $MgCl_2$, 10 mM DTT, 1 mM ATP. **Buffer 2** (1x conc.): 6% PEG8000, 50 mM NaCl, 10 mM Tris-HCl, 41.25 mM $MgCl_2$, 100 µg/ml recombinant albumin, pH 7.9 (25 °C). **Buffer 3** (1x conc.): 50 mM NaCl, 10 mM Tris-HCl, 10 mM $MgCl_2$, 100 µg/ml recombinant albumin, pH 7.9 (25 °C). **Buffer 4** (1x conc.): 100 mM NaCl, 50 mM Tris-HCl, 10 mM $MgCl_2$, 100 µg/ml recombinant albumin, pH 7.9 (25 °C). **Buffer 5** (1x conc.): 50 mM potassium acetate, 20 mM Tris-acetate, 10 mM magnesium acetate, 100 µg/ml recombinant albumin, pH 7.9 (25 °C).

**Buffer 6** (1x conc.): 50 mM Tris-HCl, 10 mM $MgCl_2$, 1 mM ATP, 10 mM DTT, pH 7.5 (25 °C). Optimization revealed buffer-dependent performance heterogeneity across orthologs, drawing attention to the need for ortholog-tailored buffer conditions. Blue curve is the positive control (with miRNA-21), while the red is negative control (blank or without miRNA-21).

**Note.**

**Buffer Optimization**: We first optimized the reaction buffer across a panel of Cas12a orthologs using Reporter 1 (R1) as the base ssDNA *trans* cleavage substrate. Six buffer formulations (**Suppl Fig. 2**) were tested to monitor Cas12a *trans* cleavage. The buffers are: **Buffer 1** (1×): 30 mM Tris–HCl (pH 7.8, 25 °C), 10 mM $MgCl_2$, 10 mM DTT, 1 mM ATP. **Buffer 2** (1×): 6% PEG 8000, 50 mM NaCl, 10 mM Tris–HCl, 41.25 mM $MgCl_2$, 100 μg/mL recombinant albumin, pH 7.9 (25 °C). **Buffer 3** (1×): 50 mM NaCl, 10 mM Tris–HCl, 10 mM $MgCl_2$, 100 μg/mL recombinant albumin, pH 7.9 (25 °C). **Buffer 4** (1×): 100 mM NaCl, 50 mM Tris–HCl, 10 mM $MgCl_2$, 100 μg/mL recombinant albumin, pH 7.9 (25 °C). **Buffer 5** (1×): 50 mM potassium acetate, 20 mM Tris–acetate, 10 mM magnesium acetate, 100 μg/mL recombinant albumin, pH 7.9 (25 °C). **Buffer 6** (1×): 50 mM Tris–HCl, 10 mM $MgCl_2$, 1 mM ATP, 10 mM DTT, pH 7.5 (25 °C).

Using the Reporter 1 (R1) and a panel of Cas12a orthologs (AsCas12a, BoCas12a, Lb5Cas12a, MbCas12a, ErCas12a, FnCas12a, LbCas12a, TsCas12a, Pb2Cas12a), we observed that buffer composition had a pronounced impact on enzyme performance. Each ortholog exhibited an optimal buffer condition, highlighting the need for ortholog-tailored reaction conditions. We selected **Buffer 5** (which is identical to NEB's rCutSmart buffer) as a versatile choice for subsequent experiments, as it supported robust activity across most Cas12a variants. This observation is consistent with reports that different Cas12a enzymes have distinct biochemical requirements and activities.[14]

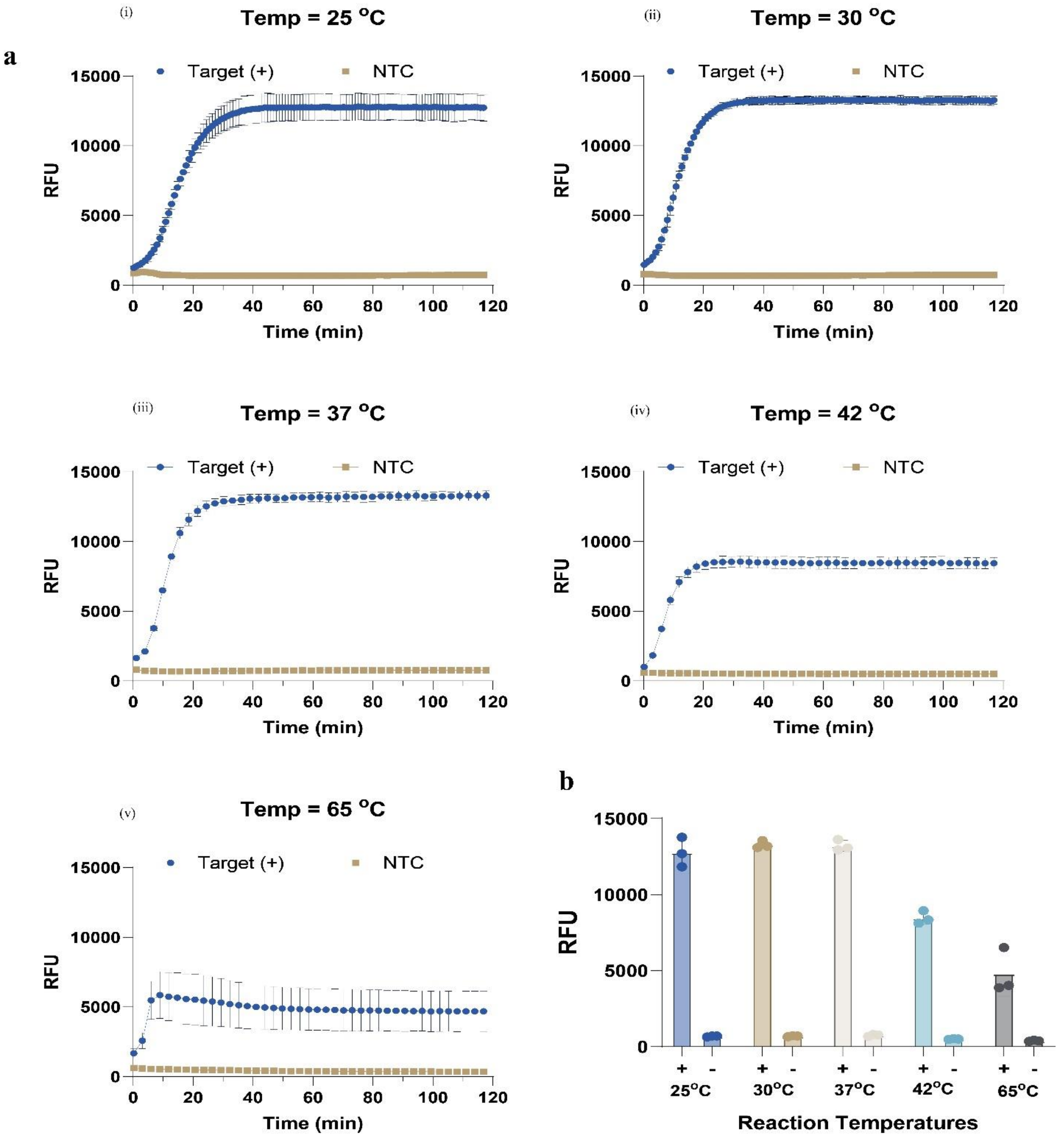

**Supplementary Fig. 3.** Temperature dependence activity with AsCas12a. (a) Kinetic activity traces of crRNA-independent system at 25 °C (i), 30 °C (ii), 37 °C (iii), 42 °C (iv), and 65 °C (v). (b) Quantitative comparison of endpoint activity at 60 min, showing optimal performance at physiological and slightly elevated temperatures.

**Note.**

**Temperature tolerance of the crRNA-independent reaction**: Focusing on AsCas12a (a widely used Cas12a), we screened reaction temperatures from 25 °C to 65 °C (**Suppl Fig. 3**). Remarkably, AsCas12a retained activity even at 65 °C, a temperature at which many standard Cas12a orthologs begin to unfold or lose function.[14] This thermostability opens the door to coupling our crRNA-independent system with high-temperature isothermal amplification methods such as LAMP, which typically operates around 60–65 °C.[15] However, we noted a trade-off at elevated temperatures: above ~42 °C the real-time fluorescence signal decreased (indicating lower *trans* cleavage reporter signal), even though the reaction endpoint and speed were not significantly compromised. This finding aligns with previous reports that wild-type Cas12a enzymes have suboptimal *trans* cleavage activity at higher temperatures, often necessitating separate two-step workflows or enzyme engineering for one-pot reactions.[15] To balance these factors, we chose **37 °C** for all further experiments, a temperature that preserves strong signal output and fast reaction kinetics, and conveniently matches the optimal range of most Cas12a enzymes and amplification enzymes in combined assays.[15]

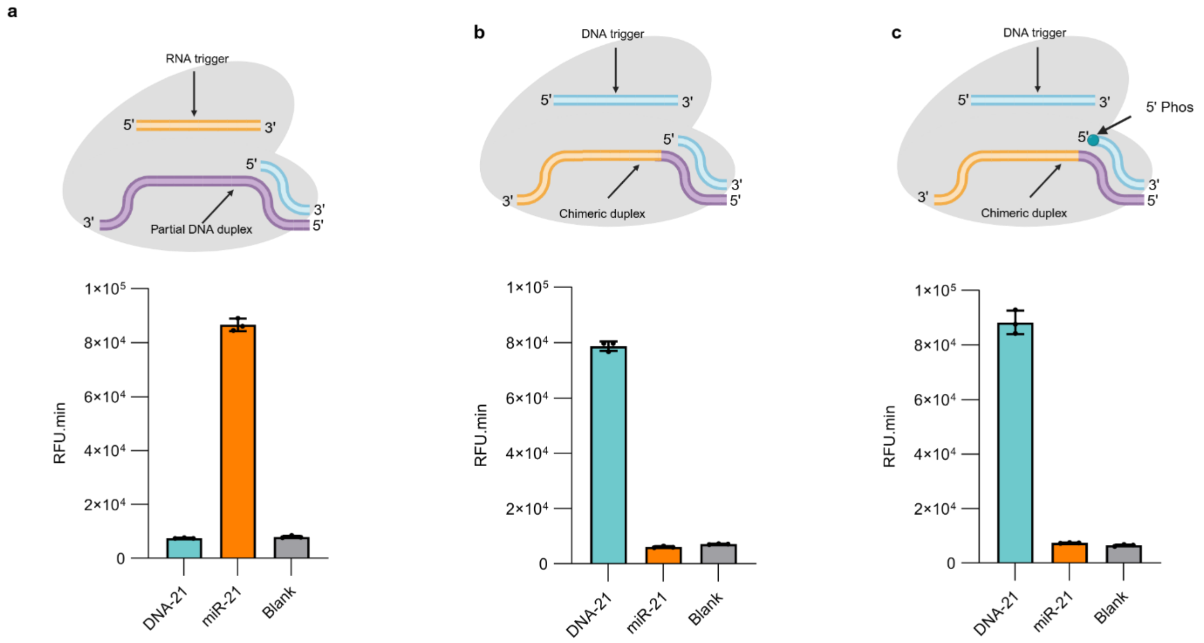

**Supplementary Fig. 4. Duplex probe configuration for crRNA-independent CRISPR/Cas12a system.** **(a)** miR-21 is identified using a partial DNA duplex probe. **(b)** miR-21 DNA equivalence (DNA-21) detection using a partial DNA-RNA duplex probe. **(c)** DNA-21 identification via in situ ligation, with a 5′-phosphorylated partial DNA-RNA duplex probe. All data are presented as mean ± SD for n ≥ 3 technical replicates. miniCaR here is miR-21. Nucleic acid colours: RNA is orange, DNA is cyan and purple. Duplex probe comprises dsDNA region with a single-stranded overhang that selectively interact with miniCaR or DNA. All sequences can be found in **Tables 2-9**.

**Note**

ssDNA and dsDNA identification. To extend our crRNA-independent system toward direct DNA detection, we adapted Modes II and IV (**Fig. 2a(ii, iv)**) to enable targeting of both ssDNA and dsDNA substrates. In this configuration, miniCaRs were designed to engage sequence elements corresponding to Influenza A, Influenza B, and RSV A, targeting regions either proximal to the canonical TTTV PAM or entirely independent of PAM constraints (**Extended Data Fig. 1(a-c)**). Remarkably, crRNA-free platform enabled robust and sequence-specific identification of both ssDNA and dsDNA targets across all tested conditions, confirming that DNA target recognition can be achieved without reliance on PAM-mediated licensing.

To further interrogate positional and sequence constraints, we systematically designed miniCaRs against multiple loci within an RSV A dsDNA substrate spanning a range of GC contents (**Extended Data Fig. 1d**). The system consistently detected all target regions in a fully PAM-independent manner, demonstrating broad targeting flexibility and tolerance to sequence composition (**Extended Data Fig. 1e**). Notably, under optimized buffer conditions (equivalent to NEB rCutSmart), crRNA-independent platform maintained strong activity across a wide range of AsCas12a concentrations (~50 nM to >150 nM), highlighting the robustness and tunability of this guide-free activation mechanism. Together, these results establish our platform as a versatile platform for direct DNA detection, significantly expanding the targeting scope of CRISPR/Cas12a systems.

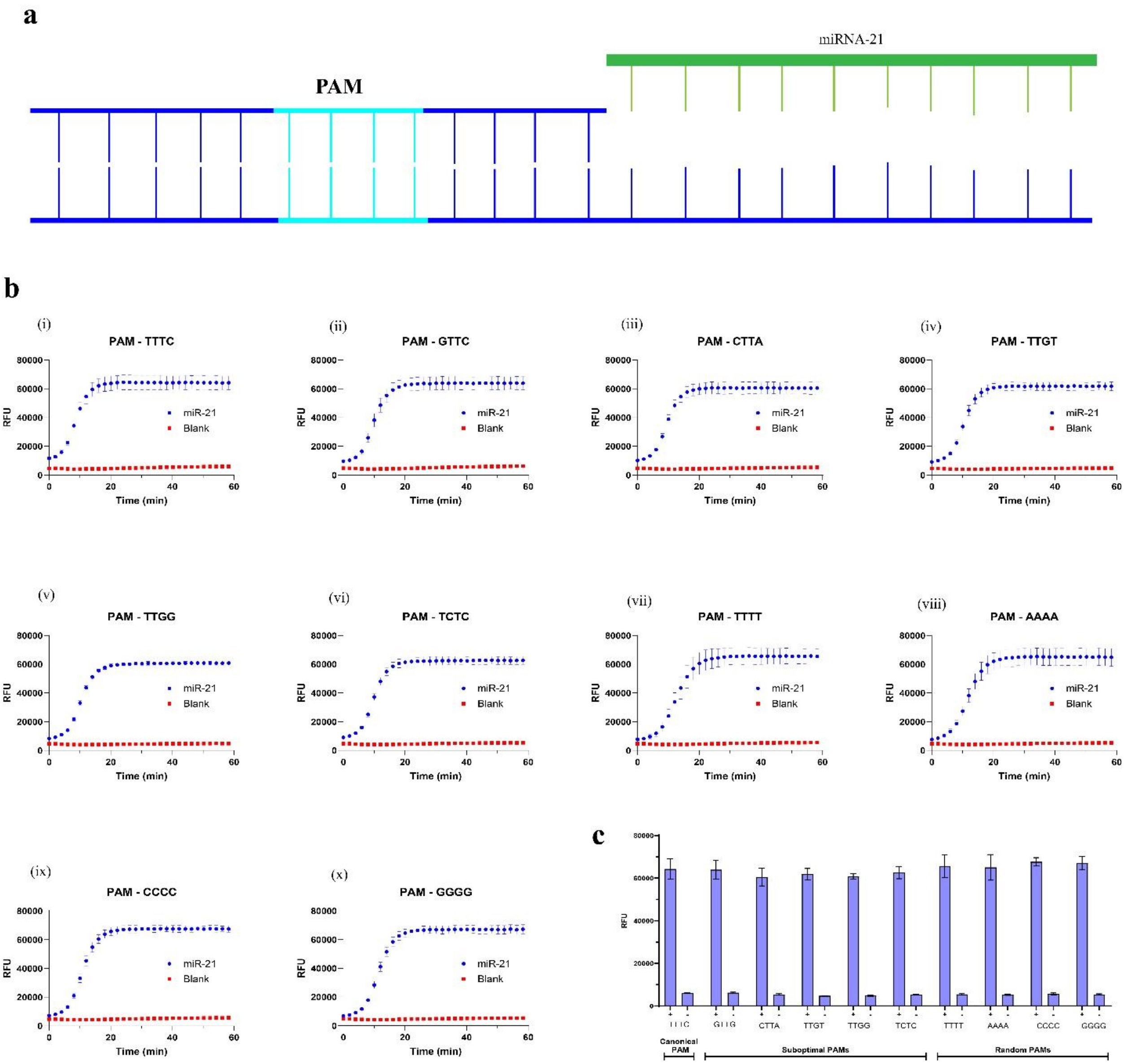

**Supplementary Fig. 5. Role of PAM combinations.** (a) Schematic of a PAM-containing duplex probe designed to target miRNA-21. (b) Kinetic cleavage profiles comparing canonical PAM (i), suboptimal PAMs (ii–vi), and random PAMs containing mononucleotide repeats (vii–x). (c) End-point analysis of cleavage activity (60 min) across all PAM variants, quantified by fluorescence intensity, demonstrating tolerance to PAM diversity while retaining activity.

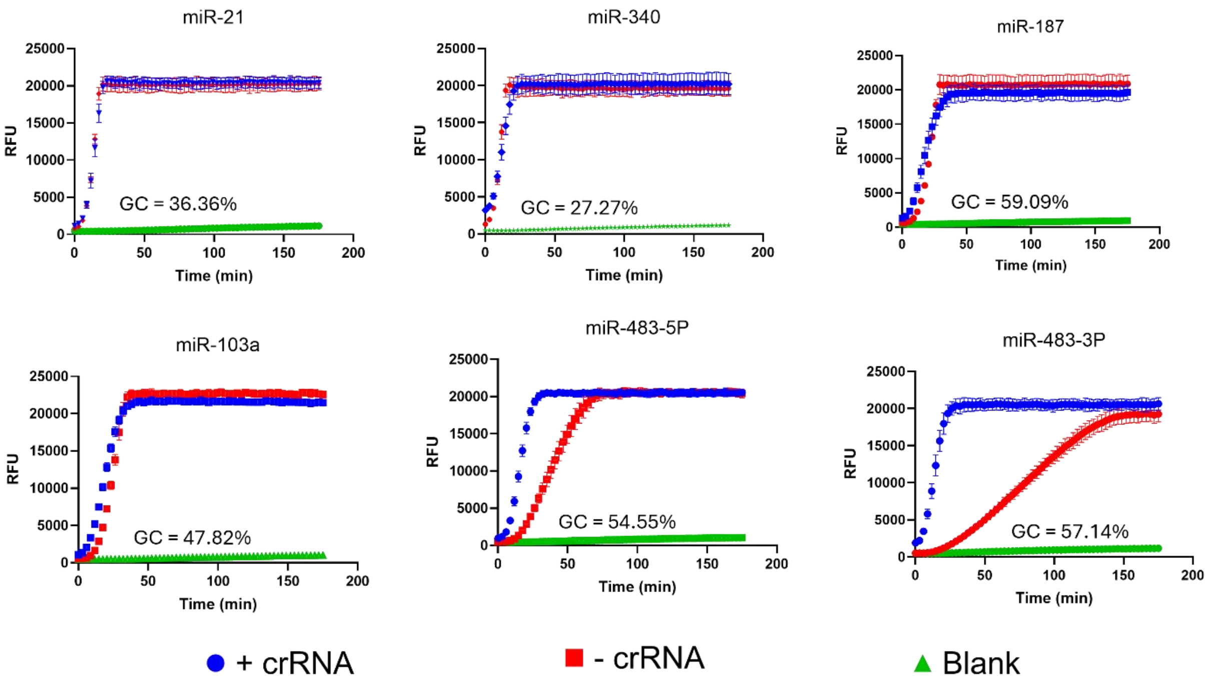

**Supplementary Fig. 6. Comparison of *trans* cleavage kinetics for representative miRNAs with and without crRNA**. Fluorescence time courses are shown for six miRNA targets (blue curves = +crRNA; red = −crRNA; green = no-target control). miR-21, miR-340, miR-187 and miR-103a exhibited comparable activation kinetics in both +crRNA and −crRNA conditions, demonstrating crRNA-independent performance. In contrast, the miR-483-5p and miR-483-3p showed noticeably slower kinetics in the absence of crRNA. All data are mean ± SD (n ≥ 3 technical replicates).

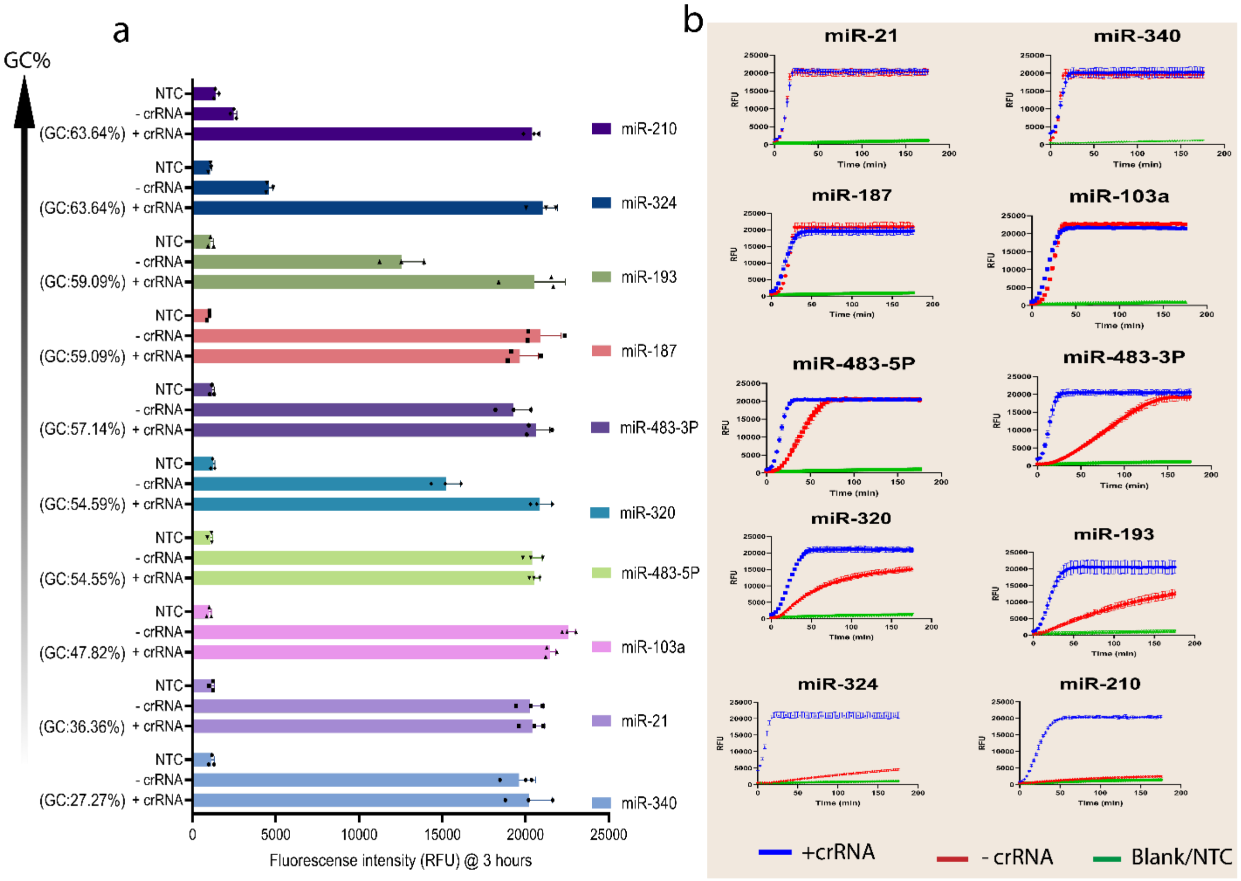

**Supplementary Fig. 7.** (a) Performance of the crRNA-independent CRISPR/Cas12a system across miRNAs with varying GC content. Data for the CRISPR/Cas12a system with crRNAs are shown for comparison. The crRNA-independent assay produces robust signals for low-GC miRNAs (e.g. miR-21, 36% GC; miR-340, 27% GC), comparable to the crRNA-driven system, whereas high-GC miRNAs (e.g. miR-210, 64%; miR-324, 64%) yield negligible signal. (b) Real-time *trans* cleavage kinetics for the ten representative miRNAs. Curves are shown in the presence (+crRNA, blue) or absence (–crRNA, red) of crRNA; green curves are no-target controls or blank. For some targets with GC content ≤60%, the +crRNA and –crRNA signals are similar. In contrast, above ~60% GC the activation signal declines to baseline (examples: miR-210, miR-324). The ten miRNAs tested (GC%) were miR-21 (36%), miR-340 (27%), miR-187 (59%), miR-103a (48%), miR-483-5p (55%), miR-483-3p (57%), miR-320 (55%), miR-193 (59%), miR-324 (64%) and miR-210 (64%). All data are mean ± SD (n ≥ 3 technical replicates). Assays were performed with AsCas12a and fluorescent reporter R1.

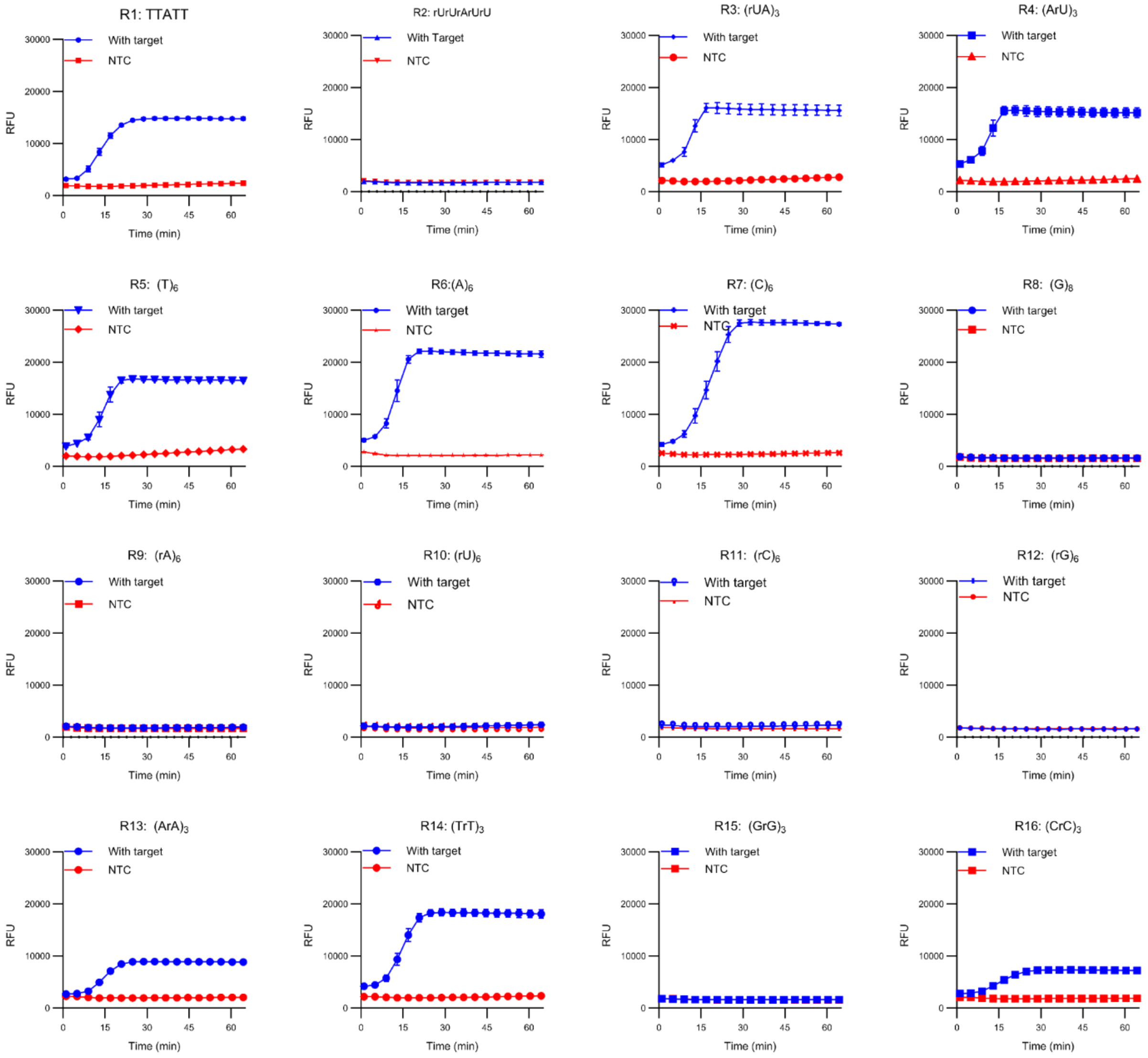

**Supplementary Fig. 8.** *Trans* cleavage kinetics of 16 substrate reporters with AsCas12a. The panel shows time courses for reporters R1–R16 (see **Table 9** for sequences). DNA-based reporters R1, R5–R7, R13–R14 and R16 were efficiently cleaved, albeit with some variation in kinetics. In contrast, RNA-only reporters (R9–R12) and G-rich reporters (R8, R12, R15) showed no detectable cleavage. This selectivity is consistent with Cas12a's known preference for DNA substrates; the lack of cleavage of G-rich reporters may be due to G-quadruplex formation quenching the fluorescent signal. All data are mean ± SD (n ≥ 3 technical replicates). Activation with target is denoted as (+), and without the target is represented as (-).

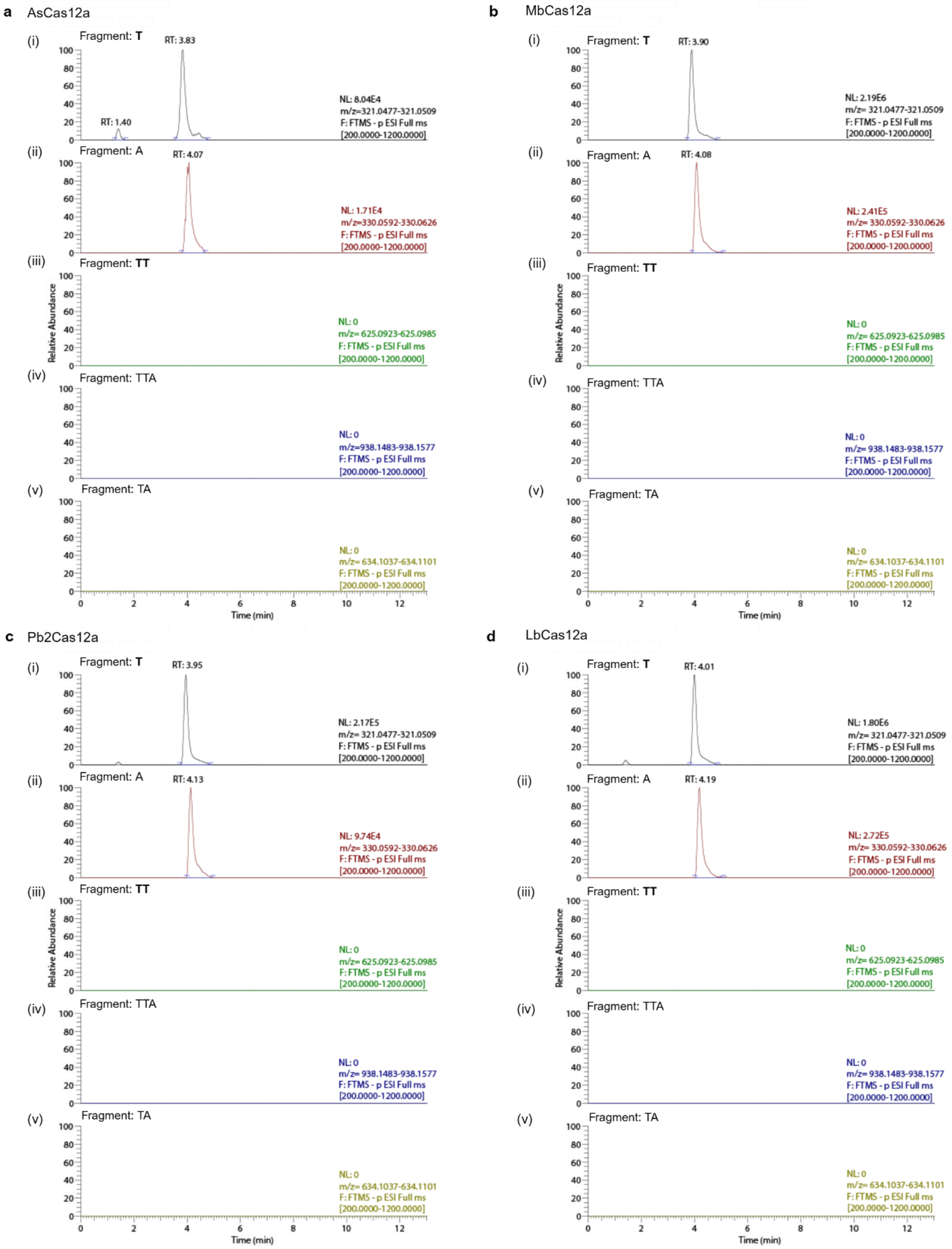

a AsCas12a
(i) Fragment: T
RT: 3.83
RT: 1.40
NL: 8.04E4
m/z=321.0477-321.0509
F: FTMS - p ESI Full ms
[200.0000-1200.0000]
(ii) Fragment: A
RT: 4.07
NL: 1.71E4
m/z=330.0592-330.0626
F: FTMS - p ESI Full ms
[200.0000-1200.0000]
(iii) Fragment: TT
NL: 0
m/z= 625.0923-625.0985
F: FTMS - p ESI Full ms
[200.0000-1200.0000]
(iv) Fragment: TTA
NL: 0
m/z=938.1483-938.1577
F: FTMS - p ESI Full ms
[200.0000-1200.0000]
(v) Fragment: TA
NL: 0
m/z= 634.1037-634.1101
F: FTMS - p ESI Full ms
[200.0000-1200.0000]
Relative Abundance
Time (min)
b MbCas12a
(i) Fragment: T
RT: 3.90
NL: 2.19E6
m/z= 321.0477-321.0509
F: FTMS - p ESI Full ms
[200.0000-1200.0000]
(ii) Fragment: A
RT: 4.08
NL: 2.41E5
m/z= 330.0592-330.0626
F: FTMS - p ESI Full ms
[200.0000-1200.0000]
(iii) Fragment: TT
NL: 0
m/z= 625.0923-625.0985
F: FTMS - p ESI Full ms
[200.0000-1200.0000]
(iv) Fragment: TTA
NL: 0
m/z= 938.1483-938.1577
F: FTMS - p ESI Full ms
[200.0000-1200.0000]
(v) Fragment: TA
NL: 0
m/z= 634.1037-634.1101
F: FTMS - p ESI Full ms
[200.0000-1200.0000]
Relative Abundance
Time (min)
c Pb2Cas12a
(i) Fragment: T
RT: 3.95
NL: 2.17E5
m/z=321.0477-321.0509
F: FTMS - p ESI Full ms
[200.0000-1200.0000]
(ii) Fragment: A
RT: 4.13
NL: 9.74E4
m/z= 330.0592-330.0626
F: FTMS - p ESI Full ms
[200.0000-1200.0000]
(iii) Fragment: TT
NL: 0
m/z= 625.0923-625.0985
F: FTMS - p ESI Full ms
[200.0000-1200.0000]
(iv) Fragment: TTA
NL: 0
m/z= 938.1483-938.1577
F: FTMS - p ESI Full ms
[200.0000-1200.0000]
(v) Fragment: TA
NL: 0
m/z= 634.1037-634.1101
F: FTMS - p ESI Full ms
[200.0000-1200.0000]
Relative Abundance
Time (min)
d LbCas12a
(i) Fragment: T
RT: 4.01
NL: 1.80E6
m/z= 321.0477-321.0509
F: FTMS - p ESI Full ms
[200.0000-1200.0000]
(ii) Fragment: A
RT: 4.19
NL: 2.72E5
m/z= 330.0592-330.0626
F: FTMS - p ESI Full ms
[200.0000-1200.0000]
(iii) Fragment: TT
NL: 0
m/z= 625.0923-625.0985
F: FTMS - p ESI Full ms
[200.0000-1200.0000]
(iv) Fragment: TTA
NL: 0
m/z= 938.1483-938.1577
F: FTMS - p ESI Full ms
[200.0000-1200.0000]
(v) Fragment: TA
NL: 0
m/z= 634.1037-634.1101
F: FTMS - p ESI Full ms
[200.0000-1200.0000]
Relative Abundance
Time (min)

**Supplementary Fig. S9. Electrospray ionization mass spectrometry (ESI–MS) analysis of crRNA-independent Cas12a *trans* cleavage products generated from the canonical reporter R1 (TTATT). (a)** AsCas12a-treated samples revealed two dominant peaks at *m/z* 321.0 and 330.0, corresponding to thymidine monophosphate (T; $C_{10}H_{15}N_2O_8P$) **(i)** and adenosine monophosphate (A; $C_{10}H_{14}N_5O_6P$) **(ii)**, respectively, detected at retention times (RT) of 3.83 s and 4.07 s. Expected higher-order cleavage intermediates, including TT ($C_{20}H_{28}N_4O_{15}P_2$) **(iii)**, TTA ($C_{30}H_{40}N_9O_{20}P_3$) **(iv)**, and TA ($C_{20}H_{27}N_7O_{13}P_2$) **(v)**, were not detected, indicating near-complete degradation of the reporter into mononucleotides. Similar fragmentation profiles were observed for MbCas12a **(b)**, Pb2Cas12a **(c)**, and LbCas12a **(d)**, where only mononucleotide products were detected. Graph annotations: NL, normalized intensity; RT, retention time; *m/z*, mass-to-charge ratio; FTMS, Fourier transform mass spectrometry. Peak tailing likely arises from phosphate-associated ionization effects and buffer components.

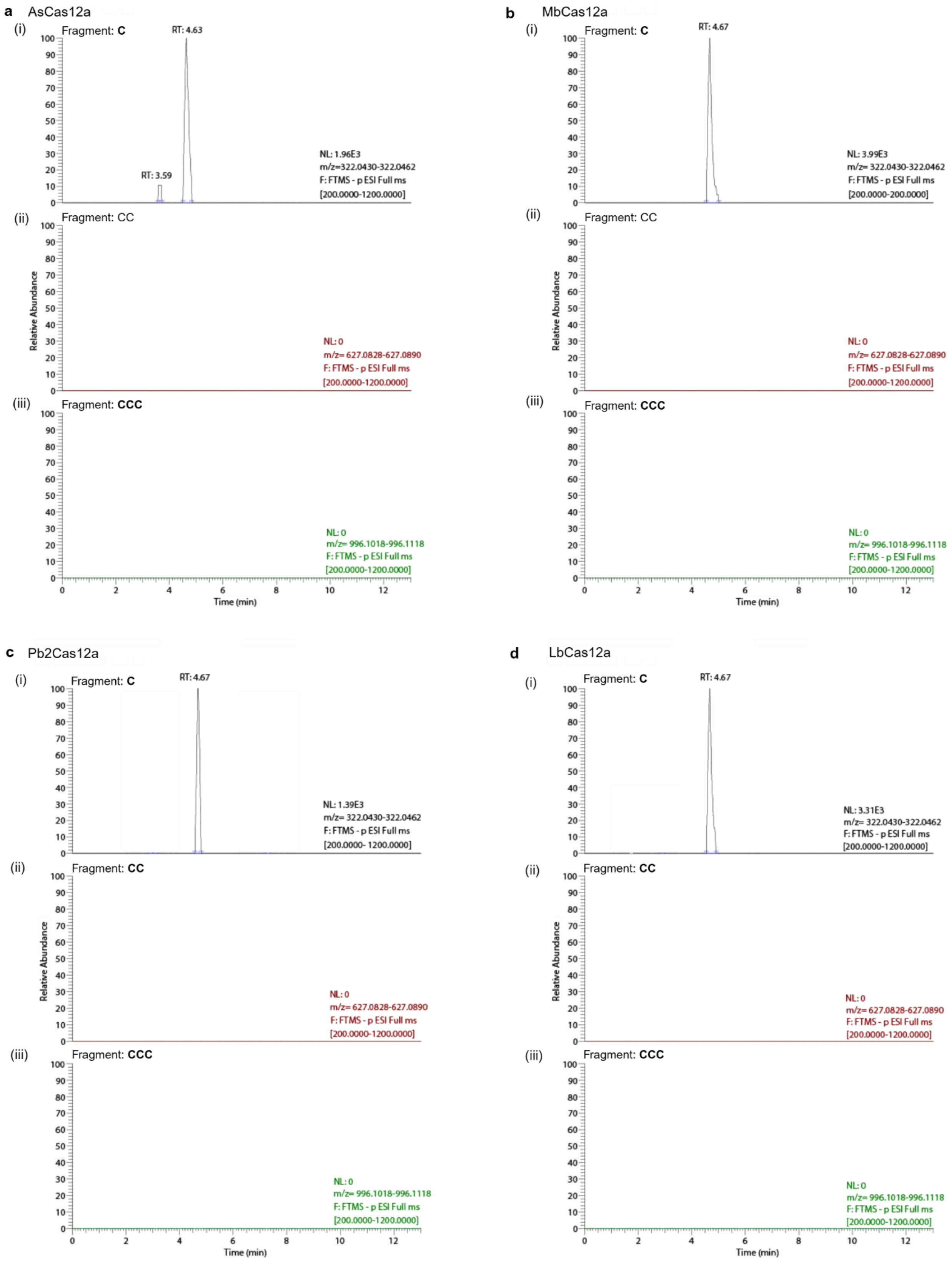

a AsCas12a
(i) Fragment: C
RT: 4.63
RT: 3.59
NL: 1.96E3
m/z=322.0430-322.0462
F: FTMS - p ESI Full ms
[200.0000-1200.0000]
(ii) Fragment: CC
NL: 0
m/z= 627.0828-627.0890
F: FTMS - p ESI Full ms
[200.0000-1200.0000]
(iii) Fragment: CCC
NL: 0
m/z= 996.1018-996.1118
F: FTMS - p ESI Full ms
[200.0000-1200.0000]
Relative Abundance
Time (min)
b MbCas12a
(i) Fragment: C
RT: 4.67
NL: 3.99E3
m/z= 322.0430-322.0462
F: FTMS - p ESI Full ms
[200.0000-200.0000]
(ii) Fragment: CC
NL: 0
m/z= 627.0828-627.0890
F: FTMS - p ESI Full ms
[200.0000-1200.0000]
(iii) Fragment: CCC
NL: 0
m/z= 996.1018-996.1118
F: FTMS - p ESI Full ms
[200.0000-1200.0000]
Relative Abundance
Time (min)
c Pb2Cas12a
(i) Fragment: C
RT: 4.67
NL: 1.39E3
m/z= 322.0430-322.0462
F: FTMS - p ESI Full ms
[200.0000- 1200.0000]
(ii) Fragment: CC
NL: 0
m/z= 627.0828-627.0890
F: FTMS - p ESI Full ms
[200.0000-1200.0000]
(iii) Fragment: CCC
NL: 0
m/z= 996.1018-996.1118
F: FTMS - p ESI Full ms
[200.0000-1200.0000]
Relative Abundance
Time (min)
d LbCas12a
(i) Fragment: C
RT: 4.67
NL: 3.31E3
m/z= 322.0430-322.0462
F: FTMS - p ESI Full ms
[200.0000-1200.0000]
(ii) Fragment: CC
NL: 0
m/z= 627.0828-627.0890
F: FTMS - p ESI Full ms
[200.0000-1200.0000]
(iii) Fragment: CCC
NL: 0
m/z= 996.1018-996.1118
F: FTMS - p ESI Full ms
[200.0000-1200.0000]
Relative Abundance
Time (min)

**Supplementary Fig. S10. ESI–MS analysis of crRNA-independent Cas12a *trans* cleavage products generated from reporter 7 (R7: CCCCCC). (a)** AsCas12a-treated samples exhibited a single dominant peak corresponding to cytidine monophosphate (C; $C_9H_{14}N_3O_8P$) at *m/z* 322.0446 and RT 4.63 s **(i)**. Expected oligomeric intermediates, including CC ($C_{18}H_{26}N_6O_{15}P_2$) at *m/z* 627.0859 **(ii)** and CCC ($C_{27}H_{38}N_9O_{26}P_3$) at *m/z* 996.1068 **(iii)**, were not detected, consistent with extensive reporter degradation into mononucleotide products. Comparable cleavage patterns were observed for MbCas12a **(b)**, Pb2Cas12a **(c)**, and LbCas12a **(d)**, each yielding predominantly mononucleotide species. Graph annotations: NL, normalized intensity; RT, retention time; *m/z*, mass-to-charge ratio; FTMS, Fourier transform mass spectrometry.

**Note**

To further validate that crRNA-independent Cas12a fluorescence outputs reflect bona fide Cas12a-mediated reporter hydrolysis, we analysed post-reaction mixtures by Electrospray ionization mass spectrometry (ESI-MS) for two representative reporters, the traditional Cas12 reporter (R1: **TTATT**) and C-rich reporter (R7: **CCCCCC**) (**Suppl Fig. 9,10, Table S10**) across four Cas12a orthologs (AsCas12a, MbCas12a, Pb2Cas12a, and LbCas12a). For the canonical reporter R1, the results yielded smooth, high-intensity peaks corresponding to only single nucleotide products as follows: AsCas12a shows m/z 321.049 (T; RT 3.83 s; $8.04\times10^4$) and m/z 330.061 (A; RT 4.07 s; $1.71\times10^4$); MbCas12a shows peaks corresponding to: m/z 321.049 (T; RT 3.9 s; $2.19\times10^6$) and m/z 330.061 (A; RT 4.08 s; $2.41\times10^5$); Pb2 yielded: m/z 321.049 (T; RT 3.9 s; $2.17\times10^5$) and m/z 330.061 (A; RT 4.13 s; $9.74\times10^4$); and LbCas12a shows: m/z 321.049 (T; RT 4.01 s; $1.80\times10^6$) and m/z 330.061 (A; RT 4.19 s; $2.72\times10^5$); whereas longer candidate intermediates (TA, TT, and TTA) were not detected under these conditions, suggesting spontaneous hydrolysis of the R1 reporter substrate. Similarly, the C-rich DNA reporter R7 (5′-C-C-C-C-C-C-3′) showed an analogous, enzyme-dependent fragmentation profile (**Suppl Fig. 10**), with detected products limited to C-mononucleotide (m/z 322.0446) across all four orthologs, which also suggests a spontaneous substrate hydrolysis (see summary in **Table 10**).

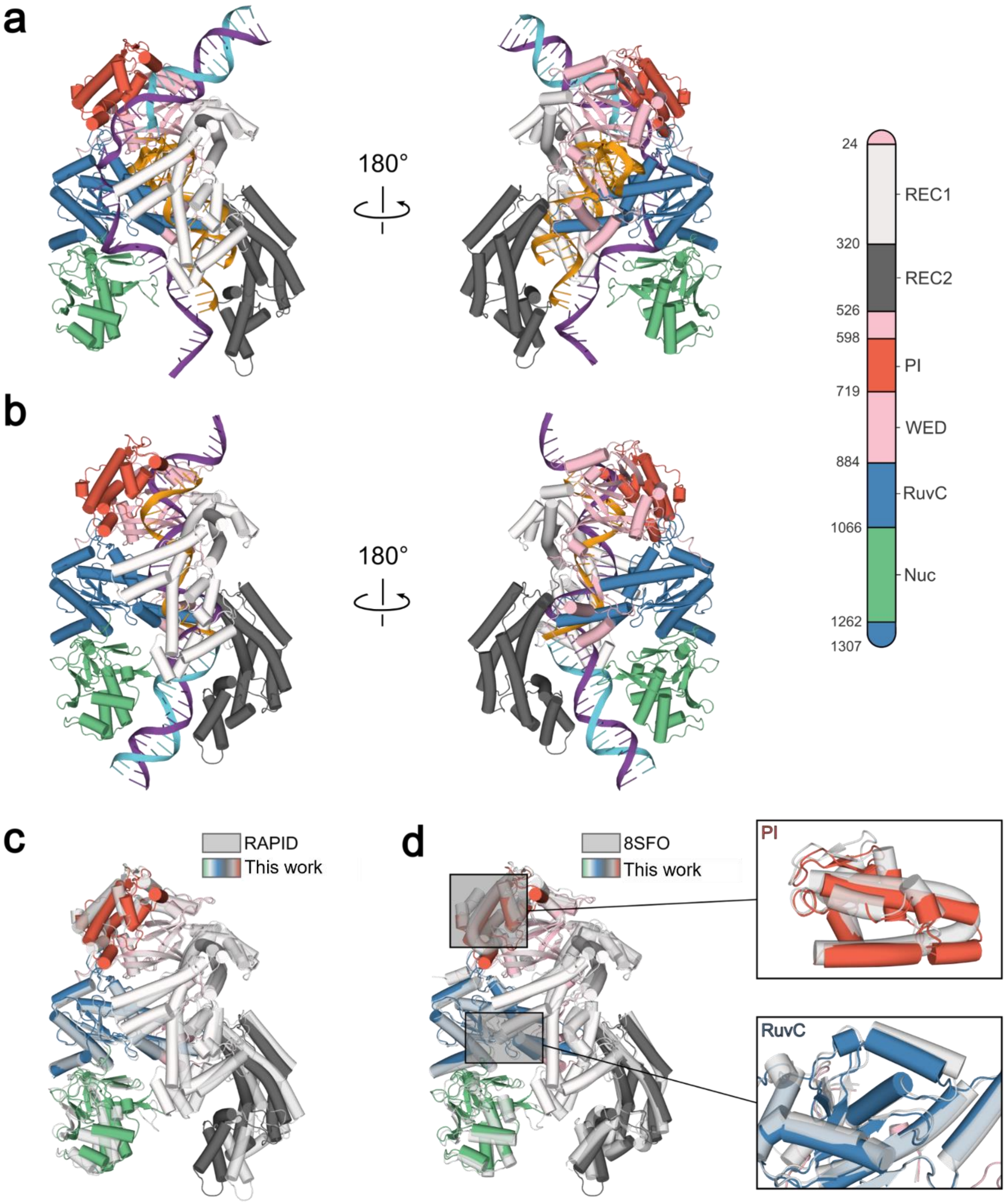

**Supplementary Fig. 11.** Boltz-2–predicted structural complexes of AsCas12a with distinct nucleic acid configurations. **(a)** crRNA-directed configuration consisting of AsCas12a in complex with crRNA, miniCaR, and a partial DNA duplex (or RAPID[12]). **(b)** crRNA-independent configuration generated by removal of the crRNA to visualize direct miniCaR engagement with AsCas12a. In this state, the miniCaR occupies the canonical crRNA-binding channel, revealing an alternative nucleic acid-binding architecture. **(c)** Structural superimposition of the crRNA-bound complex shown in **(a)** and the crRNA-independent complex shown in **(b)** to identify conformational rearrangements associated with guide-free operation. **(d)** Comparative structural analysis revealed subtle conformational shifts with Boltz-2 and pronounced shift with cryo-EM within the PI and RuvC domains, whereas the remaining domains remained largely conserved. The miniCaR sequence used in this analysis corresponds to miRNA-21.

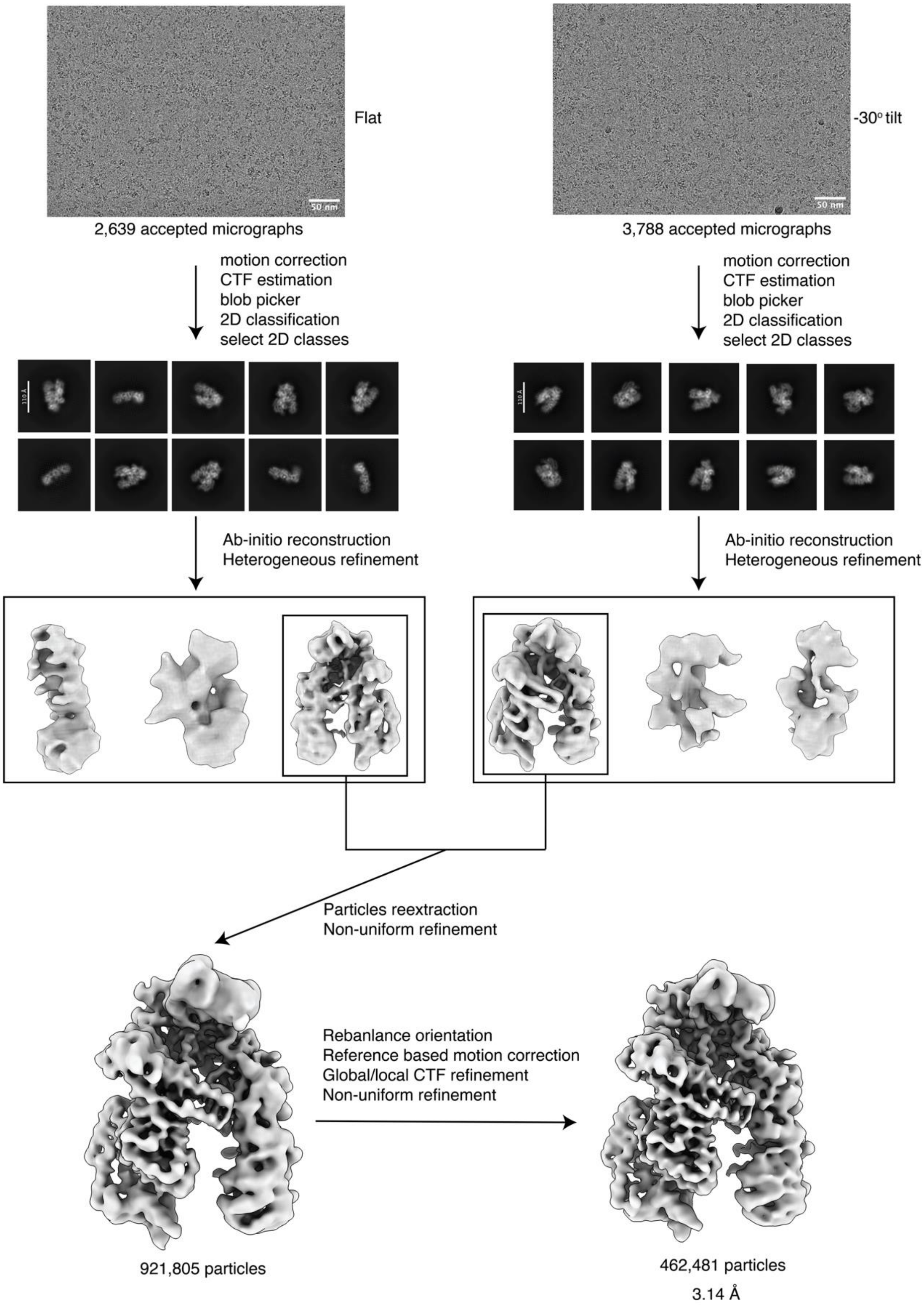

**Supplementary Fig. 12. Cryo-EM data processing workflow for crRNA-independent platform.** Cryo-EM datasets collected without tilting and at -30° stage tilt were processed individually. The particles of the best-resolved class after ab-initio reconstruction and heterogeneous refinements were combined for further processing. See Method section for details.

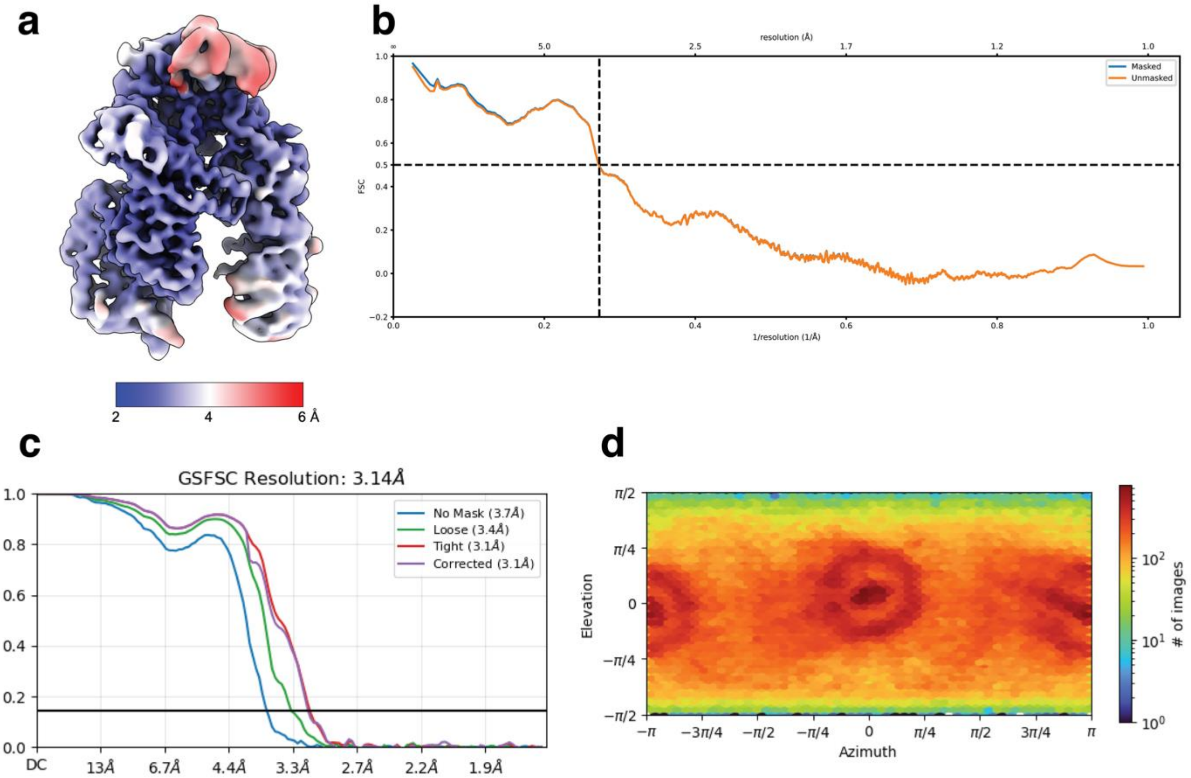

**Supplementary Fig. 13. Cryo-EM data analysis of crRNA. a**. Local resolution map. **b**. Map-to-model FSC curve. **c**. Gold-standard Fourier Shell Correlation (FSC) curve with resolution reported at FSC = 0.143. **d**. Euler angle distribution plot of particle orientation.

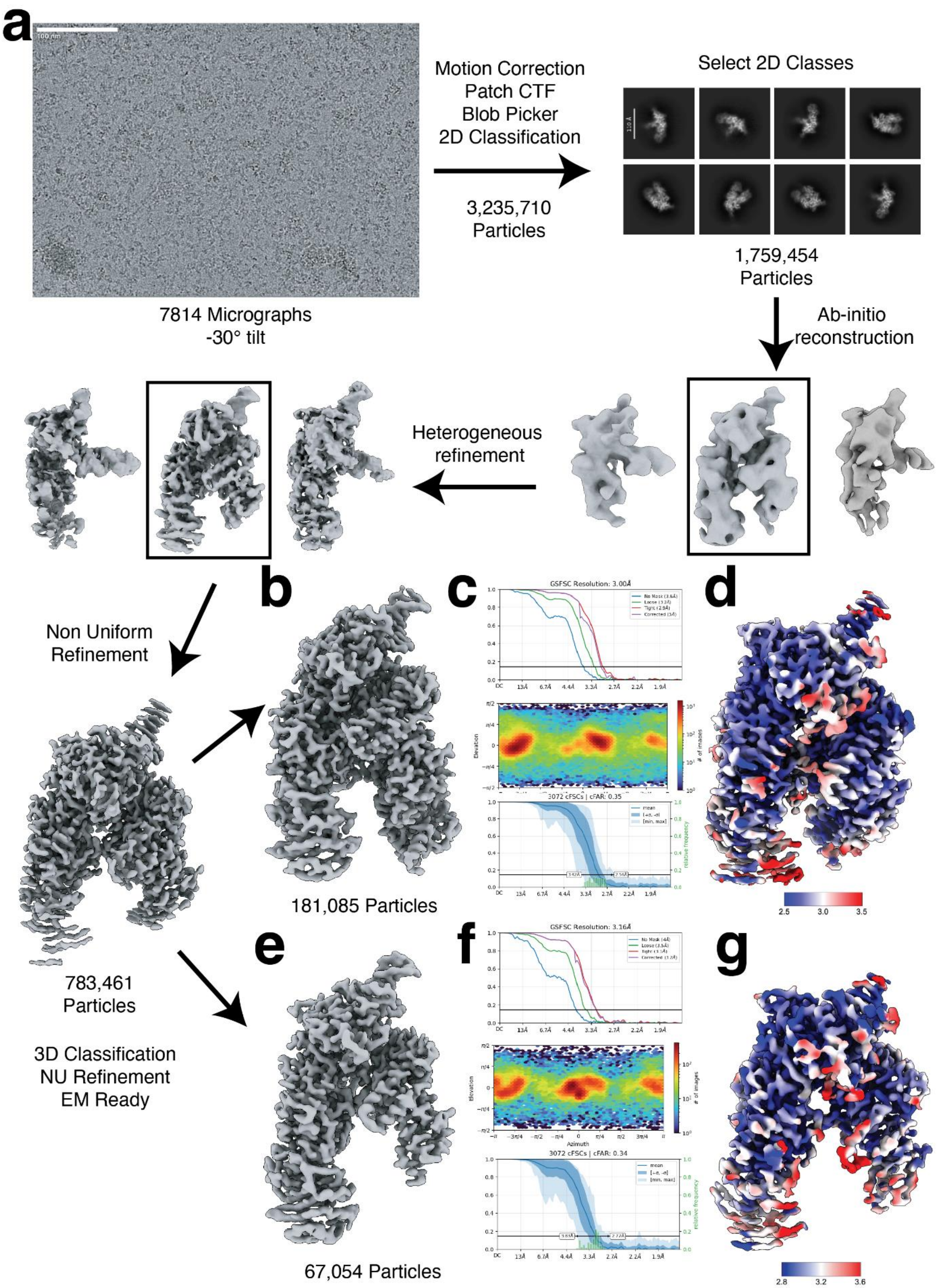
a
100 nm
7814 Micrographs
-30° tilt
Motion Correction
Patch CTF
Blob Picker
2D Classification
3,235,710
Particles
Select 2D Classes
110 Å
1,759,454
Particles
Ab-initio
reconstruction
Heterogeneous
refinement
Non Uniform
Refinement
783,461
Particles
3D Classification
NU Refinement
EM Ready
b
181,085 Particles
c
GSFSC Resolution: 3.00Å
d
2.5
3.0
3.5
e
67,054 Particles
f
GSFSC Resolution: 3.16Å
g
2.8
3.2
3.6

**Supplementary Figure 14: Cryo-EM processing with crRNA inclusion (or RAPID[12]). (a)** Cryo-EM processing pipeline for both RAPID structures **b,** Sharpened map of AsCas12a 16bp target bound complex. (**c)** from top to bottom, Fourier Shell Correlation (FSC) diagram of the 16bp target bound complex utilizing a similarity threshold of 0.143, orientation distribution map of the particles used for the final reconstruction, and conical FSC plot (cFSC) representing directional resolution of the map. (**d)** Reported local resolution for the 16bp target bound structure. (**e)** Sharpened map of AsCas12a 5bp target bound complex. (**f)** From top to bottom, FSC diagram of the 5bp target bound complex utilizing a similarity threshold of 0.143, orientation distribution map of the particles used for the final reconstruction, and conical FSC plot (cFSC) representing directional resolution of the map. (**g)** Reported local resolution for the 5bp target bound structure.

**Note**
To test how Cas12a can target ssDNA or ssRNA indiscriminately with a partial PAM-containing dsDNA probe, we solved a structure of AsCas12a bound to a gRNA and a synthetic partial duplexed DNA containing a 4nt duplex downstream of the PAM and an extended ssDNA region at the PAM strand to imitate the presence of the NTS (**Extended data Fig. 4,** otherwise known as RAPID[12]). Cryo-EM structures of this partial duplex in the presence of a ssRNA target resolved two RAPID states: a partial target state containing the duplex probe with the gRNA spacer bound to the first four DNA nucleotides and five target RNA nucleotides, and a fully target-engaged state containing the full nucleic-acid assembly (**Extended data Fig. 4a**). The partial target state most closely resembles a 9bp R-loop intermediate represented by PDB 8SFJ (**Extended data Fig. 4b**), with incomplete ordering of the bridge helix and weak density at the PAM-distal nucleic-acid segment. By contrast, the target-engaged state closely resembles the canonical 20-bp R-loop architecture (PDB 8SFO), showing completion of the bridge helix, continuous density for the ~20-bp hybrid duplex, and clear ordering of REC2, consistent with full PAM-distal engagement (**Extended data Fig. 4c-e**). Notably, however, density for the RuvC lid and the NTS was not present, indicating that extensive nucleic-acid engagement can be achieved without rigidification of all catalytic elements and maturation of the RuvC lid. Together, these structures place our guide-free platform and gRNA-containing CRISPR/Cas12a (RAPID)[12] in distinct regions of the Cas12a conformational landscape.

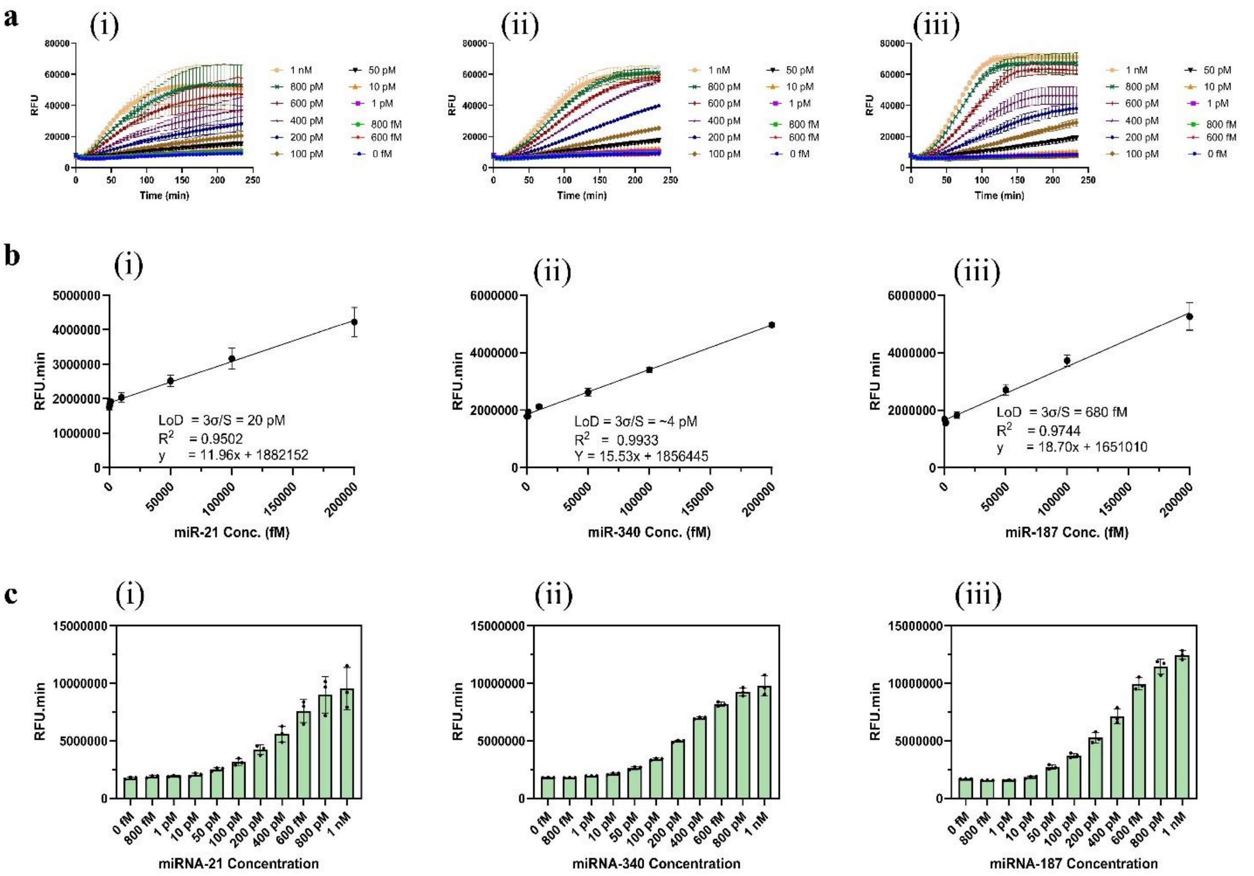

**Supplementary Fig. 15.** Sensitivity of the crRNA platform for direct, amplification-free miRNA detection. (a) Real-time fluorescence kinetics of three representative miRNAs: miR-21 (i), miR-340 (ii), and miR-187 (iii), across a serial dilution range from 1 nM to 600 fM. (b) Linear regression analysis of endpoint fluorescence intensity versus input concentration for the same three miRNAs, enabling determination of analytical performance. Calculated LoD, based on the equation LoD = 3σ/S, are 20.3 pM for miR-21 (slope S = 11.96, σ = 80803.23 fM), 3.8 pM for miR-340 (S = 15.53, σ = 19497.19 fM), and 0.68 pM for miR-187 (S = 18.7, σ = 4245.67 fM). (c) Endpoint fluorescence bar plots for miR-21 (i), miR-340 (ii), and miR-187 (iii), showing robust signal generation across the tested range, with dynamic responses spanning six orders of magnitude in input concentration. All data are presented as mean ± SD for n ≥ 3 technical replicates.

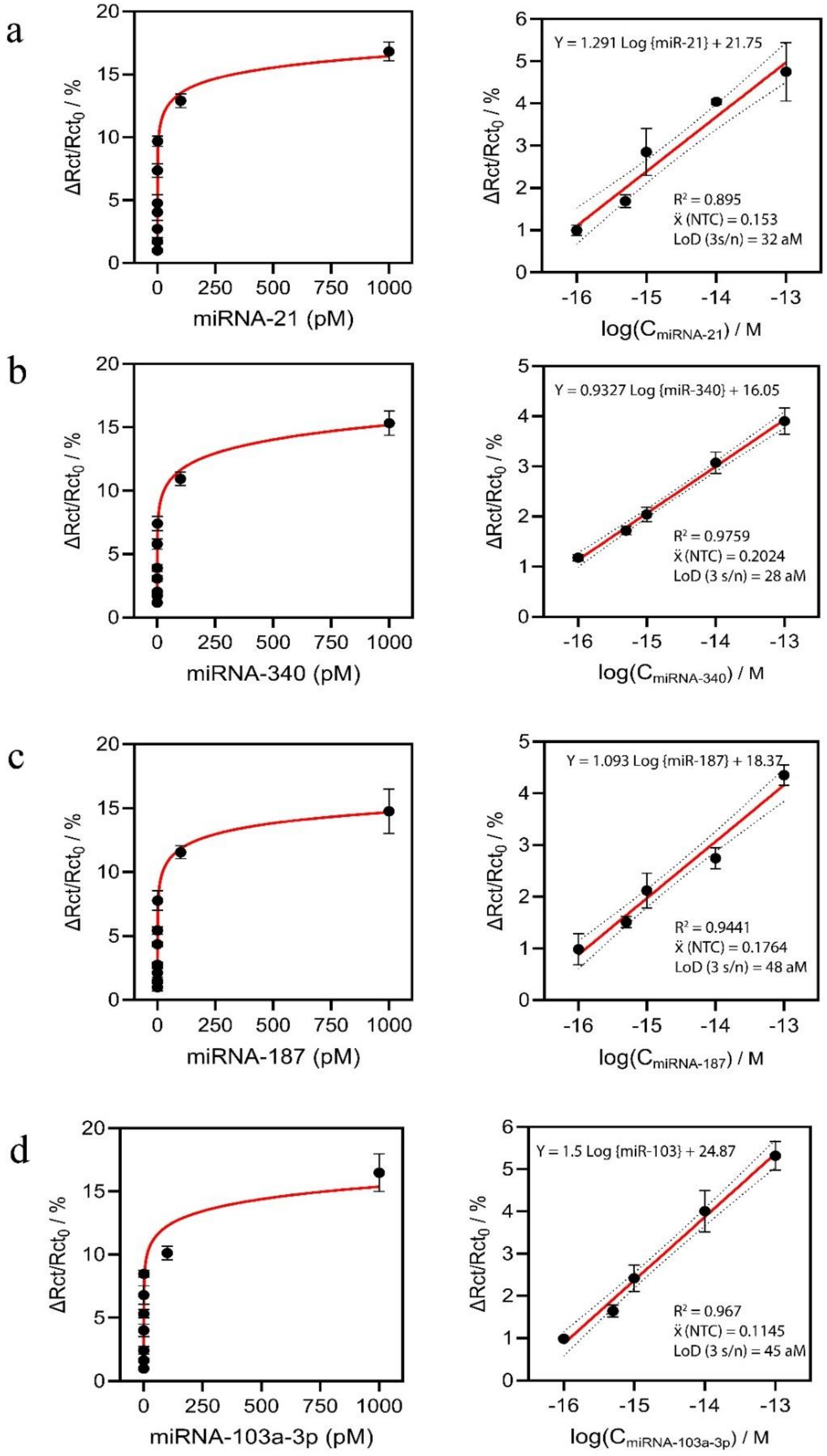

**Supplementary Fig. 16.** Dependence of resistance to charge-transfer (Rct) reduction with miRNA target concentration over the range of 100 aM to 1 nM. Calibration curves showing a linear relationship between the percentage of Rct reduction and the logarithm of miRNA concentration ($R^2 \geq 0.90$) for **(a)** miRNA-21,

**(b)** miRNA-340, **(c)** miRNA-187 and **(d)** miRNA-103a-3p. Data are shown as mean ± s.d. (n ≥ 3 independent replicates).

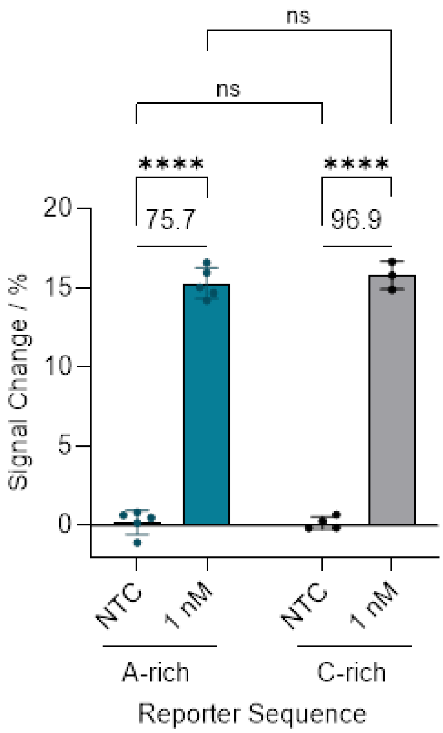

**Supplementary Fig. 17.** Fold change between the non-targeting control (NTC) and 1 nM miRNA-340, tested against A- and C- rich reporters with values indicated above each data pair. Unless otherwise stated, experiments were performed using 150 nM Cas12a, 40 nM duplex and 150 nM gRNA. Data are shown as mean ± s.d. (n ≥ 3 independent replicates). Statistical significance was analyzed using ANOVA test: where ns = not significant with $\rho > 0.05$, and the asterisk (****) denotes significant differences with $\rho < 0.0001$.

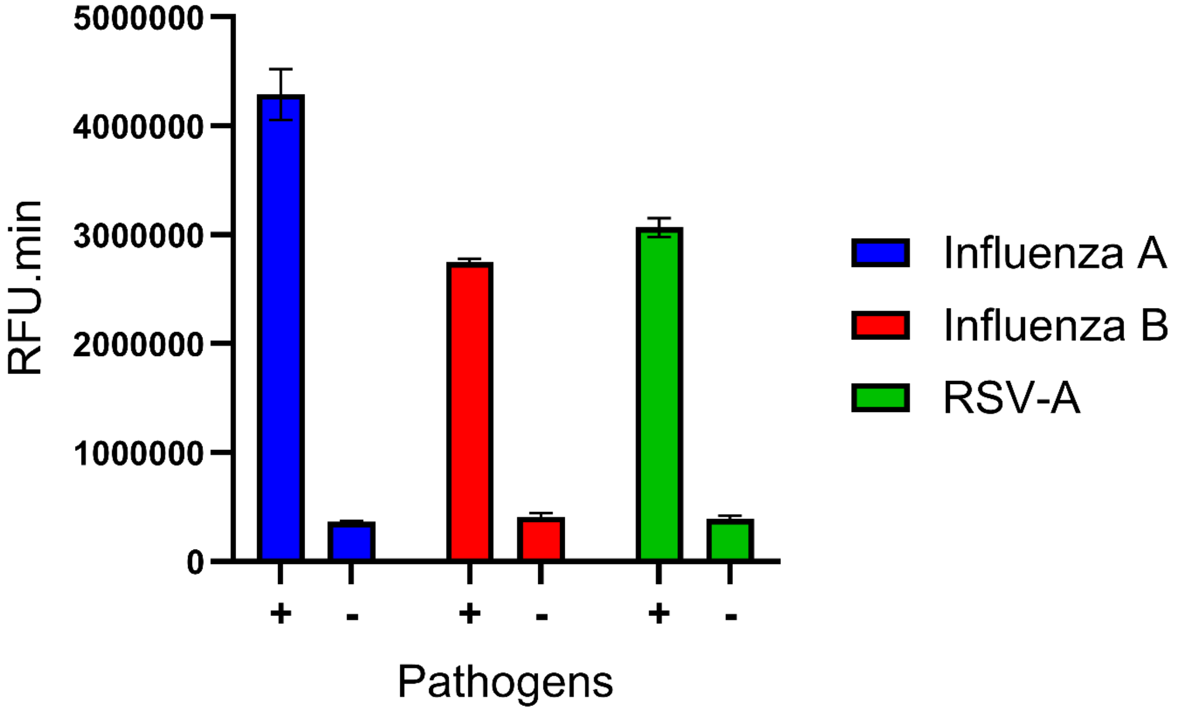

**Supplementary Fig. 18.** Preliminary testing of COMPANION-RT-LAMP on synthetic DNA targets. COMPANION-RT-LAMP was initially tested using synthetic DNA fragments corresponding to Influenza A, Influenza B, and RSV A to mimic the RT-LAMP amplicons for these viruses. The assay produced a strong fluorescence signal in the presence of each target (+), whereas no signal was observed in the negative control reactions (–). Reactions were carried out at 37 °C for 60 min. Data are presented as mean ± SD (n = 3 technical replicates).

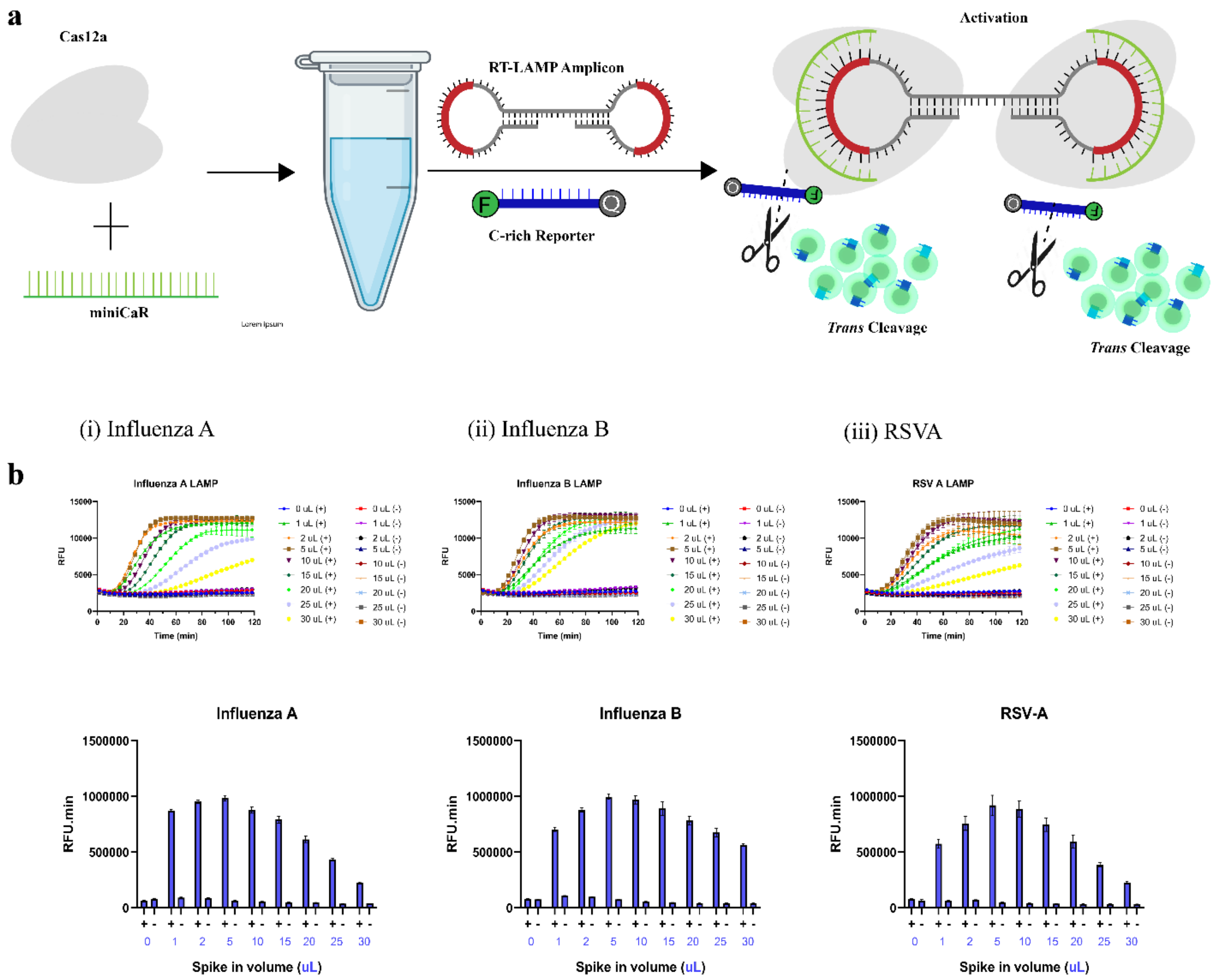

**Supplementary Fig. 19.** COMPANION-RT-LAMP workflow and LAMP input volume optimization. (a) Schematic of the COMPANION-RT-LAMP workflow and mechanism. The Cas12a–cRNA complex binds to the loop region of the LAMP-generated dumbbell DNA structure, triggering trans-cleavage of the reporter molecule in the presence of Cas12a. (b) Optimization of the RT-LAMP product volume added to the COMPANION detection reaction for Influenza A (H1N1), Influenza B, and RSV A. Real-time fluorescence curves and 60 min endpoint readings are shown for reactions containing 1, 2, 5, 10, 15, 20, 25, and 30 µL of the RT-LAMP reaction. All three targets produced the highest signal when 5 µL of LAMP product was used, which was chosen for subsequent experiments. The symbols “+” and “–” denote positive and negative controls, respectively. Reactions were performed at 37 °C and monitored in real time; bar graphs show the fluorescence intensity at 60 min. Data are mean ± SD (n = 3 technical replicates).

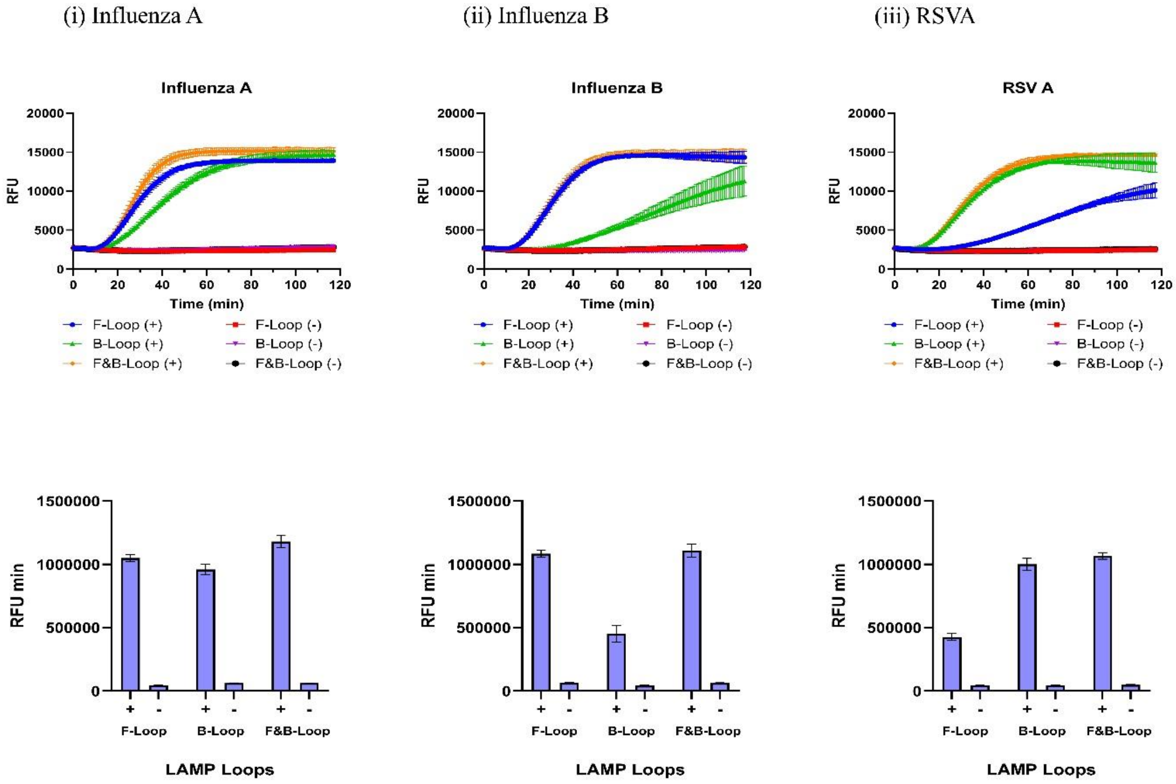

**Supplementary Fig. 20.** Comparison of different LAMP loop targets in COMPANION-RT-LAMP. Fluorescence kinetics and endpoint intensities for COMPANION-RT-LAMP assays targeting different loop regions of the LAMP amplicon. The complementary RNA (cRNA) was designed to bind either the forward loop (F-loop), the backward loop (B-loop), or both loops (F + B) of the dumbbell structure. Assays were conducted for Influenza A (H1N1) **(i)**, Influenza B **(ii)**, and RSV A **(iii)**. For each virus, real-time fluorescence curves and 60 min endpoint bar plots are shown, demonstrating that targeting both loops yields the strongest signal, followed by targeting the single loops. "+" and "–" indicate positive and negative controls, respectively. Reactions were run at 37 °C with real-time fluorescence monitoring, and endpoint measurements were taken at 60 min. Data are mean ± SD (n = 3 technical replicates).

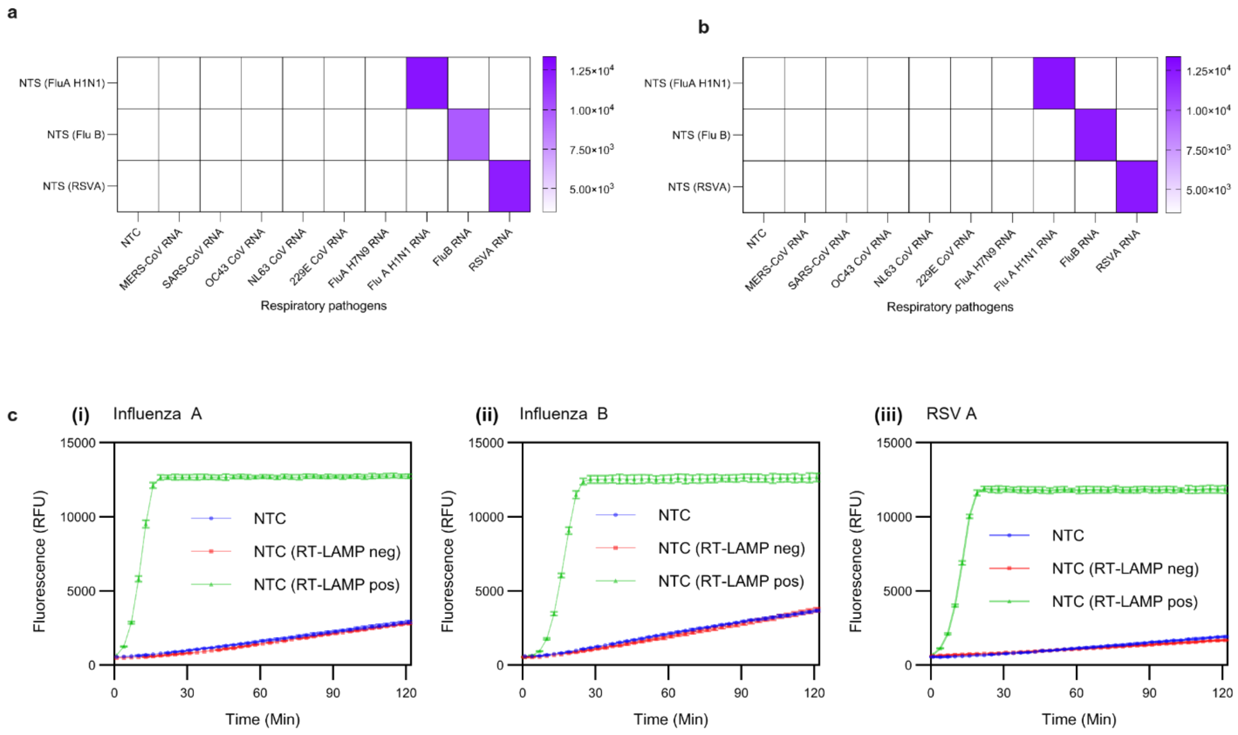

**Supplementary Fig. 21.** Specificity heatmaps and control experiments for COMPANION-RT-LAMP. (a, b) Heatmaps demonstrating the specificity of COMPANION-RT-LAMP for Influenza A (H1N1), Influenza B, and RSV A after (a) 30 min and (b) 120 min of reaction. Each heatmap includes the target virus and non-target respiratory viruses (MERS-CoV, SARS-CoV, HCoV-OC43, HCoV-NL63, HCoV-229E, and Influenza A (H7N1)). Specific fluorescence signals appear only for the intended targets at 30 min, and even after 2 h of reaction no substantial off-target signals are observed. (c) Control reactions for the COMPANION-RT-LAMP assay prior to testing patient samples. No-template controls (NTCs), as well as positive and negative control reactions for Influenza A (H1N1) **(i)**, Influenza B **(ii)**, and RSV A **(iii)**, show that the reporter remains quenched in the absence of target and is only cleaved in the presence of the correct target amplicon. All reactions were performed at 37 °C. Data are representative of n = 3 technical replicates.

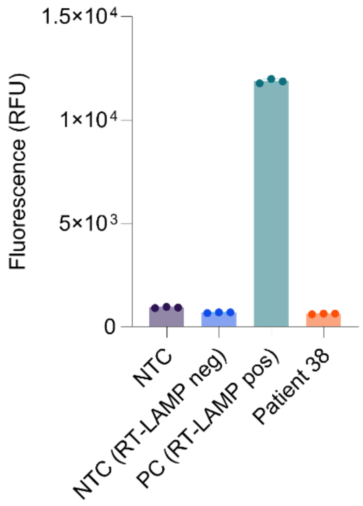

**Supplementary Fig. 22.** COMPANION assay on a low-viral-load influenza B sample. Patient sample #38 had a high RT-PCR cycle threshold (>33), indicating a low viral load. In the COMPANION assay, this sample produced no detectable *trans* cleavage signal, consistent with being near the detection limit. Negative and positive RT-LAMP controls were included for comparison. All data are mean ± SD (n ≥ 3 technical replicates).

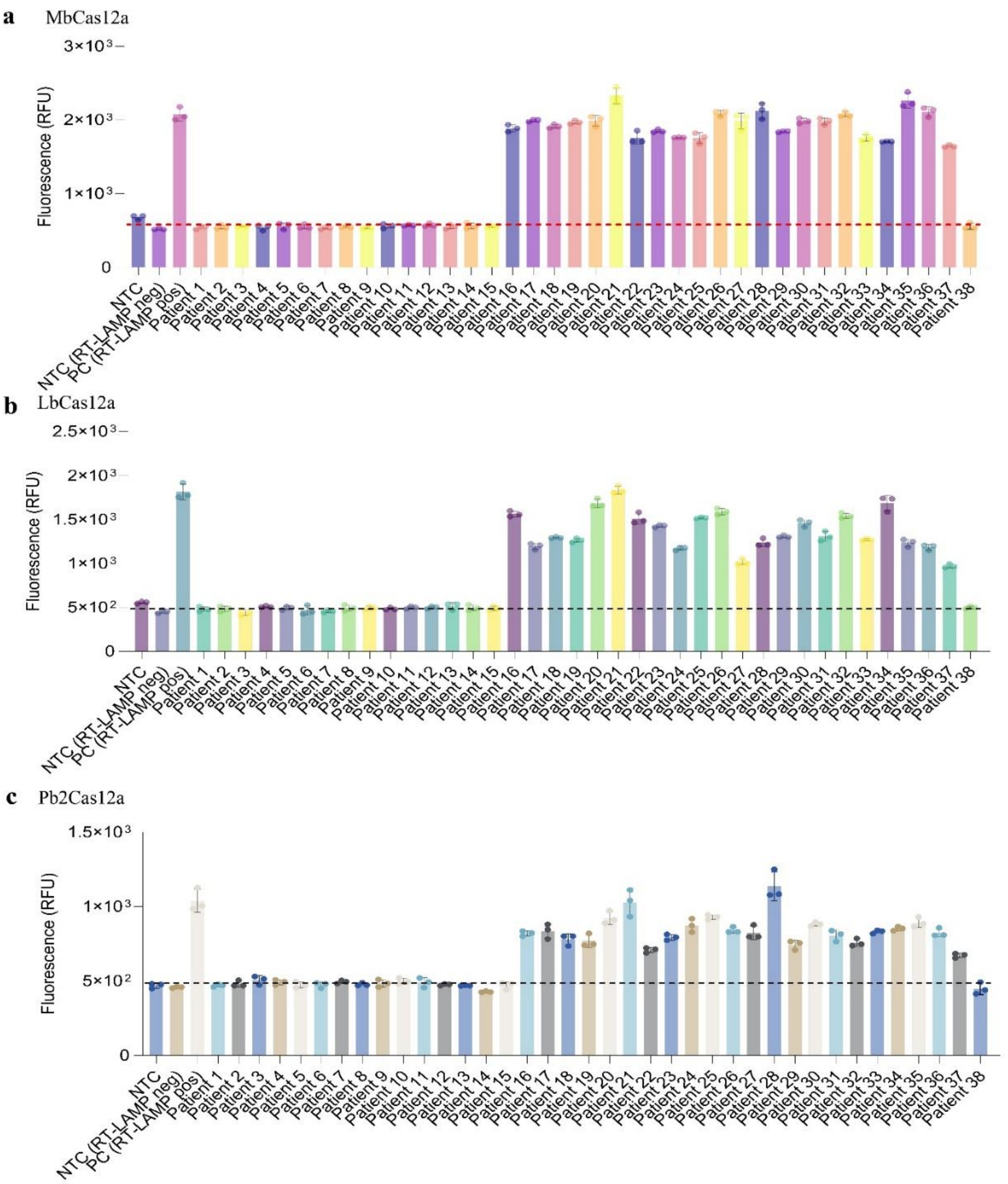

**Supplementary Fig. 23.** Patient sample detection using alternative Cas12a orthologs.COMPANION-RT-LAMP detection of patient samples for Influenza A, Influenza B, and RSV A using three Cas12a orthologs: (a) MbCas12a; (b) LbCas12a; and (c) Pb2Cas12a. Fluorescence readings at 30 min clearly distinguish the positive and negative patient samples for all three viruses, comparable to the results obtained with AsCas12a (Figure 7). All true positive samples generated strong fluorescence signals, whereas no fluorescence was observed in true negative samples. Data are presented as mean ± SD (n = 3 technical replicates).

#### General note

**Electrochemistry assay with COMPANION.**

To expand the COMPANION platform beyond fluorescence readouts, we implemented an amplification- and label-free electrochemical detection scheme using electrochemical impedance spectroscopy - EIS (**Fig. 5a**). In this assay, a thiolated ssDNA (SH-ssDNA) reporter is self-assembled on a gold screen-printed electrode surface, forming a passivating layer. Upon target recognition by the Cas12a–duplex-target complex, the *trans*-cleavage activity of Cas12a indiscriminately cuts the surface-bound DNA reporters, thus removing the insulating layer and enhancing electron transfer at the electrode.[16] This target-triggered process decreases the interfacial charge-transfer resistance ($R_{ct}$) and produces an impedance signal change proportional to the miRNA/target concentration. We first verified that the electrochemical system exhibits negligible background signals under stringent controls with A-rich and C-rich reporters (**Fig. 5b, Extended Data Fig. 5b, Suppl Fig. 17, Table 15**). Notably, omission of the miRNA target abrogated any change in $R_{ct}$, whereas the complete COMPANION reaction (with and without crRNA present) yielded a clear $R_{ct}$ decrease after CRISPR reaction. These results mirror our fluorescence assays, confirming that Cas12a alone does not trigger reporter cleavage without the duplex–target interaction. The absence of off-target signal in no-duplex and no-target controls stresses the high specificity of the assay, a critical prerequisite for clinical diagnostics.[17]

We next evaluated the analytical performance of electrochemical COMPANION (or eCOMPANION) for four oncogenic or disease-associated miRNAs (miR-21, miR-340, miR-187, and miR-103). All four targets were detected with exceptional sensitivity in the attomolar range. Specifically, the LODs, defined using the 3:1 S/N (signal-to-noise) ratio method,[18] were approximately 28 aM for miR-340, 32 aM for miR-21, 48 aM for miR-187, and 45 aM for miR-103a-3p (**Extended Data Fig. 5a(i–iv)**; **Suppl Fig. 16(a–d)**). These values are on par with or exceed the sensitivity of state-of-the-art CRISPR-based miRNA sensors.[19] The percentage reduction in $R_{ct}$ increased progressively with higher concentrations of miRNA, exhibiting a linear dependence on the logarithm of miRNA concentration over ~6 orders of magnitude, with all calibration curves showing $R^2 \geq 0.90$ (**Suppl Fig. 16**). Moreover, the dynamic range and signal amplitudes were consistent across different miRNA sequences. As shown in **Extended Data Fig. 5a(i–iv)**, high (1 nM) and low (10 fM) target levels produced comparable relative signal changes for each of the four miRNAs tested, indicating uniform assay efficiency regardless of target identity.

To further demonstrate the versatility of our platform, we tested multiple Cas12a orthologs in the electrochemical format. Four Cas12a enzymes (AsCas12a, LbCas12a, MbCas12a, and Pb2Cas12a) were each deployed with the previously optimized DNA reporters (the C-rich reporter, found to be optimal in fluorescence assays, and an A-rich reporter for comparison). All orthologs were capable of detecting miR-340 over a wide concentration range (1 nM down to 100 aM, plus NTC), with robust electrochemical signal changes observable down to ~100 aM (**Extended Data Fig. 6d**). Consistent with our fluorescence results (**Extended Data Fig. 3**), the C-rich reporter outperformed the A-rich reporter for most Cas12a variants. In particular, LbCas12a, MbCas12a, and Pb2Cas12a showed substantially lower signals with the A-rich reporter, whereas AsCas12a exhibited excellent performance with both reporters (**Extended Data Fig. 5c,d**). Quantitatively, AsCas12a yielded a ~97-fold increase in current with the C-rich reporter versus ~76-fold with the A-rich reporter at 1 nM miR-340, indicating a modest signal advantage for the C-rich design (**Suppl Fig. 17**). Importantly, all ortholog/reporters combinations retained high specificity: reactions lacking target miRNA produced minimal background current (**Extended Data Fig. 5c,d**), in agreement with the control experiments described above. These findings reinforce that the choice of Cas12a ortholog and reporter sequence can be tuned to maximize sensitivity, and they further validate COMPANION's general applicability across different CRISPR enzymes. We note that both the C-rich and A-rich reporter assays with AsCas12a delivered robust signal gains (≥75-fold), underlining AsCas12a's superior *trans* cleavage efficiency in our system, an observation also reported in other studies leveraging engineered Cas12a for sensitive detection.[17]

Finally, we challenged the electrochemical COMPANION assay with a complex biological sample matrix to assess its potential for clinical deployment. Varying concentrations of miR-340 were spiked directly into diluted human saliva (5% saliva in buffer) and tested against an identical series in standard buffer (**Fig. 5d**). Remarkably, the saliva samples produced virtually indistinguishable signal responses compared to the pure buffer controls across all target levels (1 nM to 10 fM, as well as no-target negative controls). Even at the lowest tested concentration (10 fM), the signal obtained in saliva matched that in buffer, indicating no significant loss of sensitivity or increase in noise due to the saliva matrix. No sample pretreatment beyond simple dilution was required to achieve this performance. The ability to detect picomolar-to-attomolar miRNA levels in minimally processed saliva highlights the robustness and practicality of our platform for real-world specimens.

**COMPANION–RT-LAMP diagnostic workflow development.**

Building on the established system (COMPANION mode II), we next integrated the COMPANION platform with RT-LAMP for detecting respiratory viruses in patient samples. In a conventional CRISPR/Cas12 diagnostic, a designed gRNA is essential to target the sequence of interest.[20, 21] By contrast, COMPANION-RT-LAMP employs a miniCaR, which, together with the LAMP amplicon, assembles with Cas12a to activate *trans* cleavage of a reporter probe.[22] We initially verified that COMPANION could pair with LAMP amplification by designing short dumbbell-shaped DNA constructs mimicking the LAMP F-loop structures of three respiratory pathogens: Influenza A (Flu A H1N1), Influenza B (Flu B), and respiratory syncytial virus A (RSV A) – **Suppl Fig. 18**. As expected, all three synthetic loop-mimics robustly activated Cas12a *trans* cleavage, confirming COMPANION's compatibility with LAMP (**Suppl Fig. 19,20; Tables 13**). We then proceeded to test actual RT-LAMP products for these pathogens using a fluorescent readout.

Starting with AsCas12a, we began optimizing the COMPANION–RT-LAMP workflow using synthetic RNA targets for Flu A H1N1, RSV A, and Flu B by varying the volume of RT-LAMP reaction, followed by COMPANION-based detection of the amplified products (from 0 up to 30 µL added - **Suppl Fig. 19a**). Here, fluorescence was monitored in real time using a plate reader, with a ssDNA reporter R7 labeled at the 5′ end with fluorescein (FAM) and a 3′ Iowa Black FQ quencher. An input of 5 µL RT-LAMP product yielded the strongest signal-to-noise ratio for all three viruses (**Suppl Fig. 19b**);

therefore, this volume was selected for subsequent experiments. We next evaluated which LAMP-derived loop fragment most effectively triggers COMPANION. Testing the dumbbell's F-loop, B-loop, or both together showed that each loop alone could activate Cas12a, and combining both loops gave particularly robust activity (**Suppl Fig. 20**). We therefore adopted detection using both loop fragments. Consistent fluorescence kinetics and end-point readouts were obtained for Flu A H1N1, RSV A, and Flu B (**Suppl Fig. 20 (i–iii)**), demonstrating that COMPANION-RT-LAMP can sensitively report the presence of these viral targets. With the diagnostic workflow system optimized (**Extended Data Fig 6a**), we next evaluated the detection limits of the COMPANION-RT-LAMP assay for each pathogen. Remarkably, the system was able to detect down to ~2 copies/µL of Flu A H1N1 RNA, ~20 copies/µL of RSV A, and ~10 copies/µL of Flu B in the RT-LAMP reaction (**Extended Data Fig 6b**).

Having established the sensitivity of the COMPANION–RT-LAMP system down to clinically relevant concentrations, we next evaluated its specificity against a panel of globally relevant respiratory pathogens. Each of the three viral assays was challenged against a panel of six non-target respiratory viruses, including the human coronaviruses MERS-CoV, SARS-CoV-1, OC43, NL63, and 229E, and an avian influenza A (H7N9) virus. In all cases, only the intended target produced a detectable signal, while none of the non-target viral RNAs generated measurable fluorescence (**Suppl Fig. 21**; **Extended Data Fig 6c**). This 100% exclusivity mirrors the stringent specificity observed in other CRISPR-based diagnostics when carefully designed.[20] The absence of cross-reactivity indicates that the miniCaR in COMPANION effectively discriminates the correct LAMP amplicon, analogous to how a well-designed gRNA confers specificity in traditional Cas12a assays.[20] RNase P internal control results for patient nasopharyngeal samples tested for Influenza A (H1N1), Influenza B, or RSV A for RNA quality is presented (**Extended Data Fig. 6d**).

**COMPANION-RT-LAMP Validation in Patient Samples.**

Having confirmed excellent sensitivity and specificity with synthetic RNA targets (**Suppl Fig. 21**), we next evaluated COMPANION–RT-LAMP on clinical nasopharyngeal swab specimens as a step toward deployment in real-world clinical diagnostic programs (**Fig. 5e**). A total of 102 nasopharyngeal swab specimens were collected and processed at Mount Sinai Hospital (Toronto, Canada) and later used for the clinical trials of this study. Here, diagnostic results were benchmarked against standard RT-qPCR assays performed using WHO-established protocols.[23, 24] The COMPANION–RT-LAMP workflow involved a 30-minute RT-LAMP amplification followed by a 30-minute COMPANION detection step, providing a faster sample-to-result time than conventional RT-qPCR (1.5 hours).[25]

We first conducted a patient trial for influenza A (H1N1) using 39 de-identified clinical RNA samples. COMPANION–RT-LAMP correctly identified all nine RT-qPCR–positive samples and all 30 RT-qPCR–negative samples, yielding 100% accuracy (95% CI 90.97% to 100%) with the reference assay (**Fig. 5f(i–ii)**; **Extended Data Fig. 7a**). All positive samples, ranging from Ct values in the low 20s to about 32, were detected, with no false-positive signals observed. Based on the strong performance observed for influenza A,  we advanced to a second patient trial targeting influenza B (**Fig. 5g; Extended Data Fig. 7b**). Among 38 samples (23 RT-qPCR–positive and 15 negative), COMPANION–RT-LAMP correctly identified 22 of 23 true positives and all true negatives, corresponding to an overall accuracy of approximately 97.4% (95% CI 86.19% to 99.93%). A single discordant sample produced a weak signal (**Suppl Fig.  22**) and had a high Ct value (>32) by RT-qPCR, consistent with a borderline viral load near the detection limit.[26] Across three independent patient trials, COMPANION–RT-LAMP demonstrated robust clinical performance, achieving ≥97% accuracy and reliably detecting samples with RT-qPCR Ct values up to approximately 32. Building on these results, we next evaluated COMPANION–RT-LAMP in a third patient trial targeting RSV A. In this trial, all 25 samples (2 RT-qPCR–positive and 23 negative) were correctly classified, resulting in 100% accuracy (95% CI 86.28% to 100%) with RT-qPCR (**Fig. 5h; Extended Data Fig. 7c**). Both RSV-positive samples (Ct ≈ 28–30) were readily detected, with no false results. All performance evaluation statistics are detailed in **Tables 16-18.**

To further demonstrate the COMPANION system's flexibility, we evaluated its performance with multiple Cas12a orthologs beyond AsCas12a using a subset of Influenza B patient samples (n=38). Four enzymes:

AsCas12a, MbCas12a, LbCas12a, and Pb2Cas12a, were tested in parallel (**Fig. 5i, and Suppl Fig. 23**). All orthologs successfully detected true positive samples and yielded no signal for true negatives within 30 minutes (**Fig. 5i**). While AsCas12a showed the fastest kinetics, it also exhibited higher background fluorescence, whereas MbCas12a, LbCas12a, and Pb2Cas12a displayed flatter backgrounds and greater fold-change between positive and negative signals, albeit with slower kinetics (**Extended Data Fig. 7d**). Notably, the weak false-negative Flu B sample (Sample #38) was consistently undetected by all orthologs (Suppl **Fig. 24a–c**), indicating that the signal originated from borderline target amplification rather than ortholog-specific effects. Together, these results demonstrate that COMPANION–RT-LAMP is a reliable and adaptable diagnostic platform that works with multiple Cas12a variants without losing specificity, highlighting its potential for use in clinical diagnostic settings.

**Table S1.** Sequences of all miRNA-derived miniCaRs used throughout the study.

| S/N | Name | Sequence | miRBase |
|---|---|---|---|
| 1 | miR-21-5p | UAGCUUAUCAGACUGAUGUUGA | https://mirbase.org/mature/MIMAT0000076 |
| 2 | miR-320a-3p | AAAAGCUGGGUUGAGAGGGCGA | https://mirbase.org/mature/MIMAT0000510 |
| 3 | miR-340-5p | UUAUAAAGCAAUGAGACUGAUU | https://mirbase.org/mature/MIMAT0004650 |
| 4 | miR-210-3p | CUGUGCGUGUGACAGCGGCUGA | https://mirbase.org/mature/MIMAT0000267 |
| 5 | miR-187-3p | UCGUGUCUUGUGUUGCAGCCGG | https://mirbase.org/mature/MIMAT0000262 |
| 6 | miR-193b-5p | CGGGGUUUUGAGGGCGAGAUGA | https://mirbase.org/mature/MIMAT0004767 |
| 7 | miR-324-5p | CGCAUCCCCUAGGGCAUUGGUG | https://mirbase.org/mature/MIMAT0000761 |
| 8 | miR-483-3p | UCACUCCUCUCCUCCCGUCUU | https://mirbase.org/mature/MIMAT0002173 |
| 9 | miR-483-5p | AAGACGGGAGGAAAGAAGGGAG | https://mirbase.org/mature/MIMAT0004761 |
| 10 | miR-103-3p | AGCAGCAUUGUACAGGGCUAUGA | https://mirbase.org/mature/MIMAT0000101 |
| 11 | miR-29b-3p | UAGCACCAUUUGAAAUCAGUGUU | https://mirbase.org/mature/MIMAT0000100 |
| 12 | miR-19a-3p | UGUGCAAAUCUAUGCAAAACUGA | https://mirbase.org/mature/MIMAT0000073 |
| 13 | miR-17-5p | CAAAGUGCUUACAGUGCAGGUAG | https://mirbase.org/mature/MIMAT0000070 |
| 14 | miR-92a-1-5p | AGGUUGGGAUCGGUUGCAAUGCU | https://mirbase.org/mature/MIMAT0004507 |
| 15 | miR-301a-3p | CAGUGCAAUAGUAUUGUCAAAGC | https://mirbase.org/mature/MIMAT0000688 |
| 16 | miR-214-5p | UGCCUGUCUACACUUGCUGUGC | https://mirbase.org/mature/MIMAT0004564 |
| 17 | miR-155-5p | UUAAUGCUAAUCGUGAUAGGGGUU | https://mirbase.org/mature/MIMAT0000646 |
| 18 | miR-34-3p | CAAUCAGCAAGUAUACUGCCCU | https://mirbase.org/mature/MIMAT0004557 |
| 19 | miR-125b-5p | UCCCUGAGACCCUAACUUGUGA | https://mirbase.org/mature/MIMAT0000423 |
| 20 | Let-7a-3p | CUAUACAAUCUACUGUCUUUC | https://mirbase.org/mature/MIMAT0004481 |
| 21 | miR-206 | UGGAAUGUAAGGAAGUGUGUGG | https://mirbase.org/mature/MIMAT0000462 |
| 22 | miR-19-5p | AGUUUUGCAGGUUUGCAUUUCAGC | https://mirbase.org/mature/MIMAT0040166 |
| 23 | miR-491-5p | AGUGGGGAACCCUUCCAUGAGG | https://mirbase.org/mature/MIMAT0002807 |
| 24 | miR-34b-5p | UAGGCAGUGUCAUUAGCUGAUUG | https://mirbase.org/mature/MIMAT0000685 |
| 25 | miR-34c-5p | AGGCAGUGUAGUUAGCUGAUUGC | https://mirbase.org/mature/MIMAT0000686 |
| 26 | miR-200a-5p | CAUCUUACCGGACAGUGCUGGA | https://mirbase.org/mature/MIMAT0001620 |
| 27 | miR-200c-3p | UAAUACUGCCGGGUAAUGAUGGA | https://mirbase.org/mature/MIMAT0000617 |
| 28 | miR-15a | UAGCAGCACAUAAUGGUUUGUG | https://mirbase.org/mature/MIMAT0000068 |
| 29 | miR-16-5p | UAGCAGCACGUAAAUAUUGGCG | https://mirbase.org/mature/MIMAT0000069 |
| 30 | miR-181a-5p | AACAUUCAACGCUGUCGGUGAGU | https://mirbase.org/mature/MIMAT0000256 |
| 31 | miR-194-5p | UGUAACAGCAACUCCAUGUGGA | https://mirbase.org/mature/MIMAT0000460 |
| 32 | miR-107 | AGCAGCAUUGUACAGGGCUAUCA | https://mirbase.org/mature/MIMAT0000104 |
| 33 | miR-9-3p | AUAAAGCUAGAUAACCGAAAGU | https://mirbase.org/mature/MIMAT0000442 |
| 34 | miR-489-5p | GGUCGUAUGUGUGACGCCAUUU | https://mirbase.org/mature/MIMAT0026605 |
| 35 | miR-195-5p | UAGCAGCACAGAAAUAUUGGC | https://mirbase.org/mature/MIMAT0000461 |
| 36 | miR-25-5p | AGGCGGAGACUUGGGCAAUUG | https://mirbase.org/mature/MIMAT0004498 |
| 37 | miR-106b-3p | CCGCACUGUGGGUACUUGCUGC | https://mirbase.org/mature/MIMAT0004672 |
| 38 | miR-1915-3p | CCCCAGGGCGACGCGGCGGG | https://mirbase.org/mature/MIMAT0007892 |
| 39 | miR-3665 | AGCAGGUGCGGGGCGGCG | https://mirbase.org/mature/MIMAT0018087 |
| 40 | miR-4787-5p | GCGGGGGUGGCGGCGGCAUCCC | https://mirbase.org/mature/MIMAT0019956 |
| 41 | miR-4497 | CUCCGGGACGGCUGGGC | https://mirbase.org/mature/MIMAT0019032 |
| 42 | miR-3656 | GGCGGGUGCGGGGGUGG | https://mirbase.org/mature/MIMAT0018076 |
| 43 | miR-3960 | GGCGGCGGCGGAGGCGGGGG | https://mirbase.org/mature/MIMAT0019337 |
| 44 | miR-4488 | AGGGGGCGGGCUCCGGCG | https://mirbase.org/mature/MIMAT0019022 |

**Table S2.** DNA-equivalent sequences corresponding to all miRNAs used in this study.

| S/N | Name | Sequence (DNA Equivalence) |
|---|---|---|
| 1 | miR-21-5p | TAGCTTATCAGACTGATGTTGA |
| 2 | miR-320a-3p | AAAAGCTGGGTTGAGAGGGCGA |
| 3 | miR-340-5p | TTATAAAGCAATGAGACTGATT |
| 4 | miR-210-3p | CTGTGCGTGTGACAGCGGCTGA |
| 5 | miR-187-3p | TCGTGTCTTGTGTTGCAGCCGG |
| 6 | miR-193b-5p | CGGGGTTTTGAGGGCGAGATGA |
| 7 | miR-324-5p | CGCATCCCCTAGGGCATTGGTG |
| 8 | miR-483-3p | TCACTCCTCTCCTCCCGTCTT |
| 9 | miR-483-5p | AAGACGGGAGGAAAGAAGGGAG |
| 10 | miR-103-3p | AGCAGCATTGTACAGGGCTATGA |
| 11 | miR-29b-3p | TAGCACCATTTGAAATCAGTGTT |
| 12 | miR-19a-3p | TGTGCAAATCTATGCAAAACTGA |
| 13 | miR-17-5p | CAAAGTGCTTACAGTGCAGGTAG |
| 14 | miR-92a-1-5p | AGGTTGGGATCGGTTGCAATGCT |
| 15 | miR-301a-3p | CAGTGCAATAGTATTGTCAAAGC |
| 16 | miR-214-5p | TGCCTGTCTACACTTGCTGTGC |
| 17 | miR-155-5p | TTAATGCTAATCGTGATAGGGGTT |
| 18 | miR-34-3p | CAATCAGCAAGTATACTGCCCT |
| 19 | miR-125b-5p | TCCCTGAGACCCTAACTTGTGA |
| 20 | Let-7a-3p | CTATACAATCTACTGTCTTTC |
| 21 | miR-206 | TGGAATGTAAGGAAGTGTGTGG |
| 22 | miR-19-5p | AGTTTTGCAGGTTTGCATTTCAGC |
| 23 | miR-491-5p | AGTGGGGAACCCTTCCATGAGG |
| 24 | miR-34b-5p | TAGGCAGTGTCATTAGCTGATTG |
| 25 | miR-34c-5p | AGGCAGTGTAGTTAGCTGATTGC |
| 26 | miR-200a-5p | CATCTTACCGGACAGTGCTGGA |
| 27 | miR-200c-3p | TAATACTGCCGGGTAATGATGGA |
| 28 | miR-15a | TAGCAGCACATAATGGTTTGTG |
| 29 | miR-16-5p | TAGCAGCACGTAAATATTGGCG |
| 30 | miR-181a-5p | AACATTCAACGCTGTCGGTGAGT |
| 31 | miR-194-5p | TGTAACAGCAACTCCATGTGGA |
| 32 | miR-107 | AGCAGCATTGTACAGGGCTATCA |
| 33 | miR-9-3p | ATAAAGCTAGATAACCGAAAGT |
| 34 | miR-489-5p | GGTCGTATGTGTGACGCCATTT |
| 35 | miR-195-5p | TAGCAGCACAGAAATATTGGC |
| 36 | miR-25-5p | AGGCGGAGACTTGGGCAATTG |
| 37 | miR-106b-3p | CCGCACTGTGGGTACTTGCTGC |
| 38 | miR-1915-3p | CCCCAGGGCGACGCGGCGGG |
| 39 | miR-3665 | AGCAGGTGCGGGGCGGCG |
| 40 | miR-4787-5p | GCGGGGGTGGCGGCGGCATCCC |
| 41 | miR-4497 | CTCCGGGACGGCTGGGC |
| 42 | miR-3656 | GGCGGGTGCGGGGGTGG |
| 43 | miR-3960 | GGCGGCGGCGGAGGCGGGGG |
| 44 | miR-4488 | AGGGGGCGGGCTCCGGCG |

**Table S3.** Sequences of DNA duplex probes designed for all 44 miRNAs used in this study.

| S/N | Name | Sequence Types | Duplexed Probes _ Purchased from IDT |
|---|---|---|---|
| 1 | Duplex_miR-21-5p | Sense | GAAGTTCATGTTTCTGAATCAACATCAGTCTGATAAGCTATTCAAG |
| | | Anti-sense | /5Phos/TTCAGAAACATGAACTTC |
| 2 | Duplex_miR-320a-3p | Sense | GAAGTTCAT GTTTCTGAATCGCCCTCTCAACCCAGCTTTTTCAAG |
| | | Anti-sense | TTCAGAAACATGAACTTC |
| 3 | Duplex_miR-340-5p | Sense | GAAGTTCAT GTTTCTGAA AATCAGTCTCATTGCTTTATAAAATATG |
| | | Anti-sense | TTCAGAAACATGAACTTC |
| 4 | Duplex_miR-210-3p | Sense | GAAGTTCATGTTTCTGAATCAGCCGCTGTCACACGCACAGTTCAAG |
| | | Anti-sense | TTCAGAAACATGAACTTC |
| 5 | Duplex_miR-187-3p | Sense | GAAGTTCATGTTTCTGAACCGGCTGCAACACAAGACACGAAATATG |
| | | Anti-sense | TTCAGAAACATGAACTTC |
| 6 | Duplex_miR-193b-5p | Sense | GAAGTTCATGTTTCTGAATCATCTCGCCCTCAAA ACCCCGAATATG |
| | | Anti-sense | TTCAGAAACATGAACTTC |
| 7 | Duplex_miR-324-5p | Sense | GAAGTTCATGTTTCTGAACACCAATGCCCTAGGG GATGCGAATATG |
| | | Anti-sense | TTCAGAAACATGAACTTC |
| 8 | Duplex_miR-483-3p | Sense | GAAGTTCAT GTTTCTGAAAAGACGGGAGGAGAGGAGTGAAATATG |
| | | Anti-sense | /5Phos/TT CAG AAA CAT GAA CTT C |
| 9 | Duplex_miR-483-5p | Sense | GAA GTTCAT GTT TCTGAA CTC CCTTCTTTCCTCCCGTCTTAATATG |
| | | Anti-sense | /5Phos/TT CAG AAA CAT GAA CTT C |
| 10 | Duplex_miR-103-3p | Sense | GAA GTT CATGTTTCTGAATCATAGCCCTGTACAATGCTGCTAATATG |
| | | Anti-sense | /5Phos/TT CAG AAA CAT GAA CTT C |
| 11 | Duplex_miR-29b-3p | Sense | GAAGTTCATGTTTCTGAAAACACTGATTTCAAATGGTGCTATTCAAG |
| | | Anti-sense | TTCAGAAACATGAACTTC |
| 12 | Duplex_miR-19a-3p | Sense | GAAGTTCAT GTTTCTGAATCAGTTTTGCATAGATTTGCACATTCAAG |
| | | Anti-sense | TTCAGAAACATGAACTTC |
| 13 | Duplex_miR-17-5p | Sense | GAAGTTCATGTTTCTGAACTACCTGCACTGTAAGCACTTTGTTCAAG |
| | | Anti-sense | TTCAGAAACATGAACTTC |
| 14 | Duplex_miR-92a-1-5p | Sense | GAAGTTCATGTTTCTGAAAGCATTGCAACCGATCCCAACCTTTCAAG |
| | | Anti-sense | TTCAGAAACATGAACTTC |
| 15 | Duplex_miR-301a-3p | Sense | GAAGTTCAT GTTTCTGAAGCTTTGACAATACTATTGCACTGTTCAAG |
| | | Anti-sense | TTCAGAAACATGAACTTC |
| 16 | Duplex_miR-214-5p | Sense | GAAGTTCAT GTTTCTGAAGCACAGCAAGTGTAGACAGGCATTCAAG |
| | | Anti-sense | TTCAGAAACATGAACTTC |
| 17 | Duplex_miR-155-5p | Sense | GAAGTTCATGTTTCTGAAAACCCCTATCACGATTAGCATTAATTCAAG |
| | | Anti-sense | TTCAGAAACATGAACTTC |
| 18 | Duplex_miR-34-3p | Sense | GAAGTTCAT GTTTCTGAA AGGGCAGTATACTTGCTGATTGTTCAAG |
| | | Anti-sense | TTCAGAAACATGAACTTC |
| 19 | Duplex_miR-125b-5p | Sense | GAAGTTCAT GTTTCTGAA TCACAAGTTAGGGTCTCAGGGATTCAAG |
| | | Anti-sense | TTCAGAAACATGAACTTC |
| 20 | Duplex_Let-7a-3p | Sense | GAAGTTCAT GTTTCTGAA GAAAGACAGTAGATTGTATAG TTCAAG |
| | | Anti-sense | TTCAGAAACATGAACTTC |
| 21 | Duplex_miR-206 | Sense | GAAGTTCAT GTTTCTGAA CCACACACTTCCTTACATTCCA TTCAAG |
| | | Anti-sense | TTCAGAAACATGAACTTC |
| 22 | Duplex_miR-19-5p | Sense | GAAGTTCATGTTTCTGAAGCTGAAATGCAAACCTGCAAAACTTTCAAG |
| | | Anti-sense | TTCAGAAACATGAACTTC |

| | | | |
|---|---|---|---|
| 23 | Duplex_miR-491-5p | Sense | GAAGTTCAT GTTTCTGAA CCTCATGGAAGGGTTCCCCACT TTCAAG |
| | | Anti-sense | TTCAGAAACATGAACTTC |
| 24 | Duplex_miR-34b-5p | Sense | GAAGTTCAT GTTTCTGAA CAATCAGCTAATGACACTGCCTA TTCAAG |
| | | Anti-sense | TTCAGAAACATGAACTTC |
| 25 | Duplex_miR-34c-5p | Sense | GAAGTTCAT GTTTCTGAA GCAATCAGCTAACTACACTGCCTTTCAAG |
| | | Anti-sense | TTCAGAAACATGAACTTC |
| 26 | Duplex_miR-200a-5p | Sense | GAAGTTCAT GTTTCTGAA TCCAGCACTGTCCGGTAAGATG TTCAAG |
| | | Anti-sense | TTCAGAAACATGAACTTC |
| 27 | Duplex_miR-200c-3p | Sense | GAAGTTCAT GTTTCTGAA TCCATCATTACCCGGCAGTATTA TTCAAG |
| | | Anti-sense | TTCAGAAACATGAACTTC |
| 28 | Duplex_miR-15a | Sense | GAAGTTCAT GTTTCTGAA CACAAACCATTATGTGCTGCTA TTCAAG |
| | | Anti-sense | TTCAGAAACATGAACTTC |
| 29 | Duplex_miR-16-5p | Sense | GAAGTTCAT GTTTCTGAA CGCCAATATTTACGTGCTGCTA TTCAAG |
| | | Anti-sense | TTCAGAAACATGAACTTC |
| 30 | Duplex_miR-181a-5p | Sense | GAAGTTCAT GTTTCTGAA ACTCACCGACAGCGTTGAATGTTTTCAAG |
| | | Anti-sense | TTCAGAAACATGAACTTC |
| 31 | Duplex_miR-194-5p | Sense | GAAGTTCAT GTTTCTGAA TCCACATGGAGTTGCTGTTACA TTCAAG |
| | | Anti-sense | TTCAGAAACATGAACTTC |
| 32 | Duplex_miR-107 | Sense | GAAGTTCAT GTTTCTGAA TGATAGCCCTGTACAATGCTGCTTTCAAG |
| | | Anti-sense | TTCAGAAACATGAACTTC |
| 33 | Duplex_miR-9-3p | Sense | GAAGTTCAT GTTTCTGAA ACTTTCGGTTATCTAGCTTTAT TTCAAG |
| | | Anti-sense | TTCAGAAACATGAACTTC |
| 34 | Duplex_miR-489-5p | Sense | GAAGTTCAT GTTTCTGAA AAATGGCGTCACACATACGACC TTCAAG |
| | | Anti-sense | TTCAGAAACATGAACTTC |
| 35 | Duplex_miR-195-5p | Sense | GAAGTTCAT GTTTCTGAA GCCAATATTTCTGTGCTGCTA TTCAAG |
| | | Anti-sense | TTCAGAAACATGAACTTC |
| 36 | Duplex_miR-25-5p | Sense | GAAGTTCAT GTTTCTGAA CAATTGCCCAAGTCTCCGCCT TTCAAG |
| | | Anti-sense | TTCAGAAACATGAACTTC |
| 37 | Duplex_miR-106b-3p | Sense | GAAGTTCAT GTTTCTGAA GCAGCAAGTACCCACAGTGCGGTTCAAG |
| | | Anti-sense | TTCAGAAACATGAACTTC |
| 38 | Duplex_miR-1915-3p | Sense | GAAGTTCAT GTTTCTGAA CCCGCCGCGTCGCCCTGGGG TTCAAG |
| | | Anti-sense | TTCAGAAACATGAACTTC |
| 39 | Duplex_miR-3665 | Sense | GAAGTTCAT GTTTCTGAA CGCCGCCCCGCACCTGCT TTCAAG |
| | | Anti-sense | TTCAGAAACATGAACTTC |
| 40 | Duplex_miR-4787-5p | Sense | GAAGTTCAT GTTTCTGAA GGGATGCCGCCGCCACCCCCGC TTCAAG |
| | | Anti-sense | TTCAGAAACATGAACTTC |
| 41 | Duplex_miR-4497 | Sense | GAAGTTCAT GTTTCTGAA GCCCAGCCGTCCCGGAG TTCAAG |
| | | Anti-sense | TTCAGAAACATGAACTTC |
| 42 | Duplex_miR-3656 | Sense | GAAGTTCAT GTTTCTGAA CCACCCCCGCACCCGCC TTCAAG |
| | | Anti-sense | TTCAGAAACATGAACTTC |
| 43 | Duplex_miR-3960 | Sense | GAAGTTCAT GTTTCTGAA CCCCCGCCTCCGCCGCCGCC TTCAAG |
| | | Anti-sense | TTCAGAAACATGAACTTC |
| 44 | Duplex_miR-4488 | Sense | GAAGTTCAT GTTTCTGAA CGCCGGAGCCCGCCCCCT TTCAAG |
| | | Anti-sense | TTCAGAAACATGAACTTC |

**Table S4.** Sequences of crRNAs designed for all 44 miRNAs used in this study.

| S/N | Name | crRNAs (Purchased from IDT) |
|---|---|---|
| 1 | crRNA_miR-21-5p | /AltR1/rUrArArUrUrUrCrUrArCrUrCrUrUrGrUrArGrArUrUrGrArArUrCrArAr CrArUrCrArGrUrCrUrGrArU/AltR2/ |
| 2 | crRNA_miR-320a-3p | /AltR1/rUrA rArUrU rUrCrU rArCrU rCrUrU rGrUrA rGrArU rCrGrC rCrCrU rCrUrC rArArC rCrCrA rGrCrU rUrU/AltR2/ |
| 3 | crRNA_miR-340-5p | /AltR1/rUrA rArUrU rUrCrU rArCrU rCrUrU rGrUrA rGrArU rUrGrA rArArA rUrCrA rGrUrC rUrCrA rUrUrG rCrU/AltR2/ |
| 4 | crRNA_miR-210-3p | /AltR1/rUrArArUrUrUrCrUrArCrUrCrUrUrGrUrArGrArUr UrGrArAr UrCrArGrCrCrGrCrUrGrUrCrArCrArC/AltR2/ |
| 5 | crRNA_miR-187-3p | /AltR1/rUrA rArUrU rUrCrU rArCrU rCrUrU rGrUrA rGrArU rUrGrA rArCrC rGrGrC rUrGrC rArArC rArCrA rArG/AltR2/ |
| 6 | crRNA_miR-193b-5p | /AltR1/rUrA rArUrU rUrCrU rArCrU rCrUrU rGrUrA rGrArU rUrGrA rArUrC rArUrC rUrCrG rCrCrC rUrCrA rArA/AltR2/ |
| 7 | crRNA_miR-324-5p | /AltR1/rUrA rArUrU rUrCrU rArCrU rCrUrU rGrUrA rGrArU rUrGrA rArCrA rCrCrA rArUrG rCrCrC rUrArG rGrG/AltR2/ |
| 8 | crRNA_miR-483-3p | /AltR1/rUrA rArUrU rUrCrU rArCrU rCrUrU rGrUrA rGrArU rUrGrA rArArA rGrArC rGrGrG rArGrG rArGrA rGrG/AltR2/ |
| 9 | crRNA_miR-483-5p | /AltR1/rUrA rArUrU rUrCrU rArCrU rCrUrU rGrUrA rGrArU rUrGrA rArCrU rCrCrC rUrUrC rUrUrU rCrCrU rCrC/AltR2/ |
| 10 | crRNA_miR-103-3p | /AltR1/rUrA rArUrU rUrCrU rArCrU rCrUrU rGrUrA rGrArU rUrGrA rArUrC rArUrA rGrCrC rCrUrG rUrArC rArA/AltR2/ |
| 11 | crRNA_miR-29b-3p | UAAUUUCUACUCUUGUAGAU UGAA AACACUGAUUUCAAAU |
| 12 | crRNA_miR-19a-3p | UAAUUUCUACUCUUGUAGAU UGAA UCAGUUUUGCAUAGAU |
| 13 | crRNA_miR-17-5p | UAAUUUCUACUCUUGUAGAU UGAA CUACCUGCACUGUAAG |
| 14 | crRNA_miR-92a-1-5p | UAAUUUCUACUCUUGUAGAU UGAA AGCAUUGCAACCGAUC |
| 15 | crRNA_miR-301a-3p | UAAUUUCUACUCUUGUAGAU UGAA GCUUUGACAAUACUAU |
| 16 | crRNA_miR-214-5p | UAAUUUCUACUCUUGUAGAU UGAA GCACAGCAAGUGUAGA |
| 17 | crRNA_miR-155-5p | UAAUUUCUACUCUUGUAGAU UGAA AACCCCUAUCACGAUU |
| 18 | crRNA_miR-34-3p | UAAUUUCUACUCUUGUAGAU UGAA AGGGCAGUAUACUUGC |
| 19 | crRNA_miR-125b-5p | UAAUUUCUACUCUUGUAGAU UGAA UCACAAGUUAGGGUCU |
| 20 | crRNA_Let-7a-3p | UAAUUUCUACUCUUGUAGAU UGAA GAAAGACAGUAGAUUG |
| 21 | crRNA_miR-206 | UAAUUUCUACUCUUGUAGAU UGAA CCACACACUUCCUUAC |
| 22 | crRNA_miR-19-5p | UAAUUUCUACUCUUGUAGAU UGAA GCUGAAAUGCAAACCU |
| 23 | crRNA_miR-491-5p | UAAUUUCUACUCUUGUAGAU UGAA CCUCAUGGAAGGGUUC |
| 24 | crRNA_miR-34b-5p | UAAUUUCUACUCUUGUAGAU UGAA CAAUCAGCUAAUGACA |
| 25 | crRNA_miR-34c-5p | UAAUUUCUACUCUUGUAGAU UGAA GCAAUCAGCUAACUAC |
| 26 | crRNA_miR-200a-5p | UAAUUUCUACUCUUGUAGAU UGAA UCCAGCACUGUCCGGU |
| 27 | crRNA_miR-200c-3p | UAAUUUCUACUCUUGUAGAU UGAA UCCAUCAUUACCCGGC |
| 28 | crRNA_miR-15a | UAAUUUCUACUCUUGUAGAU UGAA CACAAACCAUUAUGUG |
| 29 | crRNA_miR-16-5p | UAAUUUCUACUCUUGUAGAU UGAA CGCCAAUAUUUACGUG |
| 30 | crRNA_miR-181a-5p | UAAUUUCUACUCUUGUAGAU UGAA ACUCACCGACAGCGUU |
| 31 | crRNA_miR-194-5p | UAAUUUCUACUCUUGUAGAU UGAA UCCACAUGGAGUUGCU |
| 32 | crRNA_miR-107 | UAAUUUCUACUCUUGUAGAU UGAA UGAUAGCCCUGUACAA |
| 33 | crRNA_miR-9-3p | UAAUUUCUACUCUUGUAGAU UGAA ACUUUCGGUUAUCUAG |

| | | |
|---|---|---|
| 34 | crRNA_miR-489-5p | UAAUUUCUACUCUUGUAGAU UGAA AAAUGGCGUCACACAU |
| 35 | crRNA_miR-195-5p | UAAUUUCUACUCUUGUAGAU UGAA GCCAAUAUUUCUGUGC |
| 36 | crRNA_miR-25-5p | UAAUUUCUACUCUUGUAGAU UGAA CAAUUGCCCAAGUCUC |
| 37 | crRNA_miR-106b-3p | UAAUUUCUACUCUUGUAGAU UGAA GCAGCAAGUACCCACA |
| 38 | crRNA_miR-1915-3p | UAAUUUCUACUCUUGUAGAU UGAA CCCGCCGCGUCGCCCU |
| 39 | crRNA_miR-3665 | UAAUUUCUACUCUUGUAGAU UGAA CGCCGCCCCGCACCUG |
| 40 | crRNA_miR-4787-5p | UAAUUUCUACUCUUGUAGAU UGAA GGGAUGCCGCCGCCAC |
| 41 | crRNA_miR-4497 | UAAUUUCUACUCUUGUAGAU UGAA GCCCAGCCGUCCCGGA |
| 42 | crRNA_miR-3656 | UAAUUUCUACUCUUGUAGAU UGAA CCACCCCCGCACCCGC |
| 43 | crRNA_miR-3960 | UAAUUUCUACUCUUGUAGAU UGAA CCCCCGCCUCCGCCGC |
| 44 | crRNA_miR-4488 | UAAUUUCUACUCUUGUAGAU UGAA CGCCGGAGCCCGCCCC |

*crRNAs with "r" are directly purchased from IDT, while the ones without "r" are through in vitro transcription

**Table S5**. Sequences of miR-21 DNA duplex probes containing different PAM variants.

| S/N | Type of PAM | Name | Sequence Types | Sequences |
|---|---|---|---|---|
| 1 | Canonical PAM | Duplex_TTTC_miR-21 | Sense | GAAGTTCAT GTTTCTGAA TCAACATCAGTCTGAT AAGCTA TTCAAG |
| | | | Anti-sense | TTCAGAAACATGAACTTC |
| 2 | Suboptimal PAMs (VTTV, TCTV, TTVV, YYYN, TVTV, TTVV) | Duplex_GTTG_miR-21 | Sense | GAAGTTCAT GGTTGTGAA TCAACATCAGTCTGAT AAGCTA TTCAAG |
| | | | Anti-sense | TTCACAACCATGAACTTC |
| 3 | | Duplex_CTTA_miR-21 | Sense | GAAGTTCAT GCTTATGAA TCAACATCAGTCTGAT AAGCTA TTCAAG |
| | | | Anti-sense | TTCATAAGCATGAACTTC |
| 4 | | Duplex_TTGT_miR-21 | Sense | GAAGTTCAT GTTGTTGAA TCAACATCAGTCTGAT AAGCTA TTCAAG |
| | | | Anti-sense | TTCAACAACATGAACTTC |
| 5 | | Duplex_TTGG_miR-21 | Sense | GAAGTTCAT GTTGGTGAA TCAACATCAGTCTGAT AAGCTA TTCAAG |
| | | | Anti-sense | TTCACCAACATGAACTTC |
| 6 | | Duplex_TCTC_miR-21 | Sense | GAAGTTCAT GTCTCTGAA TCAACATCAGTCTGAT AAGCTA TTCAAG |
| | | | Anti-sense | TTCAGAGACATGAACTTC |
| 7 | Anti - PAMs (TTTT, AAAA, CCCC, and GGGG) | Duplex_TTTT_miR-21 | Sense | GAAGTTCAT GTTTTTGAA TCAACATCAGTCTGAT AAGCTA TTCAAG |
| | | | Anti-sense | TTCAAAAACATGAACTTC |
| 8 | | Duplex_AAAA_miR-21 | Sense | GAAGTTCAT GAAAATGAA TCAACATCAGTCTGAT AAGCTA TTCAAG |
| | | | Anti-sense | TTCATTTTCATGAACTTC |
| 9 | | Duplex_CCCC_miR-21 | Sense | GAAGTTCAT GCCCCTGAA TCAACATCAGTCTGAT AAGCTA TTCAAG |
| | | | Anti-sense | TTCAGGGGCATGAACTTC |
| 10 | | Duplex_GGGG_miR-21 | Sense | GAAGTTCAT GGGGGTGAA TCAACATCAGTCTGAT AAGCTA TTCAAG |
| | | | Anti-sense | TTCACCCCCATGAACTTC |
| *Letters in red depicts the PAM sites | | | | |

**Table S6.** Sequences of chimeric duplex probes for miR-21 with and without 5′ phosphorylation.

| S/N | Name | Probe type | Sequence - purchased from IDT |
|---|---|---|---|
| 1 | DNA(miR-21) | | TAG CTT ATC AGA CTG ATG TTG A |
| 2 | miR21_Chimeric duplex | Sense | GAA GTT CAT GTT TCT GAA rUrCrA rArCrA rUrCrA rGrUrC rUrGrA rUrArA rGrCrU rA TT CAA G |
| | | Anti-sense | TTC AGA AAC ATG AAC TTC |
| 3 | miR21_5'Phos chimeric duplex | Sense | GAA GTT CAT GTT TCT GAA rUrCrA rArCrA rUrCrA rGrUrC rUrGrA rUrArA rGrCrU rA TT CAA G |
| | | Anti-sense | /5Phos/TT CAG AAA CAT GAA CTT C |
| *Red portion is RNA and black is DNA | | | |

**Table S7.** Probe sequences used in fluorescence polarization assay (FPA) experiments.

| S/N | Name | Probe type | Sequence - purchased from IDT |
|---|---|---|---|
| 1 | miR-21-5p | | UAGCUUAUCAGACUGAUGUUGA |
| 2 | miR-21-5p-FAM | | /56-FAM/rUrArGrCrUrUrArUrCrArGrArCrUrGrArUrGrUrUrGrA |
| 3 | miR-210-3p-FAM | | /56-FAM/rCrU rGrUrG rCrGrU rGrUrG rArCrA rGrCrG rGrCrU rGrA |
| 4 | miR-324-5p - FAM | | /56-FAM/rCrG rCrArU rCrCrC rCrUrA rGrGrG rCrArU rUrGrG rUrG |
| | gRNA_miR-21-5p | | /AltR1/rUrArArUrUrUrCrUrArCrUrCrUrUrGrUrArGrArUrUrGrArArUrCrArArCrAr UrCrArGrUrCrUrGrArU/AltR2/ |
| 5 | Duplex_miR-21 | Sense | GAAGTTCAT GTTTCTGAA TCAACATCAGTCTGAT AAGCTA TTCAAG/36-FAM/ |
| | | Anti-sense | TTCAGAAACATGAACTTC |

**Table S8.** Probe sequences used for evaluation of the crRNA-independent system for ssDNA and dsDNA detection.

| Type | Names | Sequence |
|---|---|---|
| Influenza A (Flu A) | Flu A gBlock | GAGCTAAGAGAGCAATTGAGCTCAGTGTCATCATTTG AAAGGTTTGAGATATTCCCCAAGACAAGTTCATGGCC CAATCATGACTCGAACAAAGGTGTAACGGCAGCATGT CCTCATGCTGGAGCAAAAAGCTTCTACAAAAATTTAA TATGGCTAGTTAAAAAAGGAAATTCATACCCAAAGCT CAGCAAATCCTACATTA |
| | Flu A ssDNA | TGACTCGAACAAAGGTGTAACGGCAGCATGTCCTCAT GCTGGAGCAAAAAGCTTCTACAAAAATTTAATATGGC TAGTTAAAAAAGGAAA |
| | Flu A miniCaR 5 (GC:41%) | UAGAAGCUUUUUGCUCCAGCAU |
| | Flu A crRNA | UAAUUUCUACUCUUGUAGAU UAGAAGCUUUUUGCUCCAGCAU |
| | Flu A miniCaR 6 (GC:18%) | UUUAACUAGCCAUAUUAAAUUU |
| | Flu A miniCaR 4 (GC:64%) | CCAGCAUGAGGACAUGCUGCCG |
| | Flu A miniCaR 3 (GC:50%) | GUUCGAGUCAUGAUUGGGCCAU |

| | | |
|---|---|---|
| | Flu A miniCaR 2 (GC:59%) | GGGCCAUGAACUUGUCUUGGGG |
| | Flu A miniCaR 1 (GC:32%) | AUCUCAAACCUUUCAAAUGAUG |
| Influenza B (Flu B) | Flu B gBlock | CGGAGCTGCCTATGAAGACCTGAGAGTTTTGTCTGCATTAACAGGCACAGAATTCAAGCCTAGATCAGCATTAAAATGCAAGGGTTTCCATGTTCCAGCAAAGGAACAGGTAGAAGGGATGGGAGCAGCTCTGATGTCCATCAAGCTCCAGTTTTGGGCTCCGATGACCAGATCTGGGGGGAACGAAGTAGGTGGAGACGGA |
| | Flu B ssDNA | TGAGAGTTTTGTCTGCATTAACAGGCACAGAATTCAAGCCTAGATCAGCATTAAAATGCAAGGGTTTCCATGTTCCAGCAAAGGAACAGG |
| | Flu B miniCaR 3 (GC:41%) | AUGCUGAUCUAGGCUUGAAUUC |
| | Flu B crRNA | UAAUUUCUACUCUUGUAGAU AUGCUGAUCUAGGCUUGAAUUC |
| | Flu B miniCaR 1 (GC:59) | CAGGUCUUCAUAGGCAGCUCCG |
| | Flu B miniCaR 2 (GC:45%) | UUCUGUGCCUGUUAAUGCAGAC |
| | Flu B miniCaR 4 (GC:50%) | CUUUGCUGGAACAUGGAAACCC |
| | Flu B miniCaR 5 (GC:55%) | AUGGACAUCAGAGCUGCUCCCA |
| | Flu B miniCaR 6 (GC:64%) | CCAGAUCUGGUCAUCGGAGCCC |
| RSV A | RSV A gBlock | CTGCTGTTCAATACAATGTCCTAGAAAAAGACGATGATCCTGCATCACTTACAATATGGGTACCCATGTTCCAATCATCCTTGCCAGCAGATCTACTCATAAAAGAACTAGCCAATGTTAATATACTAGTGAAACAAATATCCACACCCAAGGGACCCTCATTAAGAGTCATGATAAACTCAAGAAGTGCAGTGCTAGCACAAATGCCCAGCAAATTTACCAT |
| | RSV A ssDNA | CCTTGCCAGCAGATCTACTCATAAAAGAACTAGCCAATGTTAATATACTAGTGAAACAAATATCCACACCCAAGGGACCCTCATTAAGAG |
| | RSV A miniCaR 4 (GC:27%) | ACUAGUAUAUUAACAUUGGCUA |
| | RSV A crRNA | UAAUUUCUACUCUUGUAGAU ACUAGUAUAUUAACAUUGGCUA |
| | RSV A miniCaR 1 (GC:41%) | CUAGGACAUUGUAUUGAACAGC |
| | RSV A miniCaR 2(GC:45%) | UAAGUGAUGCAGGAUCAUCGUC |
| | RSV A miniCaR 3 (GC:50%) | GAUGAUUGGAACAUGGGUACCC |
| | RSV A miniCaR 5 (GC:59%) | AAUGAGGGUCCCUUGGGUGUGG |
| | RSV A miniCaR 6 (GC:55%) | UGGGCAUUUGUGCUAGCACUGC |
| *All RNA probes were made through in vitro transcription | | |

**Table S9**. Sequences of FAM–quencher-labeled probes used for fluorescence readout following trans-cleavage.

| S/N | Name | Short form | Sequences (5′-3′) | Purification |
|---|---|---|---|---|
| 1 | Reporter 1 | R1 | /56-FAM/TTATTT/31ABkFQ | HPLC |
| 2 | Reporter 2 | R2 | /56-FAM/rUrUrArUrU/31ABkFQ | HPLC |
| 3 | Reporter 3 | R3 | /56-FAM/rUArUArUA/31ABkFQ | HPLC |
| 4 | Reporter 4 | R4 | /56-FAM/ArUArUArU/31ABkFQ | HPLC |
| 5 | Reporter5 | R5 | /56-FAM/TTTTTT/31ABkFQ | HPLC |
| 6 | Reporter 6 | R6 | /56-FAM/AAAAAA/31ABkFQ | HPLC |
| 7 | Reporter 7 | R7 | /56-FAM/CCCCCC/31ABkFQ | HPLC |

| | | | | |
|---|---|---|---|---|
| 8 | Reporter 8 | R8 | /56-FAM/GGGGGG/31ABkFQ | HPLC |
| 9 | Reporter 9 | R9 | /56-FAM/AAAAAA/31ABkFQ | HPLC |
| 10 | Reporter 10 | R10 | /56-FAM/UUUUUU/31ABkFQ | HPLC |
| 11 | Reporter 11 | R11 | /56-FAM/CCCCCC/31ABkFQ | HPLC |
| 12 | Reporter 12 | R12 | /56-FAM/GGGGGG/31ABkFQ | HPLC |
| 13 | Reporter 13 | R13 | /56-FAM/ArAArAArA/31ABkFQ | HPLC |
| 14 | Reporter 14 | R14 | /56-FAM/TrUTrUTrU/31ABkFQ | HPLC |
| 15 | Reporter 15 | R15 | /56-FAM/GrGGrGGrG/31ABkFQ | HPLC |
| 16 | Reporter 16 | R16 | /56-FAM/CrCCrCCrC/31ABkFQ | HPLC |

**Table S10.** Summary of ESI–MS analysis for two representative reporters, R1 and R7.

| **Substrate: TTATT** | | | | | | | |
|---|---|---|---|---|---|---|---|
| | Formula | m/z | Retention Time | Intensity | | | |
| | | | | AsCas12a | MbCas12a | Pb2Cas12 | LbCas12a |
| T | C10H15N2O8P | 321.0493 | 3.83 | 1.22E+06 | 2.85E+07 | 2.77E+06 | 2.21E+07 |
| A | C10H14N5O6P | 330.0609 | 4.07 | 2.58E+05 | 3.27E+06 | 1.30E+06 | 3.55E+06 |
| TT | C20H28N4O15P2 | 625.0954 | 3.96 | | | | |
| TTA | C30H40N9O20P3 | 938.153 | | | | | |
| TA | C20H27N7O13P2 | 634.1069 | | | | | |
| **Substrate: CCCCCC** | | | | | | | |
| | Formula | m/z | Retention Time | Intensity | | | |
| | | | | AsCas12a | MbCas12a | Pb2Cas12 | LbCas12a |
| C | C9H14N3O8P | 322.0446 | 4.63 | 2.03E+04 | 4.03E+04 | 1.09E+04 | 3.19E+04 |
| CC | C18H26N6O15P2 | 627.0859 | 0 | 0.00E+00 | 0.00E+00 | 0.00E+00 | 0.00E+00 |
| CCC | C27H38N9O26P3 | 996.1068 | 0 | 0.00E+00 | 0.00E+00 | 0.00E+00 | 0.00E+00 |

**Table S11.** ingle-nucleotide mismatch variants of miR-21 and the corresponding DNA equivalent.

| **miR-21** | | |
|---|---|---|
| S/N | Name | RNA Sequence |
| 1 | WT-miR-21-5p | UAGCUUAUCAGACUGAUGUUGA |
| 2 | M1-miR-21-5p | GAGCUUAUCAGACUGAUGUUGA |
| 3 | M2-miR-21-5p | UCGCUUAUCAGACUGAUGUUGA |
| 4 | M3-miR-21-5p | UAUCUUAUCAGACUGAUGUUGA |
| 5 | M4-miR-21-5p | UAGAUUAUCAGACUGAUGUUGA |
| 6 | M5-miR-21-5p | UAGCGUAUCAGACUGAUGUUGA |
| 7 | M6-miR-21-5p | UAGCUGAUCAGACUGAUGUUGA |
| 8 | M7-miR-21-5p | UAGCUUCUCAGACUGAUGUUGA |
| 9 | M8-miR-21-5p | UAGCUUAGCAGACUGAUGUUGA |
| 10 | M9-miR-21-5p | UAGCUUAUAAGACUGAUGUUGA |
| 11 | M10-miR-21-5p | UAGCUUAUCCGACUGAUGUUGA |
| 12 | M11-miR-21-5p | UAGCUUAUCAUACUGAUGUUGA |
| 13 | M12-miR-21-5p | UAGCUUAUCAGCCUGAUGUUGA |
| 14 | M13-miR-21-5p | UAGCUUAUCAGAAUGAUGUUGA |

| | | |
|---|---|---|
| 15 | M14-miR-21-5p | UAGCUUAUCAGACGGAUGUUGA |
| 16 | M15-miR-21-5p | UAGCUUAUCAGACUUAUGUUGA |
| 17 | M16-miR-21-5p | UAGCUUAUCAGACUGCUGUUGA |
| 18 | M17-miR-21-5p | UAGCUUAUCAGACUGAGGUUGA |
| 19 | M18-miR-21-5p | UAGCUUAUCAGACUGAUUUUGA |
| 20 | M19-miR-21-5p | UAGCUUAUCAGACUGAUGGUGA |
| 21 | M20-miR-21-5p | UAGCUUAUCAGACUGAUGUGGA |
| 22 | M21-miR-21-5p | UAGCUUAUCAGACUGAUGUUUA |
| 23 | M22-miR-21-5p | UAGCUUAUCAGACUGAUGUUGC |
| **DNA-21** | | |
| S/N | Name | DNA Sequence |
| 1 | WT-DNA-21-5p | TAGCTTATCAGACTGATGTTGA |
| 2 | M1-DNA-21-5p | GAGCTTATCAGACTGATGTTGA |
| 3 | M2-DNA-21-5p | TCGCTTATCAGACTGATGTTGA |
| 4 | M3-DNA-21-5p | TATCTTATCAGACTGATGTTGA |
| 5 | M4-DNA-21-5p | TAGATTATCAGACTGATGTTGA |
| 6 | M5-DNA-21-5p | TAGCGTATCAGACTGATGTTGA |
| 7 | M6-DNA-21-5p | TAGCTGATCAGACTGATGTTGA |
| 8 | M7-DNA-21-5p | TAGCTTCTCAGACTGATGTTGA |
| 9 | M8-DNA-21-5p | TAGCTTAGCAGACTGATGTTGA |
| 10 | M9-DNA-21-5p | TAGCTTATAAGACTGATGTTGA |
| 11 | M10-DNA-21-5p | TAGCTTATCCGACTGATGTTGA |
| 12 | M11-DNA-21-5p | TAGCTTATCATACTGATGTTGA |
| 13 | M12-DNA-21-5p | TAGCTTATCAGCCTGATGTTGA |
| 14 | M13-DNA-21-5p | TAGCTTATCAGAATGATGTTGA |
| 15 | M14-DNA-21-5p | TAGCTTATCAGACGGATGTTGA |
| 16 | M15-DNA-21-5p | TAGCTTATCAGACTTATGTTGA |
| 17 | M16-DNA-21-5p | TAGCTTATCAGACTGCTGTTGA |
| 18 | M17-DNA-21-5p | TAGCTTATCAGACTGAGGTTGA |
| 19 | M18-DNA-21-5p | TAGCTTATCAGACTGATTTTGA |
| 20 | M19-DNA-21-5p | TAGCTTATCAGACTGATGGTGA |
| 21 | M20-DNA-21-5p | TAGCTTATCAGACTGATGTGGA |
| 22 | M21-DNA-21-5p | TAGCTTATCAGACTGATGTTTA |
| 23 | M22-DNA-21-5p | TAGCTTATCAGACTGATGTTGC |

*Red letters represent mismatches

**Table S12.** Paired double-nucleotide mismatch variants of miR-21 and the corresponding DNA equivalent.

| **miR-21** | | |
|---|---|---|
| S/N | Name | Sequence |
| 1 | WT-miR-21-5p | UAGCUUAUCAGACUGAUGUUGA |
| 2 | M1-2_miR-21-5p | GCGCUUAUCAGACUGAUGUUGA |
| 3 | M3-4_miR-21-5p | UAUAUUAUCAGACUGAUGUUGA |
| 4 | M5-6_miR-21-5p | UAGCGGAUCAGACUGAUGUUGA |

| | | |
|---|---|---|
| 5 | M7-8_miR-21-5p | UAGCUUCGCAGACUGAUGUUGA |
| 6 | M9-10_miR-21-5p | UAGCUUAUACGACUGAUGUUGA |
| 7 | M11-12_miR-21-5p | UAGCUUAUCAUCCUGAUGUUGA |
| 8 | M13-14_miR-21-5p | UAGCUUAUCAGAAGGAUGUUGA |
| 9 | M14-16_miR-21-5p | UAGCUUAUCAGACUUCUGUUGA |
| 10 | M17-18_miR-21-5p | UAGCUUAUCAGACUGAGUUUGA |
| 11 | M19-20_miR-21-5p | UAGCUUAUCAGACUGAUGGGGA |
| 12 | M21-22_miR-21-5p | UAGCUUAUCAGACUGAUGUUUC |
| **DNA-21** | | |
| S/N | Name | Sequence (DNA Equivalence) |
| 1 | WT-miR-21-5p | TAGCTTATCAGACTGATGTTGA |
| 2 | M1-2_miR-21-5p | GCGCTTATCAGACTGATGTTGA |
| 3 | M3-4_miR-21-5p | TATATTATCAGACTGATGTTGA |
| 4 | M5-6_miR-21-5p | TAGCGGATCAGACTGATGTTGA |
| 5 | M7-8_miR-21-5p | TAGCTTCGCAGACTGATGTTGA |
| 6 | M9-10_miR-21-5p | TAGCTTATACGACTGATGTTGA |
| 7 | M11-12_miR-21-5p | TAGCTTATCATCCTGATGTTGA |
| 8 | M13-14_miR-21-5p | TAGCTTATCAGAAGGATGTTGA |
| 9 | M14-16_miR-21-5p | TAGCTTATCAGACTTCTGTTGA |
| 10 | M17-18_miR-21-5p | TAGCTTATCAGACTGAGTTTGA |
| 11 | M19-20_miR-21-5p | TAGCTTATCAGACTGATGGGGA |
| 12 | M21-22_miR-21-5p | TAGCTTATCAGACTGATGTTTC |

*Red letters represent mismatches

**Table S13.** LAMP primer sequences for Influenza A, Influenza B, and RSV A, together with corresponding forward and reverse miniCaRs.

| **Kubo et al. 2010 (original paper)** | **Flu A_H1N1** |
|---|---|
| Name | Sequence (5'-3') |
| F3_H1N1_FluA | GCTAAGAGAGCAATTGAGC |
| B3_H1N1_FluA | ATGTAGGATTTGCTGAGCT |
| FIP_H1N1_FluA | CGAGTCATGATTGGGCCATGACAGTGTCATCATTTGAAAGGTTT |
| BIP_H1N1_FluA | AAGGTGTAACGGCAGCATGTCCGAATTTCCTTTTTTAACTAGCCAT |
| LF_H1N1_FluA | ACTTGTCTTGGGGAATATCTC |
| LB_H1N1_FluA | ATGCTGGAGCAAAAAGCT |
| gBlock | GAGCTAAGAGAGCAATTGAGCTCAGTGTCATCATTTGAAAGGTTTGAGATATTCCCCAAGACAAGTTCATGGCCCAATCATGACTCGAACAAAGGTGTAACGGCAGCATGTCCTCATGCTGGAGCAAAAAGCTTCTACAAAAATTTAATATGGCTAGTTAAAAAAGGAAATTCATACCCAAAGCTCAGCAAATCCTACATTA |
| Forward LAMP loop FluA | CAGTGTCATCATTTGAAAGGTTTGAGATATTCCCCAAGACAAGT |
| Forward FluA miniCaR | AUGAACUUGUCUUGGGGAAUAU |
| Reverse LAMP Loop FluA | GAATTTCCTTTTTTAACTAGCCATATTAAATTTTTGTAGAAGCTTTTTGCTCCAGCATGA |
| Reverse FluA miniCaR | AAAAAGCUUCUACAAAAAUUUA |

| | |
|---|---|
| **Jang et al. 2020 (original paper)** | **Flu B** |
| Name | Sequence (5'-3') |
| F3_FluB | GAGCTGCCTATGAAGACC |
| B3_FluB | CGTCTCCACCTACTTCGT |
| FIP_FluB | GAACATGGAAACCCTTGCATTTTAAGTTTTGTCTGCATTAACAGGC |
| BIP_FluB | GAACAGRTRGAAGGAATGGGRGCGATCTGGTCATTGGAGCC |
| LF_FluB | TGCTGATCTAGGCTTGAATTCTGT |
| LB_FluB | AGCTCTGATGTCCATCAAGCTCC |
| BIP | GAACAGRTRGAAGGAATGGGRGCGATCTGGTCATTGGAGCC |
| gBlock | CGGAGCTGCCTATGAAGACCTGAGAGTTTTGTCTGCATTAACAGGCACAGAATTC AAGCCTAGATCAGCATTAAAATGCAAGGGTTTCCATGTTCCAGCAAAGGAACAGG TAGAAGGGATGGGAGCAGCTCTGATGTCCATCAAGCTCCAGTTTTGGGCTCCGAT GACCAGATCTGGGGGGAACGAAGTAGGTGGAGACGGA |
| Forward LAMP loop FluB | GTTTTGTCTGCATTAACAGGCACAGAATTCAAGCCTAGATCAGCA |
| Forward FluB miniCaR | UUAAUGCUGAUCUAGGCUUGAA |
| Reverse LAMP Loop FluB | GATCTGGTCATTGGAGCCCAAAACTGGAGCTTGATGGACATCAGAGCT |
| Reverse FluB miniCaR | UGAUGUCCAUCAAGCUCCAGUU |
| **Mahony et al. 2013 (original paper)** | **RSV A** |
| Name | Sequence (5'-3') |
| F3_RSVA | GCTGTTCAATACAATGTCCTAGA |
| B3_RSVA | GGTAAATTTGCTGGGCATT |
| FIP_RSVA | TCTGCTGGCATGGATGATTGGAGACGATGATCCTGCATCA |
| BIP_RSVA | CTAGTGAAACAAATATCCACACCCAGCACTGCACTTCTTGAGTT |
| LF_RSVA | ACATGGGCACCCATATTGTAAG |
| LB_RSVA | AGGGACCTTCATTAAGAGTCATGAT |
| gBlock | CTGCTGTTCAATACAATGTCCTAGAAAAAGACGATGATCCTGCATCACTTACAATA TGGGTACCCATGTTCCAATCATCCTTGCCAGCAGATCTACTCATAAAAGAACTAGC CAATGTTAATATACTAGTGAAACAAATATCCACACCCAAGGGACCCTCATTAAGAG TCATGATAAACTCAAGAAGTGCAGTGCTAGCACAAATGCCCAGCAAATTTACCAT |
| Forward LAMP loop RSV A | GACGATGATCCTGCATCACTTACAATATGGGTACCCATGT |
| Forward RSV A miniCaR | AUGGGUACCCAUAUUGUAAGUGA |
| Reverse LAMP Loop RSV A | CAGCACTGCACTTCTTGAGTTTATCATGACTCTTAATGAGGGTCCCTTG |
| Reverse RSV A miniCaR | UCAUUAAGAGUCAUGAUAAACU |

**Table S14.** Cas12a proteins used in this study.

| Protein | Plasmid | Tags |
|---|---|---|
| AsCas12a | https://www.addgene.org/113430/ | 10x His - MBP - TEV (N term) |
| BoCas12a | https://www.addgene.org/174673/ | 6x His - MBP - TEV (N term) NLS - 6x His - 3xHA (C term) |
| BsCas12a | https://www.addgene.org/174671/ | 6xHis - MBP - TEV - NLS (N term) NLS - 3xHA - T2A - EGFP - 6xHis (C term) |
| ErCas12a | https://www.addgene.org/174685/ | 6xHis - MBP - TEV - NLS (N term) NLS - 6xHis (C term) |
| FnCas12a | https://www.addgene.org/113432/ | 10xHis - MBP - TEV (N term) |
| LbCas12a | https://www.addgene.org/113431/ | 10xHis - MBP - TEV (N term) |
| Lb5Cas12a | https://www.addgene.org/174684/ | 6x His - MBP - TEV (N term) NLS - 6x His - 3xHA (C term) |
| MbCas12a | https://www.addgene.org/115670/ | 10xHis - MBP - TEV (N term) |
| Pb2Cas12a | https://www.addgene.org/174686/ | 6xHis - MBP - TEV (N term) NLS - 6xHis - 3xHA (C term) |
| TsCas12a | https://www.addgene.org/174687/ | 6x His - MBP - TEV (N term) NLS - 6x His - 3xHA (C term) |
| LbCas12a | https://www.addgene.org/114070/ | NLS - 6x His (C term) |

**Table S15**. DNA templates used as *trans* cleavage substrates in eCOMPANION.

| Name | Sequence (5´ – 3´) |
|---|---|
| C-rich Reporter Sequence | /5ThioMC6-D/CCCCCCCCCCCCCCCCCCCCCCCCCCC |
| A-rich Reporter Sequence | /5ThioMC6-D/ACAATTATACTCAGCAAAAAAAAAAAA |

**Table S16.** Diagnostic performance of COMPANION for FluA (H1N1) detection in respiratory patient samples.

| | RT-qPCR - | RT-qPCR + | Total |
|---|---|---|---|
| COMPANION - | 30 | 0 | 30 |
| COMPANION + | 0 | 9 | 9 |
| Total | 30 | 9 | 39 |
| Sensitivity | 100% (95% CI 66.37% to 100%) | | |
| Specificity | 100% (95% CI 88.43% to 100%) | | |
| Disease prevalence | 23.08% (95% CI 11.13% to 39.33%) | | |
| Positive Predictive Value (PPV) | 100% (95% CI 66.37% to 100%) | | |
| Negative Predictive Value (NPV) | 100% (95% CI 88.43% to 100%) | | |
| Accuracy | 100% (95% CI 90.97% to 100%) | | |

**Table S17.** Performance evaluation of COMPANION for detection of FluB in respiratory patient samples.

| | RT-qPCR - | RT-qPCR + | Total |
|---|---|---|---|
| COMPANION - | 15 | 1 | 16 |
| COMPANION + | 0 | 22 | 22 |
| Total | 15 | 23 | 38 |
| Sensitivity | 95.65% (95% CI 78.05% to 99.89%) | | |
| Specificity | 100% (95% CI 78.20% to 100%) | | |
| Disease prevalence | 60.53% (95% CI 43.39% to 75.96%) | | |

| | |
|---|---|
| Positive Predictive Value (PPV) | 100% (95% CI 84.56% to 100%) |
| Negative Predictive Value (NPV) | 93.75% (95% CI 68.81% to 99.03%) |
| Accuracy | 97.37% (95% CI 86.19% to 99.93%) |

**Table S18.** Diagnostic performance of COMPANION for RSV A detection in respiratory patient samples.

| | RT-qPCR - | RT-qPCR + | Total |
|---|---|---|---|
| COMPANION - | 23 | 0 | 23 |
| COMPANION + | 0 | 2 | 2 |
| Total | 23 | 2 | 25 |
| Sensitivity | 100% (95% CI 15.81% to 100%) | | |
| Specificity | 100% (95% CI 85.18% to 100%) | | |
| Disease prevalence | 8.0% (95% CI 0.98% to 26.03%) | | |
| Positive Predictive Value (PPV) | 100% (95% CI 15.81% to 100%) | | |
| Negative Predictive Value (NPV) | 100% (95% CI 85.18% to 100%) | | |
| Accuracy | 100% (95% CI 86.28% to 100%) | | |
